# Human Trafficking Detection in Health Care Settings: A Scoping Review

**DOI:** 10.1101/2025.10.01.25336367

**Authors:** Silvia Villatoro, Pamela Velásquez, David García, Claudia Marcela Vélez, Daniel Felipe Patiño, Esther Ekpe Adewuyi, Andrea Burkhart, Paola Ramírez, Claudia Lorena Ramírez, Iván D. Flórez

**Author notes:** Corresponding author; email: Iván Darío Flórez.

## Abstract

**Objectives:** This scoping review aimed to identify available tools for detecting human trafficking survivors in healthcare settings and to explore implementation strategies.

**Methods:** A systematic search was conducted in MEDLINE/PubMed, EMBASE, BIREME-LILACS, WHO-PAHO IRIS, and other institutional repositories on August 12, 2024, with an update on January 18, 2025.

**Results:** Out of 2,881 records screened, 140 resources were included, mostly from the U.S. (n = 113). We identified 26 screening tools, 44 toolkits and guidelines, 23 documents on educational strategies and 24 studies on other types of strategies targeting HT detection in healthcare. Tools often addressed sexual exploitation, especially in minors, with limited focus on other forms of trafficking. Only eight tools reported having undergone validation processes. Many resources emphasized trauma-informed care, indicator use, and referral protocols. Implementation strategies included training programs, integration of screening protocols into clinical workflows, and digital tools; however, system-level barriers and limited provider confidence persist.

**Conclusion:** Despite the availability of tools and guidelines, there is no consensus on definitions or standardized methods for HT identification. Most tools focus on sexual exploitation, particularly in minors, while other trafficking forms are under-addressed. Sustained training, validated tools, and interdisciplinary collaboration are essential. Systemic barriers must be addressed through clear protocols and institutional commitment to ensure effective, survivor-centered detection and care.

## Introduction

Human trafficking (HT) is recognized globally as both a serious criminal offense and a grave violation of human rights. It affects nearly every country, whether as a point of origin, transit, or destination for trafficked individuals. In 2020 alone, over 51,000 cases were identified across 160 countries, with approximately 65% of survivors being women and girls (1,2). Between 2020 and 2022, the number of detected cases rose by 43%, and preliminary figures for 2023 suggest an ongoing upward trend. Of the 202,478 cases reported in 2022, 62% involved adults and 38% involved minors. Notably, women and girls made up a larger proportion of survivors across all age groups, with 39% identified as adult women and 22% as girls, compared to 23% as adult men and 16% as boys (2). While there is no single or typical profile of a trafficking survivor, certain populations—particularly those facing social exclusion, economic vulnerability, or limited empowerment—are known to be at greater risk (1–4).

According to the Palermo Protocol, HT involves recruitment, transportation, transfer, harboring, or receipt of individuals by means of force, coercion, abduction, fraud, or deception for the purpose of exploitation (1). Despite this internationally agreed-upon definition, trafficking is often misunderstood or conflated with migrant smuggling, a separate offense characterized by consent and typically ending upon arrival at the destination. In contrast, HT may not involve cross-border movement and occurs against the individual’s will (3).

Over time, the legal and political discourse surrounding HT has evolved, leading to broader and often contested interpretations of the crime. Terms such as “modern slavery,” “sexual exploitation,” and “forced labor” are frequently used inconsistently in policy, media, and even academic literature. Legal scholar Janie Chuang has termed this phenomenon the “expansion of exploitation,” where diverse and sometimes unrelated practices—such as consensual sex work, precarious labor, or irregular migration—are subsumed under the trafficking umbrella, even in the absence of coercion or force (5). In addition, although sexual exploitation is not synonymous with HT, and not all HT involves sexual exploitation, the literature frequently uses these terms interchangeably (6). This definitional ambiguity can lead to inappropriate legal or policy responses and can potentially result in the criminalization of vulnerable populations such as migrants, sex workers, or racialized women (7,8).

In the context of healthcare, this ambiguity becomes particularly relevant. While sexual exploitation and forced labor are among the most common forms of trafficking encountered in medical settings, the conflation of terminology may obscure clinical recognition. This lack of clarity may prevent healthcare professionals from identifying red flags, delaying timely intervention, and leaving serious physical and mental health needs unaddressed. Given that victims often present with complex medical, psychological, and social consequences, precise terminology and a clear understanding of trafficking typologies are essential for accurate diagnosis, appropriate referral, and effective prevention within healthcare systems. To ensure thoroughness and accurate representation of the state of the field, this review includes tools and studies that use such terminology—even if the terms themselves are undergoing critical reassessment.

Healthcare settings play a crucial role as a gateway for identifying and supporting individuals affected by trafficking. Survivors often access care for physical injuries, infections, chronic conditions, and mental health issues while still in exploitation, with up to 88% of those subjected to sexual exploitation reporting contact with healthcare services during their trafficking experience (9). This means the healthcare system is one of the few places where victims can be identified and helped, and healthcare professionals are key frontline responders in early detection and intervention. However, healthcare systems have historically lacked adequate training, validated tools, and trauma-informed protocols to recognize trafficking, leading to missed opportunities for intervention (9,10).

Importantly, the health consequences of HT are profound. Survivors frequently endure physical abuse, sexual violence, psychological trauma, and chronic medical conditions, compounded by social and economic marginalization. Up to 99% of those subjected to trafficking—especially sexual exploitation—report physical (e.g., neurological, dental, gastrointestinal) and mental health symptoms (e.g., depression, Post-Traumatic Stress Disorder, suicidal ideation) (10,11). Despite the gravity of these outcomes, healthcare professionals often miss opportunities to identify people in trafficking situations (10). In part, this is due to the absence of validated screening tools, inconsistent terminology, and the lack of integrated training and protocols.

Therefore, the early and accurate identification of trafficking survivors in healthcare settings is not only ethically imperative but also a critical component of victim-centered care. Screening tools—when properly validated and contextually adapted—can assist clinicians in recognizing potential trafficking cases and linking survivors to appropriate support services. However, many tools currently used in practice lack rigorous psychometric or diagnostic validation, raising concerns about their reliability and effectiveness.

Previous scoping reviews, such as that by Hainaut et al. (12) have examined the validation and diagnostic properties of trafficking detection tools in healthcare settings. Building on these efforts, this review adds value by also mapping toolkits, guidelines, protocols, and indicator lists, and by synthesizing implementation strategies to support the practical integration of screening and survivor-centered care within healthcare systems.

This scoping review was conducted to: (1) identify available screening tools for the detection of HT survivors in healthcare settings; (2) assess their reported effectiveness, validation status, and diagnostic properties; and (3) synthesize available strategies to support their implementation. Our goal was to provide a comprehensive and nuanced synthesis to support evidence-based decision-making in clinical practice.

## Methods

### Protocol

This scoping review was conducted following the JBI 2020 (formerly Joanna Briggs Institute) guidance for scoping reviews (13) and is reported in accordance with the Preferred Reporting Items for Systematic Reviews and Meta-Analyses extension to scoping reviews (PRISMA-ScR) (14). The protocol was prospectively registered in the Open Science Framework (https://osf.io/nbk53/).

### Literature Search

We systematically searched for evidence syntheses (i.e., systematic reviews, scoping reviews, rapid reviews), primary studies, organizational reports, and reference documents cited in other works. An experienced librarian developed and iteratively refined the search strategies in consultation with the review team. Searches were tailored to each database, and complete strategies are provided in Appendix 1. Databases searched included MEDLINE/PubMed, Embase, Bireme-LILACS, WHO- PAHO IRIS, and institutional repositories. Initial searches were conducted on August 12, 2024, and were updated on January 18, 2025.

### Question and Study Selection

Titles and abstracts were screened independently and in duplicate using Rayyan (15) by trained reviewers (DG, LR, PV, SV). Full texts of the eligible records were also retrieved and reviewed independently and in duplicate by the same reviewers. Prior to conducting the review and throughout the process, the team completed training sessions (provided by PV, SV) to ensure consistency in applying eligibility criteria and data extraction procedures. Definitions related to trafficking, typologies, and response strategies are outlined in Appendix 2, based on the United Nations Office on Drugs and Crime (UNODC) framework (16). No language or date restrictions were applied.

We followed the PCC framework suggested by the JBI guidance (13), to frame our question. The population of interest was patients potentially experiencing human trafficking in healthcare settings, and the healthcare workers serving them. The concepts to identify were tools, strategies, and protocols aimed at identifying potential victims of human trafficking. We focused solely on the healthcare setting context, excluding all other contexts. We only included evidence syntheses, qualitative and quantitative primary research, as well as official reports.

### Data Extraction

Data were extracted using a standardized Google Form, refined iteratively through team discussion. Reviewers underwent training before full extraction. A single reviewer with verification approach was used, with 10% of included studies randomly verified by a second reviewer (DG, LR, PV, SV).

For evidence syntheses we extracted the following information title, authors, focus, search dates, tool or strategy features, outcomes, and key findings. For primary studies, we obtained design type, concepts, outcomes, characteristics of tools/strategies, validation status, and findings. For reports or policy documents, we documented: issuing body, publication year, scope, population, tools/strategies addressed, and recommendations.

### Data Synthesis

Data were synthesized narratively and descriptively. The extracted data were structured using the UNODC “International Framework for Action to Implement the Trafficking in Persons Protocol” (16) and the Cochrane Effective Practice and Organisation of Care (EPOC) taxonomy (17). Categories and interpretations were developed through team consensus.

### Results Presentation

Findings are presented corresponding to the following key themes: Screening tools to identify human trafficking; Validation studies of tools; Toolkits, guides and protocols for identification and survivor support; Indicators used in detection and Implementation strategies.

### Patient and Public Involvement

As per the Strategy for Patients Outcomes Research (SPOR) network approach (18,19), which emphasizes meaningful patient and citizen partner engagement to ensure research relevance and applicability, two citizen partners were engaged throughout this scoping review (EEA and AB). Their roles included contributing to the development of the protocol, search strategies, and data extraction form; reviewing the final report; identifying key findings; and co-producing a key message for patients/citizens and a plain language summary; Therefore, the partners are co-authors of this scientific publication.

## Results

After screening 2,881 records identified through electronic database searches, 415 full-text articles were assessed for eligibility. Of these, 275 were excluded, with reasons detailed in the PRISMA flow diagram (Figure 1). A total of 140 records were included in the final synthesis. Of these, 57 were identified through database searches and 83 through organizational portals and other sources of grey literature.

**Figure 1.**
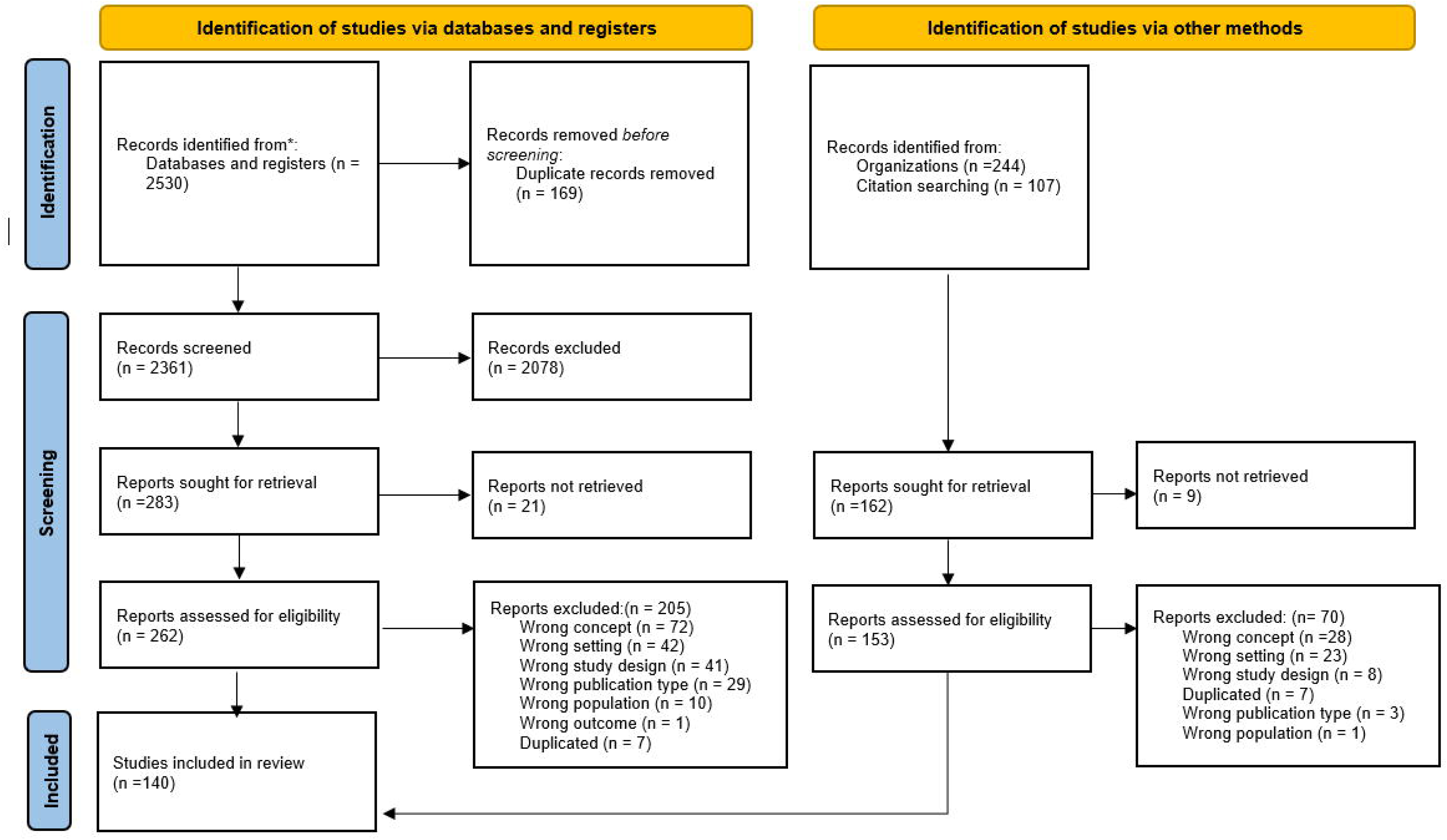
PRISMA 2020 flow diagram for new systematic reviews which included searches of databases, registers and other sources

### Study characteristics

The included resources were predominantly from the United States (n = 113), followed by the United Kingdom (n = 13), Switzerland (n = 5), Spain (n = 3), and Canada (n = 3), and individual contributions one study each from Denmark, Brazil, and Ireland (n = 1 each). In terms of study type, 47 were primary studies, including cohort studies (n = 7), case-control (n = 1), cross-sectional (n = 5), qualitative (n = 9), mixed methods (n = 1), quasi-experimental (n = 12), randomized controlled trials (RCTs) (n = 1), and studies focused on validation or evaluation of diagnostic properties (n = 8).

Eleven documents were categorized as general reports, and 55 corresponded to tools, guidelines, or direct evaluations thereof. Additionally, nine were systematic reviews, five were scoping reviews, and two were integrative reviews. Regarding the focus of the studies, the majority addressed human trafficking in general (n = 92), while others focused specifically on sexual trafficking (n = 41), or on sexual and labor trafficking (n = 7). Concerning the target population, 52 resources addressed minors, seven focused exclusively on adults, and 81 included both age groups or did not specify age.

### Screen Tools for Human Trafficking

A total of 26 screening tools were identified across 19 reports (20–38) (Table 1.1 and 1.2), five primary studies (39–43), and 12 systematic or scoping reviews (44–46,12,47–54). Most tools were developed in the United States (n = 19), with additional contributions from the United Kingdom (n = 2), Spain (n = 1), Canada (n = 1), and Ireland (n = 1). In terms of trafficking type, seven tools specifically target sex trafficking, while five focus on the sexual exploitation of minors, particularly within the broader context of human trafficking (e.g., the Child Exploitation Screening Tool, WestCoast CSE-IT). No tool was exclusively dedicated to identifying labor trafficking; however, 18 tools detect human trafficking in a general form, encompassing multiple types of exploitation, including sexual and labor trafficking (e.g., TVIT). Eight tools reported having undergone validation studies. Among these, two are designed for general human trafficking identification, while six focus specifically on detecting sexual exploitation.

**Table 1.1.** Screen Tools for Human Trafficking Validated.

| **Tool name**<br>**Organization / Author / Country** | **Detection**<br>**focus** | **Target**<br>**population** | **Tool structure** | **Application**<br>**context** | **Evaluation of**<br>**its effectiveness**<br>**or**<br>**performance.** | **PMID or URL or DOI** |
| --- | --- | --- | --- | --- | --- | --- |
| **A Short Screening Tool to Identify Victims of Child Sex Trafficking in the Health Care Setting**<br>Greenbaum, V. J. 2018<br>U.S. | Sex trafficking | Children and Adolescents | Standardized questionnaire | General healthcare settings | Effectiveness studies/<br>Diagnostic and prognostic property testing/<br>Usage experience | https://journals.lww.com/pec-online/abstract/2018/01000/a_short_screening_tool_to_identify_victims_of.8.aspx |
| **Child Exploitation Screening Tool**<br>Oxfordshire Safeguarding Children Board, 2023<br>UK | Sex trafficking | Children and Adolescents | Standardized questionnaire | Multiple contexts, adaptable to healthcare settings | Not reported | https://www.oscb.org.uk/wp-content/uploads/2024/05/CE-Screening-Tool-1.pdf |
| **Clinical CSEC Screening Tool**<br>Sy, et al., 2016<br>Asian Health Services (AHS) and Banteay Srei<br>U.S. | Sex trafficking | Children and Adolescents | List of indicators and standardized questionnaire | General healthcare settings | Not reported | https://doi.org/10.11114/ijsss.v4i6.1576 |
| **Eina de Detecció del Risc d'Explotació Sexual en la Infància i la Adolescència (EDR-ESIA tool)**<br>Benavente, B., 2022<br>Spain | Sex trafficking | Children and Adolescents | Check list of risk indicators | Multiple contexts, adaptable to healthcare settings | Diagnostic or prognostic property testing | http://www.papelesdelpsicologo.es/pii?pii=2993 |
| **Rapid Appraisal for Trafficking (RAFT)**<br>Chisolm-Straker, 2021<br>U.S. | HT in general | Adults | Standardized questionnaire | General healthcare settings | Effectiveness studies/<br>Diagnostic and prognostic property testing<br>Usage experience | [https://www.jacepopen.com/article/S2688-1152(24)00974-3/fulltext](https://www.jacepopen.com/article/S2688-1152%2824%2900974-3/fulltext) |
| **Screening survey to identify of victims of sex trafficking**<br>Mumma, B., 2017 | Sex trafficking | Adults | Standardized questionnaire | General healthcare settings | Effectiveness studies | https://pmc.ncbi.nlm.nih.gov/articles/PMC5468066/ |
| **Trafficking Victim Identification Tool (TVIT)**<br>VERA National Institute of Justice<br>Simich, L. 2014<br>U.S. | HT in general | Applicable to different age groups | Standardized questionnaire | Multiple contexts, adaptable to healthcare settings | Effectiveness studies and prognostic property testing | https://nij.ojp.gov/library/publications/improving-human-trafficking-victim-identification-validation-and-dissemination |
| **WestCoast's Commercial Sexual Exploitation-Identification Tool (CSE-IT)**<br>Basson D (WestCoast Children's Clinic), 2017<br>U.S. | Sex trafficking | Children and Adolescents | Standardized questionnaire | Multiple contexts, adaptable to healthcare settings | Not reported | https://www.westcoastcc.org/wp-content/uploads/2015/04/WCC-CSE-IT-PilotReport-FINAL.pdf |

HT: Human Trafficking, UK: United Kingdom, U.S.: United States

**Table 1.2.**
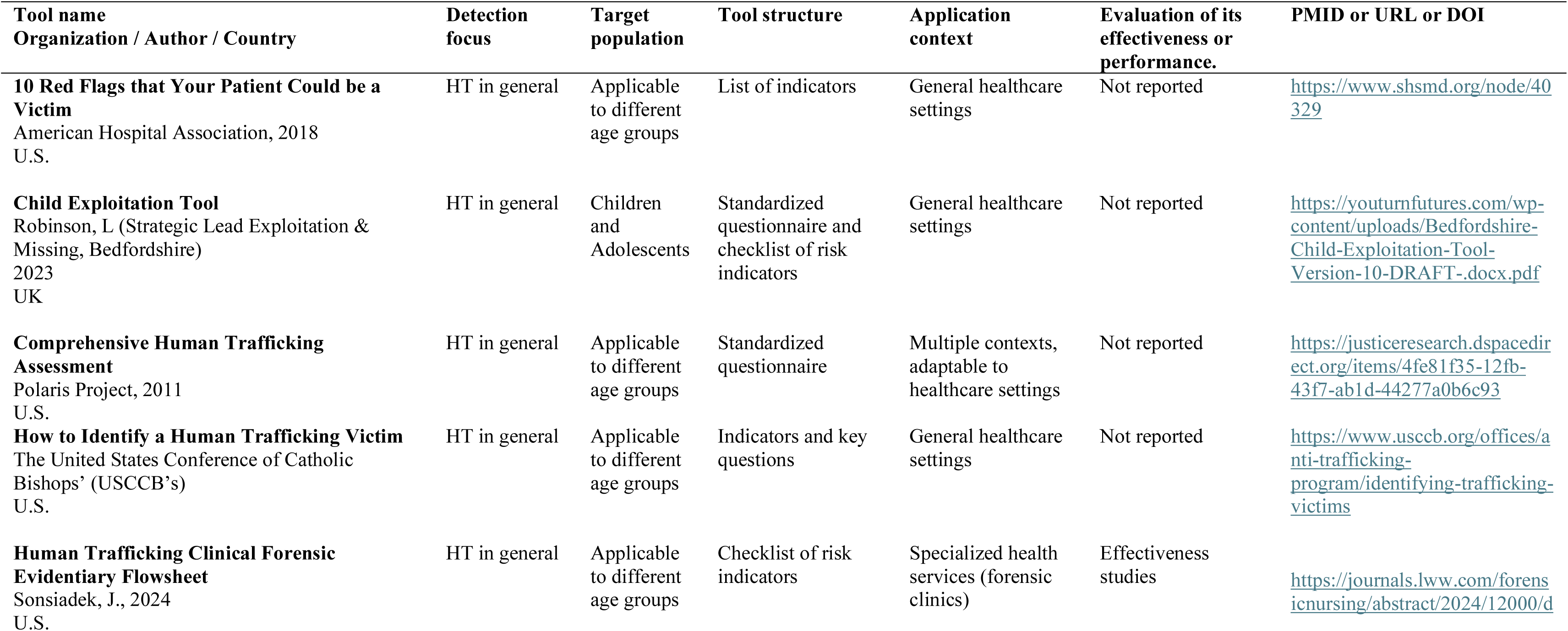
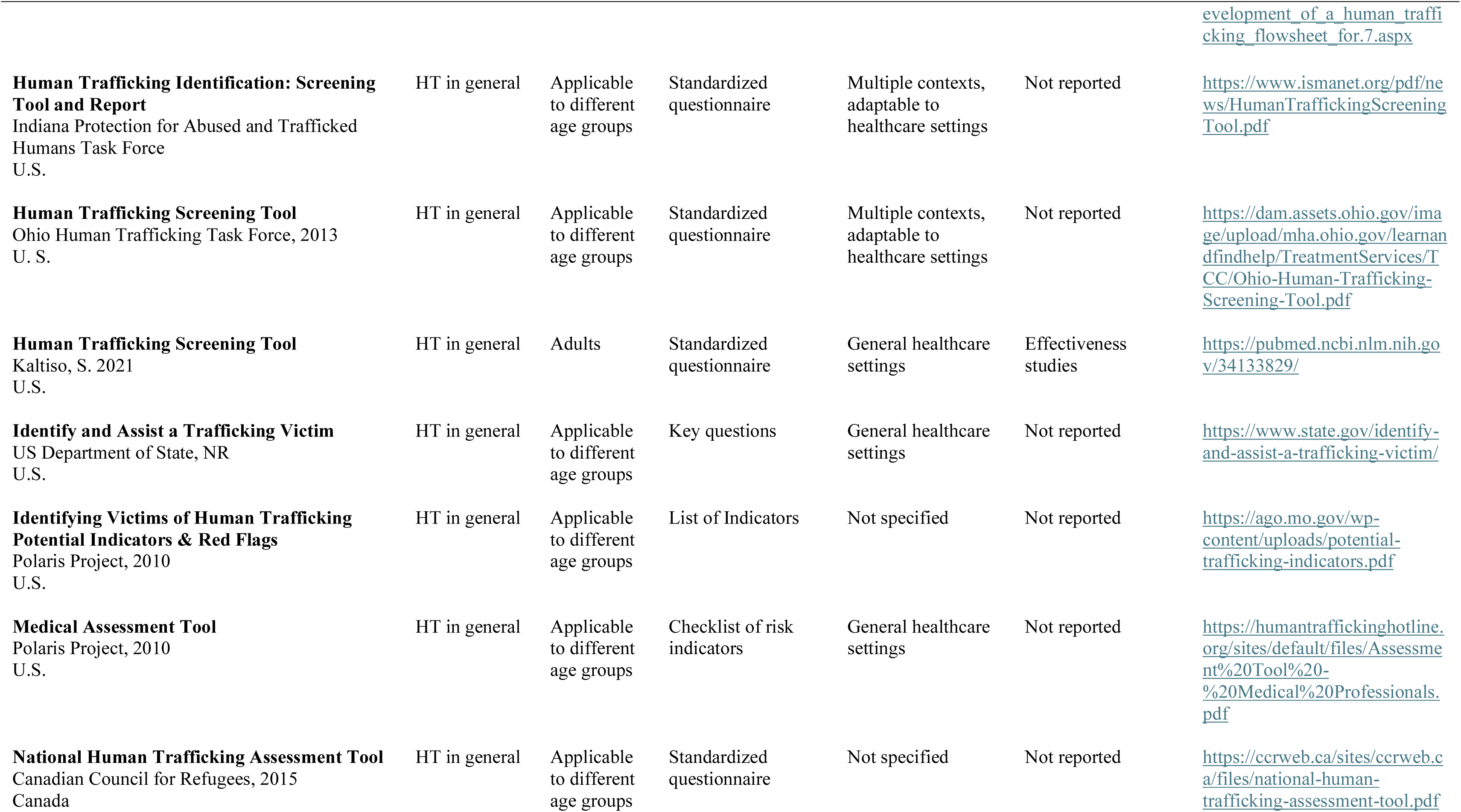
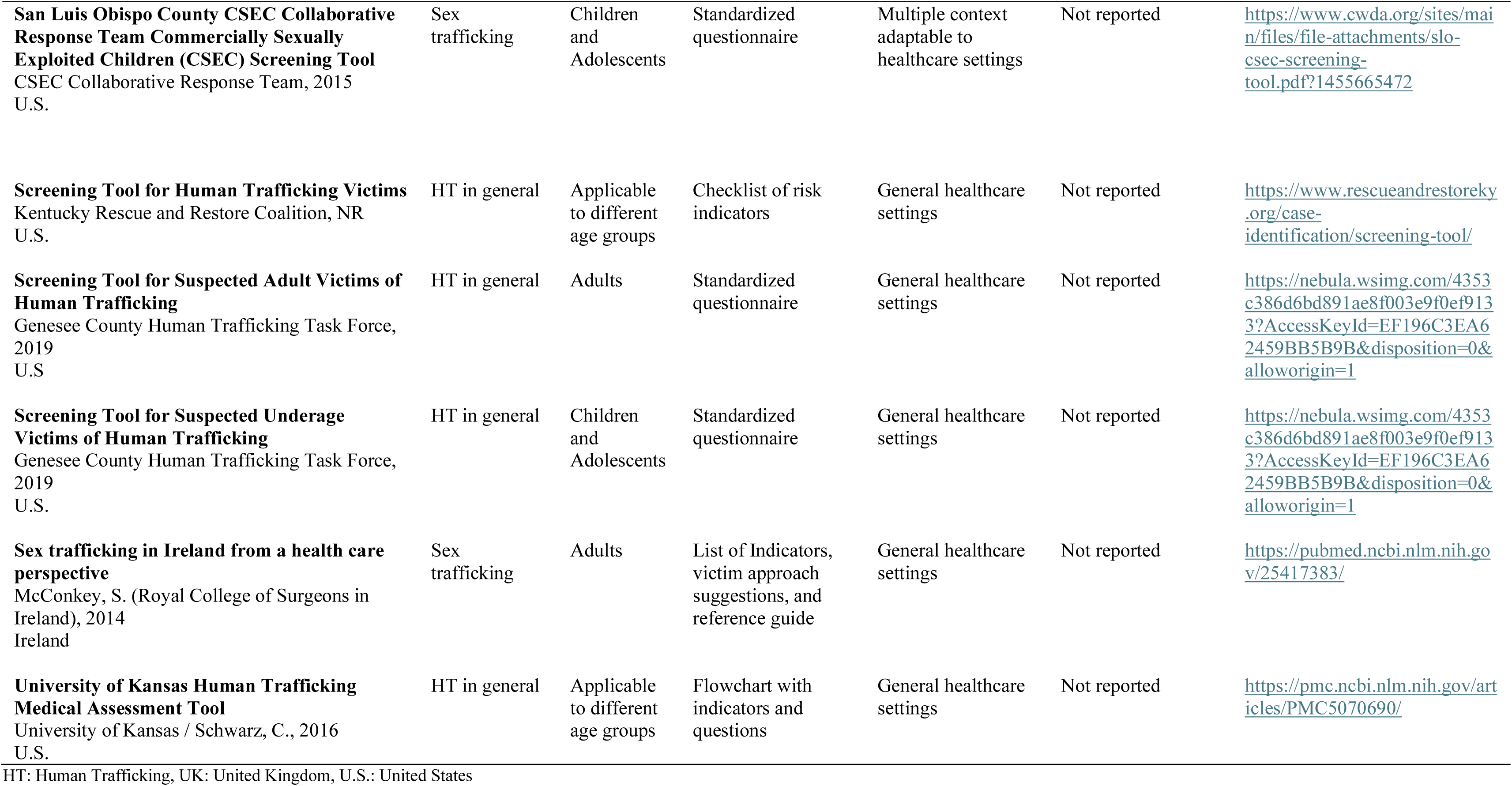
Screen Tools for Human Trafficking Non-validated.

Regarding the target population of the tools, 13 tools were designed to identify trafficking across children, adolescents, and adults, with one focusing exclusively on sex trafficking. Four tools targeted adult victims, including the Medical Assessment Tool specific to sex trafficking. Eight tools were designed solely for individuals under the age of 18; seven of these focused on sexual exploitation, while one also included broader trafficking categories (e.g., Child Exploitation Tool). Structurally, 16 tools used standardized questionnaires, eight relied on indicator checklists, and three offered a concise set of key screening questions. Eleven tools included fewer than 20 questions or indicators, facilitating rapid assessment. Three tools required specialized training for effective implementation, as noted by their authors. In addition to the tools explicitly analyzed, systematic reviews (12,44–47,50,51) identified 16 additional instruments suitable for use in healthcare contexts, although not all have been independently validated (Appendix 2).

### Validation Studies and Diagnostic Performance

A total of ten studies were identified in this review that evaluated the validity, reliability, or diagnostic performance of screening tools for human trafficking in healthcare settings (38–40,42,55–59). Among them, three provided formal evidence of validity testing (38,40,42), while the remaining six focused on evaluating diagnostic accuracy alone—such as sensitivity, specificity, or predictive performance. Five of these studies assessed the Greenbaum 6-Question Short Screening Tool for adolescents in pediatric emergency departments and child advocacy centers (39,55–58). The results suggest that using a threshold of two positive responses provides a good balance of high sensitivity (80–92%) and moderate specificity (50–75%), effectively identifying most trafficking victims while accepting a degree of false positives. A threshold of two positive responses optimized sensitivity (80–92%) while maintaining moderate specificity (50–75%), ensuring victim detection while tolerating some false positives.

The Human Trafficking Screening Tool (HTST) was tested in 26,974 adult emergency department (ED) patients, showing high predictive accuracy (AUROC 0.85), though some false positives occurred, primarily among domestic violence cases (41). One study by Mumma et al. (2017) showed that a single screening question—“Have you (or anyone you work with) ever been beaten, yelled at, raped, threatened, or made to feel physical pain for working slowly or trying to leave?”—identified 100% of known survivors with 78% specificity, outperforming physician-based detection rates (40%) (59). The Commercial Sexual Exploitation–Identification Tool (CSE-IT) (38), the Trafficking Victim Identification Tool (TVIT) (42), and the Rapid Appraisal for Trafficking (RAFT)(40) are the only screening tools with formal validation. All three demonstrated strong criterion validity, while RAFT also showed robust construct validity and consistent performance across populations. Although only CSE-IT and TVIT reported reliability testing, the internal consistency of RAFT supports its potential utility in clinical settings (Appendix 2).

### Toolkits and Guidelines for Identifying and Supporting Survivors of Human Trafficking

A total of 44 toolkits, protocols, and guidance documents for identifying victims of human trafficking were reviewed (3,60–102) (Tables 2.1 and 2.2). Most were developed in the United States (n = 31), with others originating from the United Kingdom (n = 8), Switzerland (n = 4), and Brazil (n = 1). While the majority were created for use in healthcare settings (n = 29), several toolkits and guidance developed for other settings—such as child protection or community services—explicitly indicate that they have been used in, or are adaptable to, healthcare contexts (n = 15). In terms of detection focus, most toolkits and guidance address human trafficking in general (n = 31), meaning they encompass a broad range of exploitative forms including sexual, labor, and others. A smaller subset targets only sexual and labor trafficking (n = 3) without addressing broader forms of exploitation. Additionally, ten toolkits and guidance focus specifically on sexual trafficking, with particular emphasis on commercial sexual exploitation of children. Regarding target populations, 26 toolkits and guidance are applicable to all age groups, 17 focus specifically on children and adolescents, and one is designed for adult populations. Most emphasize practical implementation and include red flag indicators, trauma-informed care strategies, legal considerations, and referral pathways. Shared principles across these resources include a victim-centered approach, privacy and confidentiality, interdisciplinary collaboration, and healthcare provider training.

**Table 2.1.**
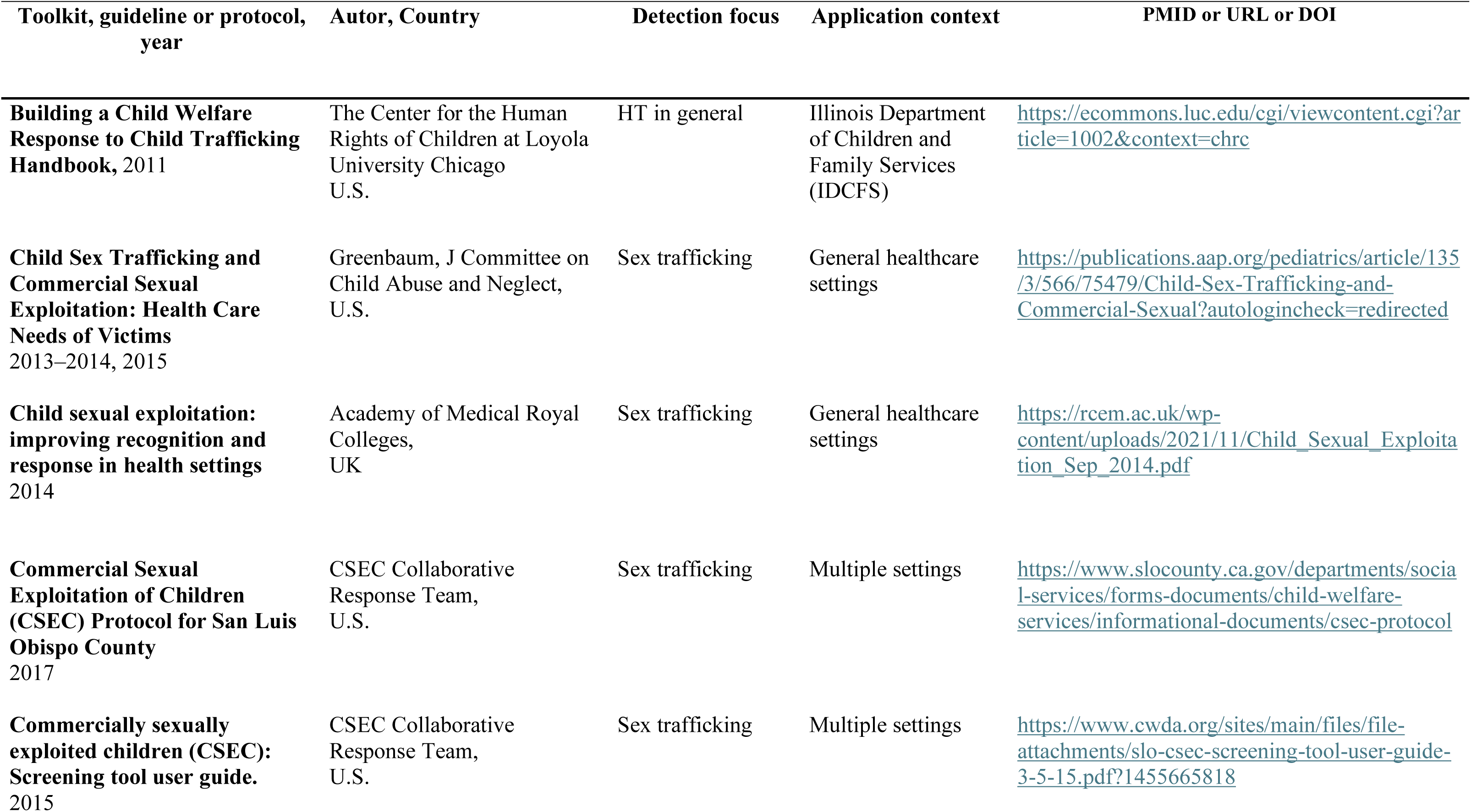
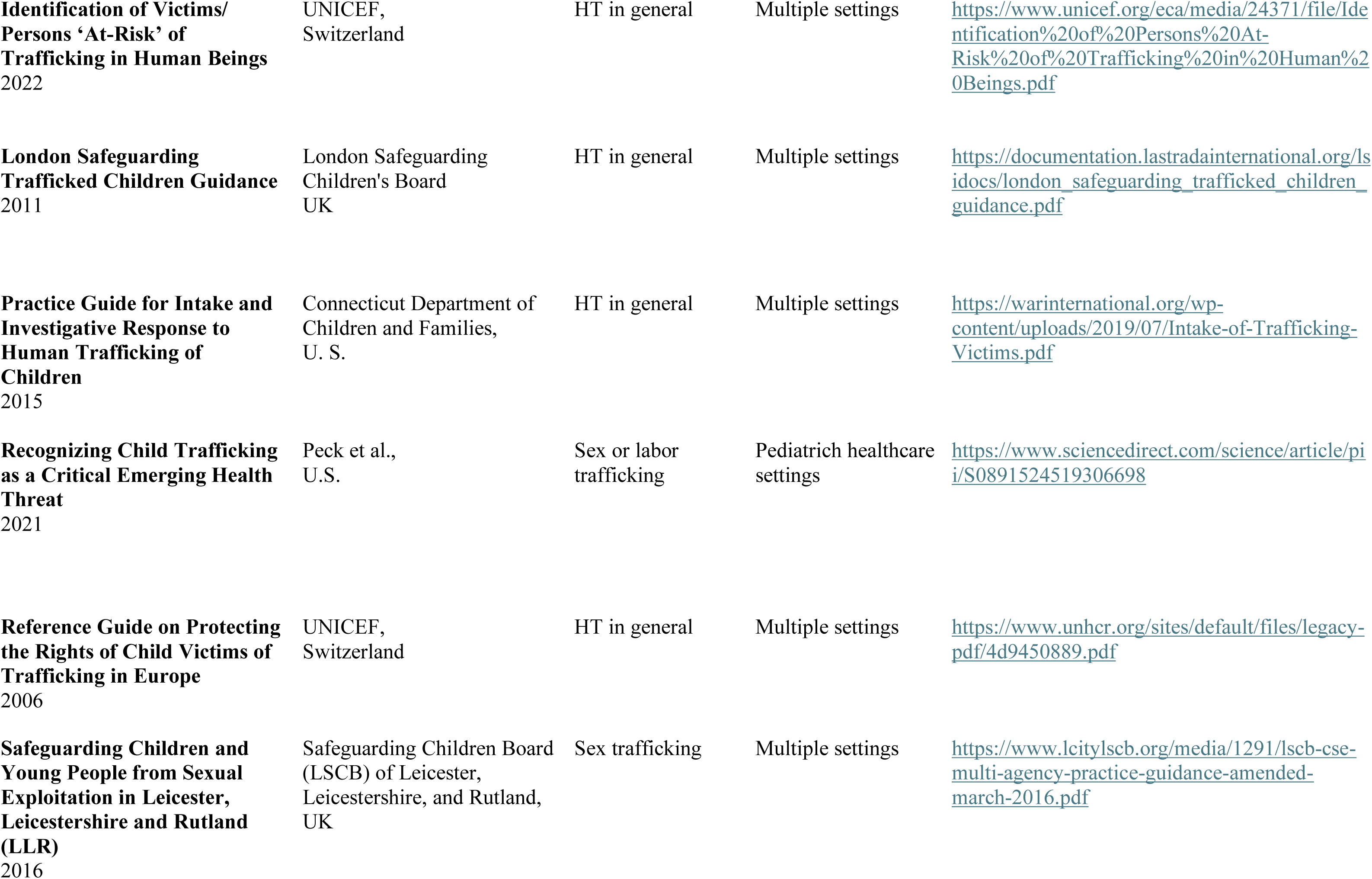
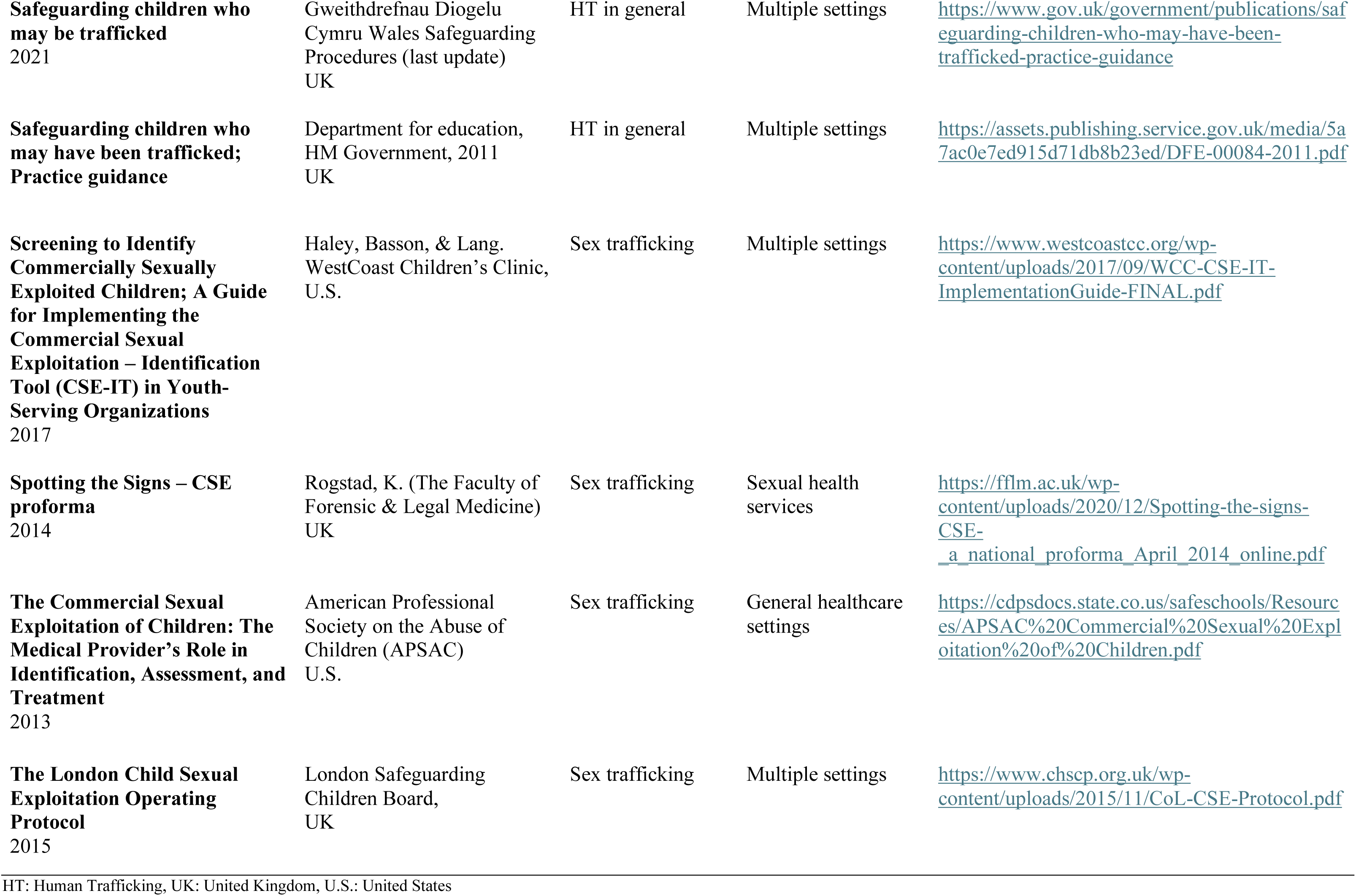
Toolkits and Guidelines for Identifying and Supporting Survivors of HT: Children and adolescents.

**Table 2.2.** Toolkits and Guidelines for Identifying and Supporting Survivors of HT: For Adults and All Ages.

| **Toolkit, guideline or protocol, year** | **Autor, Country** | **Detection focus** | **Application context** | **PMID or URL or DOI** |
| --- | --- | --- | --- | --- |
| **Adult Human Trafficking Screening Tool and Guide**<br>2018 | Macias-Konstantopoulos, W.<br>National Human Trafficking<br>Training and Technical<br>Assistance Center<br>(NHTTAC),<br>U.S. | HT in general | General healthcare<br>settings | https://acf.gov/sites/default/files/documents/otip/adult_human_trafficking_screening_tool_and_guide.pdf |
| **Assessing for Human Trafficking**<br>2021 | Human Trafficking<br>Collaborative University of<br>Michigan,<br>U. S. | HT in general | General healthcare<br>settings | https://humantrafficking.umich.edu/templates/assessing-for-human-trafficking/ |
| **Caring for Trafficked Persons**<br>2009 | International Organization for<br>Migration (IOM),<br>Switzerland | HT in general | General healthcare<br>settings | https://publications.iom.int/system/files/pdf/ct_h_andbook.pdf |
| **Committee on Health Care for Underserved Women**<br>2024 | Committee on Health Care<br>for Underserved Women<br>U.S. | HT in general | General healthcare<br>settings | https://www.acog.org/clinical/clinical-guidance/committee-opinion/articles/2019/09/human-trafficking |
| **Domestic Minor Sex Trafficking Intervene Identifying and Responding to America's Prostituted Youth,**<br>2010 | Shared Hope International,<br>U.S. | HT in general | Child Advocacy<br>Centers (CACs).<br>Social Services and<br>Mental Health<br>Settings.<br>Community<br>Organizations | https://www.thresholdglobalworks.com/pdfs/sex-trafficking-guide.pdf |
| **Framework for a Human Trafficking Protocol in Healthcare Settings,**<br>2010 | National Human Trafficking Resource Center (NHTRC)<br>U.S. | HT in general | General healthcare settings | https://humantraffickinghotline.org/en/resources/framework-human-trafficking-protocol-healthcare-settings#:~:text=This%20resource%20provides%20a%20framework%20for%20a%20human%20trafficking%20protocol#:~:text=This%20resource%20provides%20a%20framework%20for%20a%20human%20trafficking%20protocol |
| **Guia para desenvolver uma resposta às vítimas de tráfico de pessoas em ambientes de assistência à saúde**<br>2023 | HEAL Trafficking,<br>Brazil | HT in general | General healthcare settings | https://drive.google.com/file/d/1ZorgkJJwrW2R8iF3p74txOJg42HZDY-w/view |
| **Hiding in Plain Sight,** 2003 | Hughes, DM,<br>U.S. | Sex trafficking | General healthcare settings | https://www.academia.edu/35381082/Hiding_in_Plain_Sight_A_Practical_Guide_to_Identifying_Victims_of_Trafficking_in_the_U_S |
| **Human Trafficking: Care and Response** | Virginia Hospital & Healthcare Association /<br>Boyett, R.<br>U. S. | HT in general | General healthcare settings | https://vhha.com/reports/vhha-human-trafficking-resource-manual/ |
| **Human Trafficking: Guidebook on Identification, Assessment, and Response in the Health Care Setting** | Alpert, E. J.<br>U.S. | HT in general | General healthcare settings | https://addictiontraining.org/documents/resources/40_113_Human-Trafficking-Guidebook.pdf |
| **Human Trafficking: A Guide to Identification and Approach for the Emergency Physician,**<br>2016 | Shandro, J<br>U.S. | HT in general | Emergency Departments | https://pubmed.ncbi.nlm.nih.gov/27130802/ |
| **Human Trafficking: Clinical Assessment Guideline**, 2018 | Leslie, J<br>U.S. | Sex or labor<br>trafficking | Emergency<br>Departments | https://pubmed.ncbi.nlm.nih.gov/30216256/ |
| **Human Trafficking Response Program Shared Learnings Manual**<br>2019 | Dignity Health,<br>U.S. | HT in general | General healthcare<br>settings | https://www.dignityhealth.org/content/dam/dignity-health/pdfs/human-trafficking/dignity-health-htrp-sharedlearningsmanual-170512.pdf |
| **Human Trafficking Response Protocol. A Toolkit for Hospitals**<br>2020 | Michigan Department of<br>Health and Human Services-Division of Victim Services<br>U.S. | HT in general | Emergency<br>Departments | https://www.michigan.gov/mdhhs/-/media/Project/Websites/mdhhs/Division-of-Victim-Services/HTAB/HT-Toolkit/Human-Trafficking-Response-Protocol-Toolkit_Final.pdf?rev=97905539dca6465490b7a2d0b6943e7d |
| **Human Trafficking Toolkit for Healthcare Professionals** | United Against Human<br>Trafficking<br>U.S. | HT in general | General healthcare<br>settings | https://www.bcm.edu/sites/default/files/2021-03/hear-toolkit.pdf |
| **Identifying and Assisting Victims of Human Trafficking** | American Hospital<br>Association<br>U.S. | HT in general | General healthcare<br>settings | https://www.aha.org/identifying-and-assisting-victims-human-trafficking |
| **Identifying and supporting victims of modern slavery: guidance for health staff**<br>2015 | Department of Health &<br>Social Care<br>UK | HT in general | General healthcare<br>settings | https://www.gov.uk/government/publications/identifying-and-supporting-victims-of-human-trafficking-guidance-for-health-staff/identifying-and-supporting-victims-of-modern-slavery-guidance-for-health-staff |
| **Identifying Victims of Human Trafficking: What to Look for in a Healthcare Setting**<br>2016 | National Human Trafficking Resource Center<br>U.S. | HT in general | General healthcare settings | https://humantraffickinghotline.org/sites/default/files/What%20to%20Look%20for%20during%20a%20Medical%20Exam%20-%20FINAL%20-%202-16-16.pdf |
| **NHA Human Trafficking Toolkit** | Nebraska Hospital Association<br>U.S. | HT in general | General healthcare settings | https://www.nebraskahospitals.org/quality_and_safety/toolkits.html |
| **Protocol Toolkit for Developing a Response to Victims of Human Trafficking in Health Care Settings**<br>2017 | Baldwin., et al,<br>U.S. | HT in general | General healthcare settings | https://humantraffickinghotline.org/sites/default/files/HEAL%20Trafficking%20Protocol%20Toolkit%20secure%20%281%29.pdf |
| **Resource Guide on Human Trafficking for Title X Family Planning Clinics** | HHS Office of Population Affairs (OPA) Title X Family Planning Program<br>U.S. | HT in general | General healthcare settings | https://rhntc.org/sites/default/files/resources/opa_human_trafficking_resource_gde_2020-11-13.pdf |
| **Screening for Human Trafficking Guidelines for Administering the Trafficking Victim Identification Tool (TVIT)**<br>2014 | Vera Institute of Justice<br>U.S. | HT in general | Multiple settings | https://nij.ojp.gov/library/publications/screening-human-trafficking-guidelines-administering-trafficking-victim |
| **The Anatomy of Human Trafficking: Learning About the Blues: A Healthcare Provider's Guide**<br>2016 | Stevens, M,<br>U.S. | Sex or labor trafficking | General healthcare settings | https://journals.lww.com/forensicnursing/abstract/2016/06000/the_anatomy_of_human_trafficking_learning_about.2.aspx |
| **The IOM Handbook on Direct Assistance for Victims of Trafficking**<br>2007 | International Organization for Migration (IOM),<br>Switzerland | HT in general | Multiple settings | https://publications.iom.int/books/iom-handbook-direct-assistance-victims-trafficking-0 |
| **The PEARR Tool: Trauma-informed approach to victim assistance in health care settings.**<br>2019 | CommonSpirit Health, HEAL Trafficking, Pacific Survivor Center,<br>U.S. | HT in general | General healthcare settings | https://www.dignityhealth.org/content/dam/dignity-health/pdfs/pearrtoolm15nofield2019.pdf |
| **The Role of the Nurse in Combating Human Trafficking**<br>2011 | Sabella,<br>U.S. | HT in general | General healthcare settings | https://pubmed.ncbi.nlm.nih.gov/21270581/ |
| **Toolkit to Combat Trafficking in Persons**<br>2008 | United Nations Office on Drugs and Crime,<br>U.S. | HT in general | General healthcare settings | https://www.unodc.org/documents/human-trafficking/HT-toolkit-en.pdf |

HT: Human Trafficking, UK: United Kingdom, U.S.: United States

### Indicators of Human Trafficking

Various tools, guidelines, protocols, and professional training resources reviewed in this study include lists of indicators or red flags that may suggest a situation of human trafficking (HT). While widely used in clinical and service settings, these indicators are not, on their own, direct evidence of trafficking, nor do they function as formal diagnostic tools. However, their presence during patient care—especially when multiple indicators are observed—warrants further inquiry to explore the possibility that the individual may be a trafficking survivor. Indicators were identified across two toolkits (3,60), 18 protocol or guidance documents (64,65,67,68,71–75,82,83,85,87,88,90–92,95,97–99,101,102), two systematic reviews (45,103), three institutional reports or tools (20,31,104), and nine primary studies (43,49,105–111). Despite their broad application, there is no standardized classification or consensus on indicator prioritization across sources. Frequently reported signs include physical appearance-related features (e.g., hair loss, malnutrition, tattoos as branding), lack of identity documents, sexually transmitted infections (STIs), pregnancy, and uncooperative or fearful behavior. Among children and adolescents, the most commonly cited indicators involve physical abuse, malnutrition, and mental health symptoms such as anxiety or depression, whereas in adults, frequently reported signs include absence of documentation, distrust of authorities, and reproductive health concerns like STIs or unwanted pregnancies.

### Strategies to Improve Human Trafficking Identification in Healthcare Settings

Our review identified 23 studies or reports focused on educational strategies, the vast majority of which were conducted in the United States (n = 22). These included two systematic reviews (112,113) and two scoping review (52,114), along with three training programs (115–117) for which implementation data was not available. All educational strategies addressed key issues such as HT indicators, detection tools, and recommended steps following suspicion of a trafficking case. Many incorporated online components (n = 11) (106,115–124), presentations by interdisciplinary experts (n = 3) (120,125,126), case studies (n = 5) (118,124,127–129), and simulation-based training (n = 3) (121,128,130). Ten studies assessed pre- and post-intervention outcomes (118,123,127,129–131), with nearly all reporting improved knowledge (n = 10), confidence (n = 11), and awareness or attitudes (n = 7) toward HT. However, only three studies reported increases in actual case identification following training (127,130,132).

In addition to education, 24 studies explored other strategies for enhancing victim identification. Four were systematic or scoping reviews (53,103,133,134), and four examined the underuse of ICD-10-CM codes for trafficking in medical records (135–138). Seven studies evaluated the implementation of tools or protocols (e.g., PEARR, RAFT, Octavia, BCM A-HTP) (107,139–144), revealing both promising outcomes and persistent barriers such as limited use, time constraints, and low confidence in post-identification procedures. Notably, the Octavia software identified 184 high-risk cases in less than two years, vastly surpassing prior detection rates (140). Fourteen studies evaluated current practices, knowledge, and barriers among healthcare professionals (53,103,109–111,133,134,145–151). Two of these included interviews with trafficking survivors (145,146), exposing gaps in provider training, inconsistent screening practices, and the impact of stereotypes and privacy limitations. In both the U.S. and U.K., findings indicated that many professionals lacked formal protocols or sufficient awareness, often relying on personal judgment in the absence of structured tools.

Regarding the use of ICD-10 codes, studies highlighted their significant underutilization in clinical documentation. For example, Garg et al., 2022 (137) found trafficking-related codes in only 0.005% of pediatric patient encounters, and Kerr & Bryant, 2022 (135) identified just 298 cases among nearly 70 million patients. Common barriers include unfamiliarity with code application and poor integration into electronic medical records. Furthermore, patients identified through ICD-10 codes frequently exhibited mental health comorbidities (e.g., depression, PTSD, substance use), with healthcare costs for survivors averaging over $31,000 USD annually, nearly five times the national average (138).

Across all strategies reviewed, the most commonly proposed improvements include mandatory training in HT identification and trauma-informed care, structured use of validated screening tools, improved electronic documentation systems, interdisciplinary collaboration with external support networks, and enhanced referral and follow-up protocols (Appendix 2).

## Discussion

This scoping review provides a comprehensive synthesis of a wide range of resources designed to support professionals in identifying and responding to human trafficking. These resources vary significantly in format, audience, and scope. They encompass structured tools such as questionnaires and indicator checklists, as well as broader guidance documents, including toolkits, protocols, and guidelines that inform identification, care, and case management.

Detection tools were predominantly developed in the United States, with limited contributions from other regions. Their design and focus often vary according to the intended population. Tools targeting multiple age groups usually address HT in general, covering both labor and sexual exploitation. Conversely, tools specifically created for children and adolescents tend to focus more narrowly on sexual exploitation. Labor trafficking and other forms of exploitation are notably underrepresented across the tools identified.

A recurring challenge for healthcare professionals is selecting the most appropriate tool for their context. This difficulty stems from the proliferation of tools, inconsistent psychometric evaluation, and the absence of a reference standard for validation. It is essential that tools truly measure the intended construct, and a tool validated in one setting or population may not retain its validity in another. Validation is therefore crucial when adapting instruments to new contexts to ensure accurate measurement and sound decision-making(152,153). While some tools like the Greenbaum 6-question (39) screen offer practicality, their limited validation and scope restrict utility. Tools like the CSE-IT (38) and TVIT (42) offer more reliable options; however, their complexity and resource demands may hinder routine implementation. One-question screeners present a simple alternative but may lack sufficient depth for comprehensive identification.

Tool diversity—ranging from standardized questionnaires to key-question checklists—underscores the need for greater standardization and comparative assessment to inform best practices. Healthcare systems should prioritize both the validation and the feasibility of tools, ensuring they are adaptable to clinical realities and resource constraints.

While toolkits and guidelines share similarities, their functions differ. Toolkits often include structured instruments and decision-support materials designed for case identification, whereas guidelines tend to provide overarching frameworks, protocols, and systematic recommendations based on normative references or evidence syntheses. Seven comprehensive toolkits were identified that integrate screening tools, referral pathways, and trauma-informed principles (3,60–63,73,102). Additionally, a wide array of protocols and guidelines—mostly from the U.S. but also from international organizations like International Organization for Migration (IOM) and United Nations Children’s Fund (UNICEF)—target various populations and settings.

These documents vary in format and detail but are primarily directed at healthcare providers. Some also extend to law enforcement and frontline service personnel, reflecting the multisectoral nature of HT response. Many guidelines incorporate clinical indicators, trauma-informed practices, multidisciplinary coordination, legal protections, and referral mechanisms. Importantly, no clinical practice guidelines on HT met rigorous methodological standards involving systematic reviews or stakeholder engagement, representing a critical gap. The lack of high-quality, evidence-based clinical guidance on this subject limit healthcare providers’ ability to act effectively. In addition, many documents lack clarity regarding specific target users (e.g., emergency physicians) and implementation contexts (e.g., outpatient care). Recommendations are not always operationalized with tools like flowcharts or checklists, and monitoring criteria are often absent.

HT indicators, while widely used, are not predictive on their own. They serve as clinical flags that prompt further inquiry but should not be treated as diagnostic tools. Indicators are inconsistently categorized and described across documents, highlighting the need for standardization in their use and presentation.

Educational strategies aimed at raising awareness, improving knowledge, and fostering confidence among healthcare professionals have shown generally positive results. Webinars, simulations, case studies, and interprofessional training sessions frequently improved participants’ knowledge and preparedness. However, improvements were not always sustained, and only one study demonstrated a measurable increase in case identification. To be impactful, training programs must be continuous and integrated with institutional policies, tools, and support systems.

Implementation studies revealed that integrating screening tools into clinical workflows—including the use of trauma-informed models, AI-based alerts, and institutional protocols—can improve identification. Nonetheless, systemic challenges such as limited time, privacy constraints, and provider discomfort persist. Biases and lack of awareness particularly affect identification among high-risk populations.

To address these issues, mandatory and structured use of validated tools is recommended, along with standardized training embedded in medical education. Interdisciplinary collaboration and strong post-identification response protocols are also essential for improving detection and ensuring coordinated support for survivors.

Additionally, it is crucial to involve community and patient partners with lived experience in both the development and evaluation of interventions. Their participation not only enhances the contextual relevance of tools but also ensures that ethical and culturally competent approaches are applied. Future research and implementation strategies should also acknowledge the sociopolitical dimensions of trafficking, particularly in how definitions and frameworks can lead to false-positive identifications. Accurate detection will enhance data collection, which is essential for better understanding the prevalence and scope of human trafficking with greater precision.

The trafficking discourse also needs to consider the spectrum of exploitation and the need to access recourses throughout the spectrum. These strategies collectively offer a pathway towards a more effective and equitable healthcare response to human trafficking.

### Limitations and Strengths

This study presents several limitations. First, the lack of standardization in the terminology used across different reports regarding key concepts related to HT (e.g., the overlap between the terms sexual exploitation and sex trafficking in some reports) makes it challenging to categorize the evidence. Second, while a vast body of systematic reviews addresses screening tools for HT, this study, despite offering insights into the broader issue, highlights the need for decision-support tools that help interest-holders determine the most appropriate screening tool based on their profession, context, setting, and target population. Tools such as evidence maps or decision aids for professionals could significantly enhance the real-world impact of screening practices in healthcare settings, where early detection of this critical condition is essential.

Despite these limitations, this study also presents notable strengths. First, given the profound impact of HT on individuals and society—including its economic and political implications—this issue remains a research priority where collaborative efforts are crucial to advancing prevention, identification, and intervention strategies. The systematic methods, including the comprehensive and updated literature search, contribute to understanding the current landscape of screening tools for HT in clinical settings, their psychometric properties, validation, and implementation efforts.

Additionally, a significant effort was made to synthesize and summarize the extensive number of HT indicators compiled over the years by different researchers and organizations.

### Future Directions

This study provides insights into the necessary next steps to enhance the detection of HT in clinical settings. First, the development of user-friendly tools: There is an urgent need for decision-support tools that assist healthcare professionals in selecting the most appropriate screening tool based on their specific needs and context. Second, the development of evidence-based clinical guidelines is imperative to establish clear and systematic recommendations for HT screening, ensuring adoption and adaptation to resource-limited settings. Lastly, harmonization and collaboration will be instrumental. Efforts should be coordinated and collaborative, minimizing redundancy and research waste in evidence synthesis. Research priorities should focus on validating existing screening tools and implementation considerations to ensure their effectiveness in real-world settings.

## Conclusions

It is essential to improve and unify the tools used to identify and address HT, as no universal standard or consensus on definitions currently exists. Available tools largely focus on sexual exploitation among minors, while labor trafficking and other forms receive less attention, creating gaps in detection and response. Continuous training increases the confidence and awareness of professionals but requires regular reinforcement, practical tools, and interdisciplinary collaboration to sustain its impact. Systemic barriers persist, including time constraints, privacy concerns, and discomfort in addressing trafficking in healthcare settings, particularly in emergency services. To overcome these challenges, it is necessary to integrate clear detection protocols, strengthen training, and promote coordination between healthcare, social services, and survivor support organizations. A comprehensive approach, with validated tools and institutional commitment, will enable more effective identification and dignified, survivor-centered care for those affected by trafficking.

## Data Availability

All data produced in the present study are available upon reasonable request to the authors

