## Appendix 1 for "Human Trafficking Detection in Health Care Settings: A Scoping Review"

#### Contend

### Search Strategy Resources

This annex compiles all data portals, reports, and manually identified resources related to human trafficking and modern slavery as used in the search strategy.

##### Search Strategy-21 JUN 2024

| | **Medline via PUBMED** | **HITS** |
|---|---|---|
| **Baseline** | "human trafficking"[MeSH Terms] OR "human traffic*"[Title/Abstract] OR "sex traffic*"[Title/Abstract] OR "sexual traffic*"[Title/Abstract] OR "sex exploitat*"[Title/Abstract] OR "sexual exploitat*"[Title/Abstract] OR "organ traffic*"[Title/Abstract] OR "transplant tourism"[Title/Abstract] OR "child* traffic*"[Title/Abstract] OR "modern slavery"[Title/Abstract] OR "labor traffic*"[Title/Abstract] OR "forced labor"[Title/Abstract] OR "kidnap*"[Title/Abstract] | 2,672 |
| | **COMBINACIONES** | |
| Reviews No time limit | ("human trafficking"[MeSH Terms] OR "human traffic*"[Title/Abstract] OR "sex traffic*"[Title/Abstract] OR "sexual traffic*"[Title/Abstract] OR "sex exploitat*"[Title/Abstract] OR "sexual exploitat*"[Title/Abstract] OR "organ traffic*"[Title/Abstract] OR "transplant tourism"[Title/Abstract] OR "child* traffic*"[Title/Abstract] OR "modern slavery"[Title/Abstract] OR "forced labor"[Title/Abstract] OR "labor traffic*"[Title/Abstract] OR "kidnap*"[Title/Abstract]) AND *("review"[Publication Type] OR "systematic review"[Filter] OR "review"[Title])* | 398 |
| | ("human trafficking"[MeSH Terms] OR "human traffic*"[Title/Abstract] OR "sex traffic*"[Title/Abstract] OR "sexual traffic*"[Title/Abstract] OR "sex | |

| | | |
|---|---|---|
| Other designs and reports<br><br>No time limit | exploitat*"[Title/Abstract] OR "sexual exploitat*"[Title/Abstract] OR "organ traffic*"[Title/Abstract] OR "transplant tourism"[Title/Abstract] OR "child* traffic*"[Title/Abstract] OR "modern slavery"[Title/Abstract] OR "forced labor"[Title/Abstract] OR "labor traffic*"[Title/Abstract] OR "kidnap*"[Title/Abstract]) AND *("clinical trial"[Publication Type] OR "meta analysis"[Publication Type] OR "randomized controlled trial"[Publication Type] OR "trial"[Title] OR "qualitative"[Title] OR "observational"[Title] OR "case"[Title] OR "cohort"[Title] OR "stud*"[Title] OR "analys*"[Title] OR "global"[Title] OR "countr*"[Title] OR "region*"[Title] OR "nation*"[Title] OR "internat*"[Title] OR "report"[Title] OR "brief"[Title] OR "polic*"[Title] OR "who"[Title] OR "organizat*"[Title])* | 635 |
| | **BVSalud (BIREME-LILACS-WHOIris-AIM (Africa)** | **HITS** |
| | ("human trafficking") OR ("sex trafficking") OR ("organ trafficking") OR ("labor trafficking") OR ("child trafficking") AND ( db:("LILACS" OR "INDEXPSI" OR "IBECS" OR "LIPECS" OR "MINSAPERU" OR "WHOLIS" OR "colecionaSUS" OR "AIM" OR "SES-SP" OR "BIGG" OR "PIE" OR "SMS-SP")) | **88** |
| | **EMBASE** | **HITS** |

History | Save | Delete | Print view | Export | Email | Combine > using And Or | Collapse

| | | |
|---|---|---|
| **#15** | #12 OR #13 | 803 |
| **#14** | #12 OR #13 | 803 |
| **#13** | #11 AND (**'article'**/it OR **'review'**/it) | 426 |
| **#12** | #11 AND (**'case report'**/de OR **'case study'**/de OR **'clinical article'**/de OR **'clinical study'**/de OR **'clinical trial'**/de OR **'cohort analysis'**/de OR **'comparative study'**/de OR **'controlled study'**/de OR **'cross sectional study'**/de OR **'evidence based practice'**/de OR **'evidence based practice center'**/de OR **'exploratory research'**/de OR **'grounded theory'**/de OR **'intervention study'**/de OR **'interview'**/de OR **'major clinical study'**/de OR **'meta analysis'**/de OR **'model'**/de OR **'multicenter study'**/de OR **'observational study'**/de OR **'participatory research'**/de OR **'pilot study'**/de OR **'practice guideline'**/de OR **'prospective study'**/de OR **'qualitative research'**/de OR **'questionnaire'**/de OR **'randomized controlled trial'**/de OR **'retrospective study'**/de OR **'semi structured interview'**/de OR **'structured interview'**/de OR **'structured questionnaire'**/de OR **'systematic review'**/de) | 552 |
| **#11** | #10 AND [embase]/lim NOT ([embase]/lim AND [medline]/lim) | 1,107 |
| **#10** | #1 OR #2 OR #3 OR #4 OR #5 OR #6 OR #7 OR #8 OR #9 | 3,968 |
| **#9** | **'labor trafficking'** | 82 |
| **#8** | **'modern slavery'** | 69 |
| **#7** | **'slavery'**/exp OR **'slavery'** | 1,082 |
| **#6** | **'organ trafficking'**/exp OR **'organ trafficking'** | 491 |
| **#5** | **'child sexual exploitation'**/exp OR **'child sexual exploitation'** | 267 |
| **#4** | **'sexual exploitation'**/exp OR **'sexual exploitation'** | 1,273 |
| **#3** | **'child trafficking'**/exp OR **'child trafficking'** | 127 |
| **#2** | **'sex trafficking'**/exp OR **'sex trafficking'** | 655 |
| **#1** | **'human trafficking'**/exp OR **'human trafficking'** | 1,538 |

Duplicates between #12 and #13 resolved using Zotero. Exported results: 748 records.

**EXPORTED TO RAYYAN**
**143 duplicates resolved, total for screening: 1,753.**

#### 10 AUG 2024 – Update

**To retrieve relevant articles excluded by the publication filter**

| | |
|---|---|
| (("human trafficking"[MeSH Terms] OR "human traffic*"[Title/Abstract] OR "sex traffic*"[Title/Abstract] OR "sexual traffic*"[Title/Abstract] OR "sex exploitat*"[Title/Abstract] OR "sexual exploitat*"[Title/Abstract] OR "organ traffic*"[Title/Abstract] OR "transplant tourism"[Title/Abstract] OR "child* traffic*"[Title/Abstract] OR "modern slavery"[Title/Abstract] OR "forced labor"[Title/Abstract] OR "labor traffic*"[Title/Abstract] OR "kidnap*"[Title/Abstract]) **NOT** ((("human trafficking"[MeSH Terms] OR "human traffic*"[Title/Abstract] OR "sex traffic*"[Title/Abstract] OR "sexual traffic*"[Title/Abstract] OR "sex exploitat*"[Title/Abstract] OR "sexual exploitat*"[Title/Abstract] OR "organ traffic*"[Title/Abstract] OR "transplant tourism"[Title/Abstract] OR "child* traffic*"[Title/Abstract] OR "modern slavery"[Title/Abstract] OR "forced labor"[Title/Abstract] OR "labor traffic*"[Title/Abstract] OR "kidnap*"[Title/Abstract]) AND ("clinical trial"[Publication Type] OR "meta-analysis"[Publication Type] OR "randomized controlled trial"[Publication Type] OR "trial"[Title] OR "qualitative"[Title] OR "observational"[Title] OR "case"[Title] OR "cohort"[Title] OR "stud*"[Title] OR "analys*"[Title] OR "global"[Title] OR "countr*"[Title] OR "region*"[Title] OR "nation*"[Title] OR "internat*"[Title] OR "report"[Title] OR "brief"[Title] OR "polic*"[Title] OR "who"[Title] OR "organizat*"[Title])) OR (("human trafficking"[MeSH Terms] OR "human traffic*"[Title/Abstract] OR "sex traffic*"[Title/Abstract] OR "sexual traffic*"[Title/Abstract] OR "sex exploitat*"[Title/Abstract] OR "sexual exploitat*"[Title/Abstract] OR "organ traffic*"[Title/Abstract] OR "transplant tourism"[Title/Abstract] OR "child* traffic*"[Title/Abstract] OR "modern slavery"[Title/Abstract] OR "forced labor"[Title/Abstract] OR "labor traffic*"[Title/Abstract] OR "kidnap*"[Title/Abstract]) AND ("review"[Publication Type] OR "systematic review"[Filter] OR "review"[Title])))) AND *("clinical trial"[Publication Type] OR "meta-analysis"[Publication Type] OR "randomized controlled trial"[Publication Type] OR "Observational Study"[Publication Type] OR "Comparative Study"[Publication Type] OR "Evaluation Study"[Publication Type] OR "Surveys and Questionnaires"[MeSH Terms] OR "Cohort Studies"[MeSH Terms] OR "stud*"[Title] OR "trial"[Title] OR "qualitative"[Title] OR "case"[Title] OR "cohort"[Title] OR "meta-analysis"[Title/Abstract] OR "metanalysis"[Title/Abstract] OR "rct"[Title/Abstract] OR "controlled"[Title/Abstract] OR "Observational Study"[Title/Abstract] OR "Comparative Study"[Title/Abstract] OR "Evaluation Study"[Title/Abstract] OR "Cross-sectional"[Title/Abstract] OR "survey*"[Title/Abstract] OR "questionnair*"[Title/Abstract] OR "Follow-Up"[Title/Abstract] OR "Longitudinal"[Title/Abstract] OR "Prospective"[Title/Abstract] OR "Retrospective"[Title/Abstract])* | **Baseline block**<br><br>**Search exported**<br><br>**21 JUN**<br><br>More sensitive publication filter |
| **TOTAL** | **450** |

Duplicates identified. Rayyan re-flagged previously reviewed duplicates. You can see them already labeled as included or not. Only new duplicates were resolved.
A total of 22 duplicates with Embase had already been reviewed.

References uploaded – Pubmed Act_451r_12AUG2024

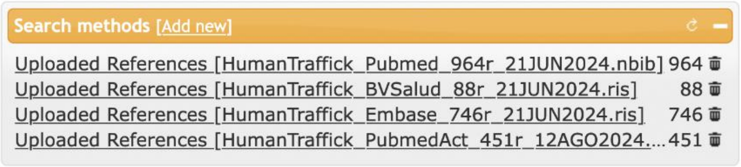

#### Data Portals Searched – 01 JUL 2024

**United Nations and Global Agencies**

- **UN Network on Migration**
  https://migrationnetwork.un.org/

- **UN ICAT – Inter-Agency Coordination Group against Trafficking in Persons**
  https://icat.un.org/

- **UNHCR Operational Data Portal**
  https://data.unhcr.org/

- **UNODC – Human Trafficking Research Portfolio**
  https://www.urban.org/policy-centers/justice-policy-center/projects/human-trafficking-research-portfolio

- **UNODC SHERLOC Databases**
  https://sherloc.unodc.org/cld/en/st/home.html

- **ILO Forced Labor Resources**
  https://libguides.ilo.org/c.php?g=259896&p=2698609

**International Organization for Migration (IOM)**

- **Global Migration Data Portal (GMDAC)**
  https://www.migrationdataportal.org/
  - Thematic Data Overviews (38)
  - Regional Data Overviews (14):
    - Africa: Eastern, Middle, Northern, Southern, Western
    - Americas *(section under construction)*
    - Asia: Central, South-eastern, Southern, Western
    - Europe, Oceania
- **UN Migration Platform**
  https://migrantprotection.iom.int/en

- **Global Compact for Migration (GCM)**
  https://www.iom.int/global-compact-migration

**Counter Trafficking Data Collaborative (CTDC)**

- **VCMS and HTCDS Toolkit**
  https://www.ctdatacollaborative.org/page/human-trafficking-case-data-standards-toolkit-and-guidance-htcds

- **Global Victim–Perpetrator Synthetic Dataset**
  https://www.ctdatacollaborative.org/global-victim-perpetrator-synthetic-dataset

- **Global Data Hub and Dashboards**
  https://www.ctdatacollaborative.org/page/dashboards-datasets

**Governmental and Independent Sources**

- **US Department of State – Office to Monitor and Combat Trafficking in Persons**
  https://www.state.gov/.../office-to-monitor-and-combat-trafficking-in-persons/

- **US Department of Justice – Human Trafficking Prosecution Unit**
  https://www.justice.gov/crt/human-trafficking-prosecution-unit-htpu

- **USAID – Countering Trafficking in Persons**
  https://www.usaid.gov/trafficking

- **Walk Free – Global Data on Modern Slavery (2023)**
  https://www.walkfree.org/

- **Alberta Anti-Human Trafficking Data Portal**
  https://www.endhtalberta.ca/

**REPORTS – IN PROGRESS**

**2024**

- **US Department of State – Trafficking in Persons Report**
  https://www.state.gov/reports/2024-trafficking-in-persons-report/

**2023**

- **IOM – Making Each Case Count: Leveraging Administrative Data on Trafficking in Persons**
  https://publications.iom.int/books/making-each-case-count-leveraging-administrative-data-trafficking-persons

- **UNDP – Human Development Report 2023–24**
  https://data.unhcr.org/en/documents/details/108475

- **Walk Free – Global Slavery Index 2023**
  https://cdn.walkfree.org/content/uploads/2023/05/17114737/Global-Slavery-Index-2023.pdf

- **UN SSE & FAST – Finance Against Slavery & Trafficking Initiative**
  https://cdn.walkfree.org/content/uploads/2023/11/30123753/SSE-FAST-WalkFree-Modern-Slavery-2023.pdf

**2022**

- **UNODC – Global Report on Trafficking in Persons**
  https://www.unodc.org/unodc/en/data-and-analysis/glotip.html
  - *Country Profiles*:
    - Central and South-Eastern Europe
    - East Asia and the Pacific
    - Eastern Europe and Central Asia
    - North Africa and the Middle East
    - North America, Central America and the Caribbean
    - South America
    - South Asia
    - Sub-Saharan Africa
    - Western and Southern Europe
- **ILO – Global Estimates of Modern Slavery: Forced Labour and Forced Marriage**
  https://www.ilo.org/media/370821/download

- **ICAT – Annual Report**
  https://icat.un.org/sites/g/files/tmzbdl461/files/publications/icat_2022_co-chairs_annual_report_6.pdf

- **Urban Institute (USA)**
  - *Recommendations for Practitioners in Antitrafficking Task Forces*
    https://www.urban.org/research/publication/recommendations-practitioners-engaged-antitrafficking-task-forces
  - *Evaluation of the Enhanced Collaborative Model Task Forces*
    https://www.urban.org/research/publication/findings-evaluation-enhanced-collaborative-model-task-forces-combat-human-trafficking
  - *Collaboration and Challenges in Antitrafficking Task Forces*
    https://www.urban.org/research/publication/collaboration-and-challenges-antitrafficking-task-forces

- **Walk Free – Global Estimates of Modern Slavery: Forced Labour and Forced Marriage**
  https://cdn.walkfree.org/content/uploads/2022/09/12142341/GEMS-2022_Report_EN_V8.pdf

- **Walk Free – Modern Slavery Response & Remedy Framework**
  https://cdn.walkfree.org/content/uploads/2022/07/12132831/Walk-Free-Response-and-Remedy-Framework-1.pdf

**2021**

- **UNODC Toolkit – Human Rights and Gender Equality in TIP Interventions**
  https://www.unodc.org/documents/human-trafficking/GLO-ACTII/UNODC_Toolkit_for_mainstreaming_Human_Rights_and_Gender_Equality_February_2021.pdf

- **ICAT – 20th Anniversary of the TIP Protocol: Analytical Review**
  https://icat.un.org/sites/g/files/tmzbdl461/files/publications/icat_analytical_paper_2020_final_0.pdf

- **ICAT – Issue Brief 11: Organ Removal**
  https://icat.un.org/sites/g/files/tmzbdl461/files/publications/icat_brief_tip_for_or_final.pdf

- **ICAT – Issue Brief 10: Public Procurement**
  https://icat.un.org/sites/g/files/tmzbdl461/files/publications/icat_issue_brief_10_on_public_procurement_0.pdf

**2020**

- **ILO – World Migration Report 2020**
  https://publications.iom.int/books/world-migration-report-2020

- **UNODC – Legislative Guide for TIP Protocol**
  https://www.unodc.org/documents/human-trafficking/2020/TiP_LegislativGuide_Final.pdf

- **UNODC – Model Legislative Provisions on TIP**
  https://www.unodc.org/documents/human-trafficking/2020/TiP_ModelLegislativeProvisions_Final.pdf

- **UNODC – Interlinkages between TIP and Marriage**
  https://www.unodc.org/documents/human-trafficking/2020/UNODC_Interlinkages_Trafficking_in_Persons_and_Marriage.pdf

**Earlier Reports**

- **2019 – ILO: Ending Child Labour and Human Trafficking in Global Supply Chains**
  https://www.ilo.org/media/405766/download

- **2017 – ICAT: TIP in Humanitarian Crises**
  https://icat.un.org/sites/g/files/tmzbdl461/files/publications/icat-ib-02-final.pdf

- **2017 – ILO: Regional Brief for the Americas on Modern Slavery and Child Labour**
  https://labordoc.ilo.org/permalink/41ILO_INST/j3q9on/alma995073593002676

- **2016 – IOM: Prevalence Indication Survey**
  https://data.unhcr.org/en/documents/details/47217

- **2015 – UN Migration Network: Human Rights of Vulnerable Populations in the Inter-American System**
  https://migrationnetwork.un.org/resources/human-rights-migrants-refugees-stateless-persons-victims-human-trafficking-and-internally

- **2015 – UN Migration Network: Addressing TIP in Times of Crisis**
  https://migrationnetwork.un.org/resources/addressing-human-trafficking-and-exploitation-times-crisis-evidence-and-recommendations

- **2014 – UNHCR: Gender Traffic Lights**
  https://data.unhcr.org/en/documents/details/46165

- **2010 – UN Migration Network: Principles and Guidelines on Human Rights and TIP**
  https://migrationnetwork.un.org/resources/recommended-principles-and-guidelines-human-rights-and-human-trafficking

#### Search Update – 18 JAN 2025

*(Resources identified manually through reference tracking and additional grey literature searches.)*

**Protocols and Toolkits**

- **Human Trafficking Response Protocol: A Toolkit for Hospitals** (Michigan, 2021)
  https://www.michigan.gov/mdhhs/.../Human-Trafficking-Response-Protocol-Toolkit_Final.pdf

- **Human Trafficking of Children Protocol** (Michigan, 2017)
  https://www.michigan.gov/mdhhs/.../HumanTraffickingProtocol.pdf

- **HUMAN TRAFFICKING SCREENING TOOL – ONGOING CASES**
  Michigan Department of Health and Human Services
  View Form

- **Practice Guide: Intake and Investigative Response to Human Trafficking of Children**
  Connecticut Department of Children and Families (2015)
  https://portal.ct.gov/.../humantraffickingbpg.pdf

**Screening Tools and Assessment Instruments**

- **National Human Trafficking Assessment Tool**
  Canadian Council for Refugees
  https://ccrweb.ca/en/national-human-trafficking-assessment-tool

- **Labor and Human Trafficking in Nebraska – NHA**
  https://www.nebraskahospitals.org/file_download/...

- **Labor Trafficking Self-Assessment Card – USER GUIDE**
  The Advocates for Human Rights
  https://www.theadvocatesforhumanrights.org/res/byid/8376

- **Adult Human Trafficking Screening Tool and Guide** (OTIP, 2018)
  - English Version
  - Spanish Version

**Reference Frameworks and Guides**

- **Framework for a Human Trafficking Protocol in Healthcare Settings** (NHTRC, 2010)
  https://humantraffickinghotline.org/.../framework-human-trafficking-protocol-healthcare-settings

- **Hiding in Plain Sight: A Practical Guide to Identifying Victims of Trafficking in the U.S.** (2003)
  https://www.academia.edu/...

- **Potential Trafficking Indicators | Polaris Project**
  https://ago.mo.gov/.../potential-trafficking-indicators.pdf

**Case Studies and Implementation Materials**

- **Covenant House NY – Youth Experience Report** (2013)
  Homelessness, Survival Sex and Human Trafficking

- **SHYIP Protocol Guidelines – Ramsey County, MN** (2009)
  https://www.ramseycounty.us/.../SHYIP_guidelines_feb_2010.pdf

- **Building a Child Welfare Response to Child Trafficking Handbook** (Loyola Univ. Chicago & IOFA, 2011)
  https://ecommons.luc.edu/.../chrc

- **Dignity Health – Human Trafficking Response Program Manual** (2019)
  https://www.dignityhealth.org/.../dignity-health-htrp-sharedlearningsmanual-170512.pdf

**UK-Specific Resources**

- **The London Child Sexual Exploitation Operating Protocol (2nd ed., 2015)**
  https://dera.ioe.ac.uk/id/eprint/23041/1/Satellite.pdf

- **ILO & EU Commission – Operational Indicators of TIP**
  https://www.ilo.org/sites/.../wcms_105023.pdf
- **Kent and Medway – CSE Identification, Assessment & Planning Tools** (2020)
  https://view.officeapps.live.com/...
- **Kent & Medway Multi-agency Procedures**
  https://www.proceduresonline.com/kentandmedway
- **Support Level Guidance – Kent**
  https://www.kscb.org.uk/.../SLG-sheet-v13.pdf
- **Bedfordshire Child Exploitation Tool (2023?)**
  https://www.bavex.co.uk/.../Beds-CE-tool-Update23.pdf
- **UK Child Exploitation Disruption Toolkit**
  https://assets.publishing.service.gov.uk/.../Child_Exploitation_Disruption_Toolkit_082022.pdf
- **LLR Multi-Agency CSE Practice Guidance (2015)**
  https://www.lcitylscb.org/media/1291/lscb-cse-multi-agency-practice-guidance-amended-march-2016.pdf

#### Data Portals – 18 JAN 2025

**Key International Reports and Explainers**

- **UNODC – Global Report on Trafficking in Persons 2024**
  https://www.unodc.org/documents/data-and-analysis/glotip/2024/GLOTIP2024_BOOK.pdf
- **UNODC – Explainer: Understanding Human Trafficking for Organ Removal**
  https://www.unodc.org/unodc/en/frontpage/2024/June/explainer_-understanding-human-trafficking-for-organ-removal.html
- **US Department of State – TIP Report: United States 2024**
  https://www.state.gov/reports/2024-trafficking-in-persons-report/united-states/
- **US Bureau of Justice Statistics – Human Trafficking Data Collection Activities 2024**
  https://bjs.ojp.gov/library/publications/human-trafficking-data-collection-activities-2024

**Healthcare and Training Resources**

- **IOM – Human Trafficking for Healthcare Providers (E-Course)**
  https://migrantprotection.iom.int/en/learning/e-courses/human-trafficking-healthcare-providers

- **HEAL Trafficking – 2024 Impact Report**
  https://healtrafficking.org/wp-content/uploads/2024/02/2024-TIP-Report_HMS.pdf

- **American Health Association – Combating Human Trafficking**
  https://www.aha.org/combating-human-trafficking

- **Trafficking in Persons Reference List – Forensic Nursing Network Inc**
  https://www.otterbein.edu/alumni/wp-content/uploads/sites/4/2024/03/Human-Trafficking-Resources-for-Healthcare-2024.pdf

- **Polaris – A Roadmap for Systems and Industries to Prevent and Disrupt Human Trafficking**
  https://polarisproject.org/wp-content/uploads/2018/08/A-Roadmap-for-Systems-and-Industries-to-Prevent-and-Disrupt-Human-Trafficking-Health-Care.pdf

- **Polaris – The Typology of Modern Slavery**
  https://polarisproject.org/the-typology-of-modern-slavery/

- **Children's Hospitals Association – Addressing Child Trafficking in Hospitals**
  https://www.childrenshospitals.org/education/events/addressing-child-trafficking-in-childrens-hospitals

- **HEAL Trafficking Course – Texas (2024)**
  https://www.medbridge.com/educate/courses/human-trafficking-for-healthcare-professionals-texas-2024-hanni-stoklosa-Dec22-1-Dec23-1

**Legislation and Policy Resources**

- **SB 963 – California Hospitals Self-Identification Procedure**
  - US Digital Democracy:
    https://calmatters.digitaldemocracy.org/bills/ca_202320240sb963
  - California Legislative Info:
    https://leginfo.legislature.ca.gov/faces/billTextClient.xhtml?bill_id=202320240SB963

- **Texas HHS – Required Signage for Hospitals and Abortion Facilities**
  https://www.hhs.texas.gov/sites/default/files/documents/doing-business-with-hhs/provider-portal/facilities-regulation/human-trafficking-hb-2552.pdf

**Institutional Statements and Programs**

- **CareSource – Human Trafficking (2024 IHCP Seminar)**
  https://www.in.gov/medicaid/providers/files/IHCP-Works-2024-CareSource-Human-Trafficking.pdf
- **NHS Bradford Teaching Hospitals – Modern Slavery Statement 2024–25**
  https://www.bradfordhospitals.nhs.uk/wp-content/uploads/2024/11/BTHFT-Slavery-and-Human-Trafficking-Statement-24-25-Approved.pdf
- **HCA Healthcare – Awareness and Prevention Program**
  https://hcahealthcaretoday.com/2022/07/29/we-have-a-duty-to-respond-hca-healthcares-human-trafficking-awareness-and-prevention-program/

**Professional Guidance and Toolkits**

- **AMA Journal of Ethics – Health Care Organizations and Human Trafficking**
  https://journalofethics.ama-assn.org/article/how-should-health-care-organizations-limit-roles-human-trafficking-their-labor-and-supply-chains/2024-04
- **Preble Street – Victims Going Undetected in Healthcare**
  https://www.preblestreet.org/2023/01/26/trafficking-healthcare/
- **The Doctors Company – Identifying Victims in Clinical Settings**
  https://www.thedoctors.com/articles/is-your-patient-a-victim-of-human-trafficking/
- **Missouri Hospital Association – Human Trafficking Toolkit**
  https://web.mhanet.com/media-library/human-trafficking-toolkit/
- **County of Marin EMS – Sexual Assault/Human Trafficking Protocol (2024)**
  https://ems.marinhhs.org/sites/default/files/policy_procedure/GPC%2010-%20Sexual%20Assault-Human%20trafficking%202024.pdf
- **American Nurse – Patient as Trafficking Victim**
  https://www.nationwidechildrens.org/...
- **Ohio State University – What Providers Need to Know**
  https://ccme.osu.edu/storage/Webcasts-Files/1034/What%20Physician%20Needs%20To%20Know%20About%20Human%20Trafficking%20-%204.pdf
- **ACOG – Clinical Opinion on Human Trafficking**
  https://www.acog.org/clinical/clinical-guidance/committee-opinion/articles/2019/09/human-trafficking
- **ASHRM – Creating Safe Havens in Physician Training**
  https://www.ashrm.org/education-events/creating-human-trafficking-victim-medical-safe-haven-resident-physician-education

- **Martha's Vineyard Hospital – Training Collaboration** https://mvhospital.org/marthas-vineyard-hospital-global-strategic-operatives-present-educational-training-focusing-on-human-trafficking/

#### List of excluded resources and reasons for exclusion

| Author / year | Name | Exclusion reason |
|---|---|---|
| ICAT, 2020 | 20th anniversary of the adoption of the UN Protocol to Prevent, Suppress and Punish Trafficking in Persons, especially Women and Children An analytical review | Different concept |
| Jarrell et. al., 2023 | A Case of Human Trafficking in Appalachia and What Emergency Physicians Can Learn from It. | Different study design |
| Azab & Levine, 2023 | A novel human trafficking curriculum | Different setting |
| Gifford, 2019 | A practical guide to conducting a child sexual abuse examination | Different study design |
| Jacobson et. al., 2022 | A protocol for a qualitative study on sex trafficking: Exploring knowledge, attitudes, and practices of physicians, nurses, and social workers in Ontario, Canada. | Different concept |
| Choi et. al., 2020 | A qualitative needs assessment of human trafficking in Ethiopia: recommendations for a comprehensive, coordinated response. | Different concept |
| Dhavala et. al., 2018 | A QUIP to improve staff engagement with a departmental safeguarding pathway for young people: A journey from failure to success | Different concept |
| Greenbaum, 2018 | A Short Screening Tool to Identify Victims of Child Sex Trafficking in the Health Care Setting | Duplicate |
| Vlades et. al., 2023 | A simulated pedagogical intervention to educate nurse practitioner students about human trafficking. | Different setting |
| Cooke-Sporing et. al., 2023 | A Simulation-Based Human Trafficking Curriculum for Emergency Medicine Residents | Not available |
| Young et. al., 2024 | A Teach-the-Teacher Module for Human Trafficking Bedside Instruction. | Different setting |
| Cole et. al., 2018 | A Theory-based Didactic Offering Physicians a Method for Learning and Teaching Others About Human Trafficking. | Different setting |
| Cole et al., 2018 | A theory-based didactic offering physicians a method for learning and teaching others about human trafficking. | Different concept |
| Langerman et. al., 2019 | Acceptability of Adolescent Social and Behavioral Health Screening in the Emergency Department. | Different population |
| Knudtzen et. al., 2022 | Accessing vulnerable undocumented migrants through a healthcare clinic including a community outreach programme: a 12-year retrospective cohort study in Denmark | Different outcome |
| Wells et. al., 2021 | Addressing Adolescent Safety in the Time of Telemedicine: A Modified Human Trafficking Standardized Patient Case | Different concept |
| Mishori and Ravi, 2015 | Addressing suspected labor trafficking in the office | Different publication type |
| Giannoukos et. al., 2018 | Advances in chemical sensing technologies for VOCs in breath for security/threat assessment, illicit drug detection, and human trafficking activity. | Different study design |
| Thomas-Smith et. al., 2020 | Advocacy & Pediatric Human Trafficking | Different study design |
| Pierce, 2012 | American Indian adolescent girls: vulnerability to sex trafficking, intervention strategies. | Different setting |

| | | |
|---|---|---|
| Moore and Williams, 2020 | An Audit of the child sexual exploitation risk questionnaire (CSERQ15) in Wales | Not available |
| Robitz et. al., 2022 | An integrated approach to providing care for people who have been trafficked. | Not available |
| Klein et. al., 2023 | Approaches to the teaching and evaluation of trauma-informed care principles in an emergency department setting: a systematic review | Different population |
| Fernandes et. al., 2016 | Are we 'spotting the signs?' | Not available |
| Jimenez et. al., 2015 | Aspects of abuse: commercial sexual exploitation of children. | Different study design |
| McAmis et. al., 2021 | Assessing Healthcare Provider Knowledge of Human Trafficking | Different concept |
| Panlilio et. al., 2019 | Assessing risk of commercial sexual exploitation among children involved in the child welfare system. | Different setting |
| Panlilio et al., 2019 | Assessing risk of commercial sexual exploitation among children involved in the child welfare system. | Different setting |
| Hurst et. al., 2020 | Assessing the Utility of a Statewide Human Trafficking Screening Tool in Colorado | Not available |
| Williams et. al., 2017 | Assessment of risk of child sexual exploitation at initial health assessments for looked after children: How well do we do? | Not available |
| Ward et. al., 2019 | Association between STI and child sexual exploitation in children under 16 years old attending sexual health clinics in England: findings from a case-control study. | Different concept |
| Son 2014 | Barriers to Access, Disclosure, and Identification in Healthcare for Potentially Trafficked Youth in Vermont, in 142nd APHA Annual Meeting and Exposition. | Different concept |
| Garg et. al., 2020 | Barriers to the access and utilization of healthcare for trafficked youth: A systematic review. | Different concept |
| Landers 2017 | Baseline Characteristics of Dependent Youth Who Have Been Commercially Sexually Exploited: Findings from a Specialized Treatment Program | Different concept |
| Pocock 2018 | Because if we talk about health issues first, it is easier to talk about human trafficking; findings from a mixed methods study on health needs and service provision among migrant and trafficked fishermen in the Mekong | Different concept |
| Stoklosa et. al., 2022 | Because the resources aren't there, then we fail. We fail as a society: A Qualitative Analysis of Human Trafficking Provider Perceptions of Child Welfare Involvement among Trafficked Mothers. | Different concept |
| Twis et. al., 2024 | Beyond Victim Identification: A Practitioner's Guide to Designing a Youth Anti-Sex Trafficking Advocacy Program. | Different concept |
| McDonald et. al., 2023 | Building a specialized model of care for youth involved in sex trafficking in child welfare: A systematic review and interviews with experts-by-experience. | Different concept |
| Chisolm-Straker, 2019 | Building RAFT: Trafficking Screening Tool Derivation and Validation Methods | Duplicate |
| Chisolm-Straker, 2019 | Building RAFT: Trafficking Screening Tool Derivation and Validation Methods | Duplicate |
| Lefevre 2017 | Building Trust with Children and Young People at Risk of Child Sexual Exploitation: The Professional Challenge | Different concept |
| Katsanis et. al., 2019 | Caring for trafficked and unidentified patients in the EHR shadows: Shining a light by sharing the data. | Different concept |
| IOM 2009 | Caring for Trafficked Persons: Guidance for Health Providers. | Different concept |
| Rafferty 2016 | Challenges to the rapid identification of children who have been trafficked for commercial sexual exploitation | Different concept |
| Lepianka and Colbert, 2020 | Characteristics and Healthcare Needs of Women Who Are Trafficked for Sex in the United States: An Integrative Literature Review. | Different concept |
| Talbott et. al., 2023 | Characteristics and Perspectives of Human Trafficking Education: A Survey of U.S. Medical School Administrators and Students. | Different setting |
| Varma et. al., 2015 | Characteristics of child commercial sexual exploitation and sex trafficking victims presenting for medical care in the United States. | Different concept |
| Suniega et. al., 2022 | Child Abuse: Approach and Management. | Different study design |

| | | |
|---|---|---|
| Greenbaum, 2021 | Child Labor and Sex Trafficking. | Different publication type |
| No authors listed, 2018 | Child Labor Trafficking Essentials for Forensic Nurses. | Different study design |
| Greenbaum, 2018 | Child Sex Trafficking and Commercial Sexual Exploitation. | Different study design |
| Greenbaum and Crawford-Jakubiak, 2015 | Child sex trafficking and commercial sexual exploitation: health care needs of victims. | Different study design |
| Safeguarding children parthnership, 2016 | Child Sexual Exploitation (CSE) Risk Assessment Toolkit | Different setting |
| Mason-Jones and Loggie, 2020 | Child sexual exploitation. An analysis of serious case reviews in England: poor communication, incorrect assumptions and adolescent neglect. | Different concept |
| Thakur and Gurbani, 2021 | Child trafficking & human trafficking: Legal aspects arole of a doctor | Different study design |
| White, 2022 | Collaboration and Challenges in Antitrafficking Task Forces | Different study design |
| Chung and English, 2015 | Commercial sexual exploitation and sex trafficking of adolescents. | Different study design |
| Benavente et. al., 2022 | Commercial Sexual Exploitation of Children and Adolescents in Europe: A Systematic Review. | Different study design |
| Hornor and Sherfield, 2018 | Commercial Sexual Exploitation of Children: Health Care Use and Case Characteristics. | Different concept |
| Bauer and Magana, 2018 | Commercial sexual exploitation of children: What healthcare providers do (and don't) know | Different setting |
| Gallo et. al., 2022 | Community Health Centers and Sentinel Surveillance of Human Trafficking in the United States. | Different study design |
| Jadhav and Gandhewar, 2022 | Comparative Analysis of Various Machine Learning Models for Child Safety and Security System for Protecting Them from Child Trafficking and Assault | Different setting |
| Kachelski et. al., 2023 | Comparative healthcare use by adolescents screening positive for sexual exploitation. | Different concept |
| Palines 2020 | Comparing mental health disorders among sex trafficked children and three groups of youth at high-risk for trafficking: A dual retrospective cohort and scoping review. | Different concept |
| Wirtz et. al., 2016 | Comprehensive development and testing of the ASIST-GBV, a screening tool for responding to gender-based violence among women in humanitarian settings. | Different population |
| Organización Mundial de la Salud, 2013 | Comprender y abordar la violencia contra las mujeres: trata de personas | Different concept |
| Hurts et. al., 2021 | Confidential Screening for Sex Trafficking Among Minors in a Pediatric Emergency Department | Duplicate |
| Institute of Medicine; National Research Council, 2014 | Confronting Commercial Sexual Exploitation and Sex Trafficking of Minors in the United States: A Guide for Providers of Victim and Support Services | Different concept |
| Brown et. al., 2024 | Cross-sector collaboration in Project Catalyst: Creating state partnerships to address the health impact of intimate partner violence | Different setting |
| Herrero-Villoria et. al., 2022 | Cultural Adaptation and Validation into Spanish of the Scale to Measure Attitudes Towards the Sex Trafficking of Women and Girls in Students of the University of Salamanca. | Different setting |
| Burgess et. al., 2008 | Cyber child sexual exploitation. | Different setting |
| Pollock et. al., 2024 | Dermatology's role in the fight against human trafficking: A report from the AAD Ad Hoc Task Force and call to action. | Different study design |
| Murphy et. al., 2016 | Development and Pilot Test of a Commercial Sexual Exploitation Prevention Tool: A Brief Report. | Different population |
| Gray et. al., 2024 | Development of an Index to Measure the Exposure Level of UN Peacekeeper-Perpetrated Sexual Exploitation/Abuse in Women/Girls in the Democratic Republic of Congo. | Different setting |
| Interiano-Shiverdecker et. al., 2023 | Development of Child Sex Trafficking Counseling Competencies in the United States: A Delphi Study. | Different setting |

| | | |
|---|---|---|
| Ministério Público Federal, 2014 | Diálogos da cidadania: tráfico de pessoas: conhecer para se proteger | Different study design |
| Thompson et. al., 2017 | Do clinicians receive adequate training to identify trafficked persons? A scoping review of NHS Foundation Trusts. | Different concept |
| Schroeder et. al., 2024 | Do Social Service Interventions for Human Trafficking Survivors Work? A Systematic Review and Meta-Analysis. | Different concept |
| Ministério da Saúde, 2022 | Documento técnico: enfrentamento ao tráfico de pessoas para profissionais de saúde | Different concept |
| Goldberg et. al., 2017 | Domestic Minor Sex Trafficking Patients: A Retrospective Analysis of Medical Presentation. | Different concept |
| Moore et. al., 2021 | Domestic Minor Sex Trafficking: A Case Series of Male Pediatric Patients. | Different concept |
| Kaplan et. al., 2018 | Domestic Minor Sex Trafficking: Medical Follow-up for Victimized and High-Risk Youth. | Different study design |
| Leitch & Snow, 2010 | Domestic minor sex trafficking: Practitioner guide and intake tool. | Duplicate |
| Coughlin et. al., 2020 | Educating pediatric health-care providers about human trafficking. | Different study design |
| López-Domene et. al., 2019 | Emergency Care for Women Irregular Migrants Who Arrive in Spain by Small Boat: A Qualitative Study. | Different population |
| Peeler, 2019 | Emergency Care of Pediatric Asylum Seekers in the United States | Different study design |
| Muste et. al., 2023 | Emergency department evaluation of nurse triage questions about safe-at-home and abuse or neglect in traumatic ocular injuries. | Different population |
| Khanna et. al., 2020 | Empowering nurses: Using knowledge to screen & identify victims of human trafficking | Different setting |
| Harlow et al., 2019 19 | EMS professionals: critical partners in human trafficking response | Different publication type |
| De Vries et al., 2020 | Enhancing the identification of commercial sexual exploitation among a population of high-risk youths using predictive regularization models. | Different setting |
| Anderson and Modi, 2021 | Evaluating a virtual interdisciplinary human trafficking training program in the emergency department | Not available |
| Panlilio et al., 2022 | Evaluating and validating the classification accuracy of a screening instrument to assess risk for commercial sexual exploitation of child welfare-involved children and adolescents | Different setting |
| Shery-Ann et. al., 2018 | Evaluation of a Screening Tool for Child Sex Trafficking Among Patients with High-Risk Chief Complaints in a Pediatric Emergency Department | Duplicate |
| Kim et. al., 2023 | Evaluation of Services for the Commercial Sexual Exploitation of Children and Youth: A Scoping Review. | Different concept |
| Dimitropoulos et. al., 2022 | Experiences of Canadian mental health providers in identifying and responding to online and in-person sexual abuse and exploitation of their child and adolescent clients. | Different concept |
| Timmel et. al., 2020 | From family violence to trauma informed care: A multidisciplinary workshop for medical students | Different setting |
| Winks et. al., 2023 | Frontline Medical Professionals' Ability to Recognize and Respond to Suspected Youth Sex Trafficking. | Different concept |
| Barbosa, 2023 | Gender Dysphoria, ASD, and Sex Trafficking: Addressing Barriers in the Mental Health Care of Neurodiverse Gender-Minority Youth in a Rural State | Different concept |
| ILO, Walk Free & OIM, 2022 | Global estimates of modern slavery | Different concept |
| International Labour Office (ILO), 2022 | Hard to see, harder to count Survey guidelines to estimate forced labour of adults and children | Different concept |
| Viergever et. al., 2015 | Health care providers and human trafficking: what do they know, what do they need to know? Findings from the Middle East, the Caribbean, and Central America. | Different concept |
| Isaac 2011 | Health Care Providers’ Training Needs Related to Human Trafficking: Maximizing the Opportunity to Effectively Screen and Intervene. | Different study design |
| Ehrhardt-Humbert et. al., 2023 | Health Care Utilization by Pediatric Human Trafficking Victims in an Urban Health Care System | Different concept |
| Eappen et. al., 2022 | Health Services to Meet Physical, Mental, and Social Needs of 126 Females Who Survived Boko Haram Abduction and Captivity: Providers' Perspective. | Different concept |

| | | |
|---|---|---|
| Ertl et. al., 2020 | Healthcare needs and utilization patterns of sex-trafficked youth: Missed opportunities at a children's hospital. | Different concept |
| Andersson and Örmon, | Healthcare providers' experience of identifying and caring for women subjected to sex trafficking: a qualitative study. | Different concept |
| Alhajji et. al., 2021 | Helping survivors of human trafficking | Different publication type |
| Spadafore et. al., 2021 | Histories of trauma: A qualitative analysis of lifetime traumatic experiences among emergency department patients. | Different population |
| Covenant House, 2013 | Homelessness, survival sex and human trafficking: As experienced by the youth of Covenant House New York | Different setting |
| Gerassi et. al., 2023 | How Do Providers Assess Young People for Risk of Sex Trafficking? Observed Indicators, Follow-Up, and Assessment Questions from a Sample Social Service Providers. | Different setting |
| Jessop et. al., 2018 | How good are we at 'spotting the signs'? | Not available |
| Cooper, 2016 | How to spot signs of child sexual exploitation | Different publication type |
| Buśet. al., 2019 | Human trafficking – Multinational challenge for forensic science | Different publication type |
| Shekhar and Macias-Konstantopoulos, 2023 | Human Trafficking and Emergency Medical Services (EMS). | Different publication type |
| Ross 2015 | Human trafficking and health: A cross-sectional survey of NHS professionals' contact with victims of human trafficking. | Different concept |
| Oram 2016 | Human trafficking and health: A survey of male and female survivors in England | Different concept |
| Deutscher et. al., 2017 | Human trafficking awareness, a learning module for improved recognition of victims in the emergency room | Different setting |
| Findlay et. al., 2016 | Human trafficking didactic session resulted in improved awareness | Different setting |
| Das et. al., 2023 | Human Trafficking Education: A Pilot Study of Integration into Medical School Curriculum. | Different setting |
| Schwarz et. al., 2016 | Human Trafficking Identification and Service Provision in the Medical and Social Service Sectors. | Different study design |
| Einbond et. al. 2020 | Human Trafficking in Adolescents: Adopting a Youth-centered Approach to Identification and Services. | Different study design |
| Beck et. al., 2017 | Human Trafficking in Ethiopia: A Scoping Review to Identify Gaps in Service Delivery, Research, and Policy. | Different concept |
| Patel 2010 | Human Trafficking in the Emergency Department. | Different study design |
| Tiller and Reynolds, 2020 | Human Trafficking in the Emergency Department: Improving Our Response to a Vulnerable Population. | Different study design |
| Raker and Hromadik, 2021 | Human Trafficking in the Radiology Setting | Different publication type |
| The Lancet Regional Health – Western Pacific, 2022 | Human trafficking is more than a crime | Different publication type |
| State of Florida Department of Children and Family. | Human Trafficking of Children Indicator Tool. | Different setting |
| Peck and Meadows-Oliver, 2019 | Human Trafficking of Children: Nurse Practitioner Knowledge, Beliefs, and Experience Supporting the Development of a Practice Guideline: Part One. | Different study design |
| Peck JL, Doiron ML | Human trafficking policies of professional nursing organizations: Opportunity for innovative and influential policy voice. | Different concept |
| Baldwin 2009 | Human Trafficking Victims: At an Abortion Clinic Near You? 2009 | Not available |
| Scott-Tilley and Crites, 2016 | Human Trafficking, Sexual Assault, or Something Else? A Complicated Case with an Unexpected Outcome. | Different study design |
| Shandro et. al., 2016 | Human Trafficking: A Guide to Identification and Approach for the Emergency Physician. | Different study design |
| ACOG, 2019 | Human Trafficking: ACOG COMMITTEE OPINION | Different publication type |

| | | |
|---|---|---|
| Leslie, 2018 | Human Trafficking: Clinical Assessment Guideline. | Different study design |
| Pulvino et. al., 2023 | Human Trafficking: Screening and Linkage to Care | Not available |
| Pulvino, 2023 | Human Trafficking: Screening and Linkage to Care | Duplicate |
| Trout, 2010 | Human trafficking: the role of nurses in identifying and helping victims. | Different publication type |
| Dovydaitis 2009 | Human trafficking: the role of the health care provider. | Different study design |
| Cheetham and Hurst, 2022 | Human Trafficking: When to Suspect in the Pediatric Emergency Department? | Different study design |
| Pegram, 2020 | Human trafficking: would you recognize it? | Different study design |
| Hachey & Phillippi, 2017 | Identification and management of human trafficking victims in the emergency department | Different study design |
| Hachey and Phillippi, 2017 | Identification and Management of Human Trafficking Victims in the Emergency Department. | Different study design |
| Gibbons and Stoklosa, 2016 | Identification and Treatment of Human Trafficking Victims in the Emergency Department: A Case Report. | Different study design |
| Grosgogeat et. al., 2024 | Identification of a Human Trafficking Victim: A Simulation. | Different publication type |
| Baldwin 2011 | Identification of human trafficking victims in health care settings. | Different concept |
| Rambhatla et. al., 2021 | Identification of skin signs in human-trafficking survivors. | Different concept |
| Tracy y Macias-Konstantopoulos, 2017 | Identifying and assisting sexually exploited and trafficked patients seeking Women's health care services. | Different study design |
| Hunt et. al., 2020 | Identifying human trafficking in adults | Different publication type |
| Weiss et. al., 2023 | Identifying Human Trafficking in the Hospital Via an Abuse Screening Tool | Not available |
| Nguyen et. al., 2018 | Identifying Human Trafficking Victims on a Psychiatry Inpatient Service: a Case Series. | Different concept |
| Mostajabian et. al., 2019 | Identifying Sexual and Labor Exploitation among Sheltered Youth Experiencing Homelessness: A Comparison of Screening Methods. | Different setting |
| Sinha 2019 | Identifying victims of human trafficking in central Pennsylvania: A survey of health-care professionals and students. J | Different concept |
| Greenbaum, 2016 | Identifying Victims of Human Trafficking in the Emergency Department | Not available |
| Coughlin et. al., 2019 | Identifying victims of sex trafficking: Assessing medical student knowledge and confidence after a brief workshop | Different publication type |
| Balchan, 2018 | Identifying youth at risk for commercial sexual exploitation of children (CSEC) in a foster care clinic | Not available |
| Brandt 2018 | Identifying youth at risk for commercial sexual exploitation within child advocacy centers: a State-wide pilot study | Different setting |
| Song, 2021 | Impact of COVID-19 on the exploitation of children | Different study design |
| Ficker et. al., 2023 | Incidence of Sexually Transmitted Infections and Pregnancy Among Adolescent Sex Trafficking Victims | Different concept |
| Raj et. al., 2019 | Incorporating Clinical Associations of Domestic Minor Sex Trafficking into Universal Screening of Adolescents. | Different concept |
| OIM, 2019 | Informe sobre las Migraciones en el Mundo 2020 | Different population |
| UNODC, 2020 | INTERLINKAGES BETWEEN Trafficking in Persons and Marriage | Different setting |
| Hartsock and Helft, 2019 | International Travel for Living Donor Kidney Donation: A Proposal for Focused Screening of Vulnerable Groups. | Different publication type |
| Lamb-Susca and Clements, 2018 | Intersection of Human Trafficking and the Emergency Department. | Different publication type |
| Lamb-Susca et al., 2018 20 | Intersection of human trafficking and the emergency department. | Different study design |
| Shared Hope InternationalLeitch L, Snow M. | Intervene Practitioner Guide and Intake Tool | Not available |

| | | |
|---|---|---|
| Lin et. al., 2024 | Intimate Partner Violence and Human Trafficking Screening and Services in Primary Care Across Underserved Communities in the United States-Initial Examination of Trends, 2020-2021. | Different concept |
| Ulibarri et. al., 2017 | Introduction to Special Section: Research, Treatment, and Policy Regarding Trafficking and Sexual Exploitation of Children and Adolescents | Different setting |
| Resolution Hope National Campaign to End Child Trafficking., 2013 | Know What to Look For. | Not available |
| Lawrence et. al., 2020 | Knowledge Base of Nurses Before and After a Human Trafficking Continuing Education Course. | Different concept |
| Greenbaum et. al., 2022 | Labor trafficking of children and youth in the United States: A scoping review. | Different concept |
| UN, 2020 | LEGISLATIVE GUIDE FOR THE PROTOCOL TO PREVENT, SUPPRESS AND PUNISH TRAFFICKING IN PERSONS, ESPECIALLY WOMEN AND CHILDREN | Different setting |
| OIM, 2014 | LINEAMIENTOS REGIONALES PARA LA IDENTIFICACION PRELIMINAR DE PERFILES Y MECANISMOS DE REFERENCIA DE POBLACIONES MIGRANTES EN CONDICION DE VULNERABILIDAD | Different concept |
| Enríquez, 2015 | Lo psicológico en el plan integral de lucha contra la trata de personas Colombia 2007- 2012 | Different setting |
| Bick et. al., 2017 | Maternity care for trafficked women: Survivor experiences and clinicians' perspectives in the United Kingdom's National Health Service. | Different concept |
| Chisolm-Straker, 2018 | Measured Steps: Evidence-based Anti-Trafficking Efforts in the Emergency Department. | Different study design |
| Pacífico, 2011 | Mecanismos institucionais de prevenção e combate ao tráfico de pessoas no Brasil | Different publication type |
| Kaplan, 2023 | Medical Presentations and Needs of Exploited Youth in the Pediatric Emergency Setting | Different study design |
| Beck, 2015 | Medical Providers’ Understanding of Sex Trafficking and Their Experience With At-Risk Patients | Different concept |
| Borham et. al., 2019 | Medical School Curricular Development: Sex Trafficking, Adverse Childhood Experiences, and Trauma-Informed Car | Different setting |
| Weis et. al., 2017 | Medical tourism: The role of the primary care provider | Different publication type |
| Altun et. al., 2017 | Mental health and human trafficking: Responding to survivors' needs | Different publication type |
| Iglesias-Rios 2018 | Mental health, violence and psychological coercion among female and male trafficking survivors in the greater Mekong sub-region: A cross-sectional study | Different concept |
| Ruiz-Gonzalez et. al., 2022 | Midwives' experiences and perceptions in treating victims of sex trafficking: A qualitative study. | Different concept |
| Migration data portal, 2023 | Migration data in Eastern Africa | Different publication type |
| Miller et. al., 2007 | Migration, sexual exploitation, and women's health: a case report from a community health center. | Different study design |
| Mæng et. al., 2014 | Mobile health initiative for foreign prostitutes in the Region of Central Jutland and the Region of North Jutland. | Different concept |
| UN, 2020 | Model Legislative Provisions Against Trafficking In Persons | Different setting |
| Such et. al., 2020 | Modern slavery and public health: a rapid evidence assessment and an emergent public health approach. | Different concept |
| Minderoo Foundation’s Walk Free initiative and the Human Rights Resources and Energy Collaborative (HRREC) | Modern Slavery Response & Remedy Framework | Different concept |

| | | |
|---|---|---|
| Greenbaum et. al., 2018 | Multi-level prevention of human trafficking: The role of health care professionals. | Different concept |
| Paraskevas and Bookes 2018 | Nodes, guardians and signs: Raising barriers to human trafficking in the tourism industry. | Different setting |
| Bono-Neri and Toney-Butler, 2023 | Nursing students' knowledge of and exposure to human trafficking content in undergraduate curricula. | Different concept |
| Ropero-Padilla et. al., 2022 | Nursing students' perceptions of identifying and managing sex trafficking cases: A focus group study. | Different setting |
| NWG, 2017 | NWG Network Child Sexual Exploitation (CSE) Risk Assessment Tool | Not available |
| McConkey et. al., 2023 | Operationalizing Child Sex Trafficking Screening in an Academic Emergency Department | Not available |
| Dimas et. al., 2022 | Operations research and analytics to combat human trafficking: A systematic review of academic literature. | Different concept |
| No authors listed, 2018 | Oral and Dental Aspects of Child Abuse and Neglect. | Different publication type |
| Ministerio de Salud de Peru, 2021 | Orientaciones técnicas para el cuidado integral de la salud mental del niñas, niños y adolescentes víctimas y sobrevivientes de trata de personas. Documento técnico | Different concept |
| Langerman et. al., 2018 | Patient and caregiver attitudes towards comprehensive behavioral health screening in the emergency department | Not available |
| Titchen et. al., 2023 | Physician Understanding of Youth Labor and Sex Trafficking: A Need for Training | Not available |
| Ministerio del Interior de Peru, 2021 | Plan nacional contra la trata de personas 2017 - 2021 | Different setting |
| Speck et. al., 2018 | Policy brief on the nursing response to human trafficking. | Different publication type |
| Anderson et. al., 2024 | Preliminary Evidence of Validity for the Verbally Pressured and Illegal Sexual Exploitation Modules of the Sexual Experiences Survey-Victimization. | Different setting |
| Ma et al., 2020 23 | Preparing residents to deal with human trafficking. | Different setting |
| Franchek-Roa, 2017 | Preparing your healthcare system to identify and respond to victims of abuse, neglect and exploitation | Not available |
| Dank, 2017 | Pretesting a Human Trafficking Screening Tool in the Child Welfare and Runaway and Homeless Youth Systems | Different setting |
| Ottisova et. al., 2016 | Prevalence and risk of violence and the mental, physical and sexual health problems associated with human trafficking: an updated systematic review. | Different concept |
| UN | Project Delta 8.7 Informing policies that contribute to achieving SDG Target 8.7 on modern slavery, human trafficking, and forced and child labour. | Different publication type |
| Ambagtsheer et. al., 2018 | Proposal for an anonymous reporting code organ trafficking: how transplant professionals can play a role in preventing this trafficking | Different concept |
| Cooper, 2015 | Protecting children from sexual exploitation. | Different publication type |
| Ministerio de Salud Pública y Asistencia Social, 2012 | Protocolo de atención en salud integral con pertinencia cultural para la niñez y la adolescencia en situación de trabajo infantil y sus peores formas | Different concept |
| Ministerio del Interior de Peru, 2018 | Protocolo intersectorial para la prevención y persecución del delito y la protección. atención y reintegración de víctimas de trata de personas - D.S. N° 005-2016-IN | Different setting |
| Gordon et. al., 2018 | Psychiatry's Role in the Management of Human Trafficking Victims: An Integrated Care Approach. | Different study design |
| Basu et. al., 2021 | Recognizing and intervening in child sex trafficking. | Different study design |
| Prakash et. al., 2023 | Recognizing Human Trafficking in Radiology | Different publication type |
| Jouk, 2021 | Recognizing Suspected Human Trafficking in the Pediatric Intensive Care Unit | Different concept |
| Danaher et. al., 2018 | Recognizing, diagnosing, and preventing child maltreatment: an update for pediatric clinicians. | Different population |

| | | |
|---|---|---|
| Taskforce on Trafficking of Women and Girls, 2014 | Report of the taskforce on trafficking of women and girls. | Different concept |
| Hoffman and Argeros, 2022 | Researching the Effectiveness of an Online Human Trafficking Awareness Program Among Community Health Nursing Students. | Different setting |
| Tambini Stollwerck et. al., 2024 | Responding to human trafficking among refugees: prevalence and test accuracy of a modified version of the adult human trafficking screening tool. | Different setting |
| Williamson et. al., 2020 | Responding to the health needs of trafficked people: A qualitative study of professionals in England and Scotland. | Different concept |
| Talbott et. al., 2020 | Review of Published Curriculum on Sex Trafficking for Undergraduate Medical Trainees. | Different setting |
| Jaeckl and Laughon, 2021 | Risk Factors and Indicators for Commercial Sexual Exploitation/Domestic Minor Sex Trafficking of Adolescent Girls in the United States in the Context of School Nursing: An Integrative Review of the Literature. | Different concept |
| Kent and Medway, Safeguarding Children Board, 2017 | Safeguarding children at risk of sexual exploitation. Risk assessment toolkit. | Not available |
| Wilkinson and Cranston, 2015 | Safeguarding for anesthetists: working to protect children | Different population |
| Dwyer and Rogstad, 2022 | Safeguarding, child sexual exploitation and sexual assault | Different publication type |
| San Luis Obispo County, 2014 | San Luis Obispo County CSEC collaborative response team commercial sexual exploitation of children (CSEC screening tool) | Duplicate |
| Ministério da Saúde, 2014 | Saúde, migração, tráfico e violência contra mulheres: o que o SUS precisa fazer: caderno pedagógico | Different publication type |
| Ministério da Saúde, 2013 | Saúde, migração, tráfico e violência contra mulheres: o que o SUS precisa saber | Different publication type |
| Spencer-Hughes 2017 | Screening for child sexual exploitation in online sexual health services: An exploratory study of expert views | Different concept |
| Spencer-Hughes et. al., 2017 | Screening for Child Sexual Exploitation in Online Sexual Health Services: An Exploratory Study of Expert Views. | Different setting |
| Costelloet. al., 2021 | Screening for human trafficking in the pediatric emergency department: A pre-and post-intervention study | Not available |
| Indiana Coalition Against Sexual Assault. | Screening Tool for Victims of Human Trafficking. | Different setting |
| The US Department of Health and Human Services | Screening Tool for Victims of Human Trafficking. | Not available |
| O'Connell and Lomax, 2016 | Service evaluation of the use of the young person's proforma in relation to central and community sexual health clinics | Not available |
| McAlpine et. al., 2016 | Sex trafficking and sexual exploitation in settings affected by armed conflicts in Africa, Asia and the Middle East: systematic review. | Different concept |
| Barron 2016 | Sex trafficking assessment and resources (STAR) for pediatric attendings in Rhode Island | Different concept |
| Haney et. al., 2020 | Sex Trafficking in the United States: A Scoping Review. | Different setting |
| Bortel et al., 2008 | Sex trafficking needs assessment for the State of Minnesota | Different setting |
| Chaffee and English, 2015 | Sex trafficking of adolescents and young adults in the United States: healthcare provider's role. | Different study design |
| Moore et. al., 2017 | Sex Trafficking of Minors. | Different setting |
| Scott-Wellington et. al., 2021 | Sex trafficking screening tool in the emergency department | Different publication type |
| Lorvinsky et. al., 2023 | Sex trafficking survivors' experiences with the healthcare system during exploitation: A qualitative study. | Different concept |
| Richie-Zavaleta et al., 2020 26 | Sex trafficking victims at their junction with the healthcare setting-a mixed-methods inquiry. | Different concept |
| Rapoza, 2022 | Sex Trafficking: A Literature Review with Implications for Health Care Providers. | Different study design |

| | | |
|---|---|---|
| UN Dept of Field Support, 2018 | Sexual Exploitation and Abuse: Risk management toolkit | Different setting |
| South Gloucestershire Council, 2020. | Sexual exploitation risk assessment framework (SERAF) | Not available |
| Clutton & Coles, 2007 | Sexual exploitation risk assessment framework: A pilot study. | Different setting |
| Mays A, Harvill Z, Mejia J. | Sexually Exploited Children Screening Protocol: A Multidisciplinary Model Designed for the Clinical an School Health Setting. | Different setting |
| Ashby et. al., 2015 | Spotting the Signs: a national toolkit to help identify young people at risk of child sexual exploitation. | Different publication type |
| Begley et. al., 2022 | Student Perceptions of an Interprofessional Short Course Designed to Increase Awareness of Human Trafficking. | Different setting |
| Wretman et. al., 2021 | Study protocol for an evaluability assessment of an anti-human trafficking program. | Different setting |
| Webster, 2018 | Sutton practice toolkit for safeguarding children from sexual exploitation (CSE). (2nd ed.). | Not available |
| Fang et. al., 2018 | Tattoo Recognition in Screening for Victims of Human Trafficking. | Different study design |
| Weiss and Kiluk,2018 | Teaching human trafficking to 3RD year medical students | Different setting |
| Stevens and Berishaj, 2016 | The Anatomy of Human Trafficking: Learning About the Blues: A Healthcare Provider's Guide. | Different study design |
| Kim et. al., 2018 | The anti-human trafficking collaboration model and serving victims: Providers' perspectives on the impact and experience. | Different concept |
| Magdaleno et. al., 2023 | The Development and Implementation of a Forensic Education Module for Nebraska Critical Access Providers: A Pilot Study. | Different concept |
| Tuharyati et. al., 2020 | The eradication of women and children trafficking in jember regency in relations to the legal protection for victims in the health perspective | Different concept |
| Adam and Webb, 2018 | The Exploitation of Children: Understanding Human Sex Trafficking | Different concept |
| Lederer 2014 | The Health Consequences of Sex Trafficking and Their Implications for Identifying Victims in Healthcare Facilities. | Different concept |
| Wick, 2024 | The Impact of Pharmacists and Pharmacy Technicians in Recognizing and Responding to Human Trafficking | Not available |
| Forbes et. al., 2017 | THE in betweenness: 16 & 17 year olds attending SRH are vulnerable | Not available |
| Koss et. al., 2024 | The Revised Sexual Experiences Survey Victimization Version (SES-V): Conceptualization, Modifications, Items and Scoring. | Different study design |
| Peters-Mosquera et. al., 2023 | The Role Nurses Can Play in Addressing and Preventing the Prevalence of Missing or Murdered Indigenous Women and Girls (MMIWG). | Different concept |
| Mercer et. al., 2018 | The use of standardized patients to increase medical student awareness of and confidence in screening for human trafficking | Different setting |
| Boparan et. al., 2020 | To what extent are community pharmacists ‚Spotting the Signs’ and acting appropriately according to the child sexual exploitation safeguarding training? | Not available |
| Tomsett et. al., 2024 | Tools for the identification of victims of domestic abuse and modern slavery in remote services: A systematic review. | Different setting |
| Shah et. al., 2021 | TraffickED and TraumatizED: The Effectiveness of a Human Trafficking Seminar in Building Confidence in Trauma-Informed Care | Different setting |
| Cannon et. al., 2018 | Trafficking and Health: A Systematic Review of Research Methods. | Different concept |
| ICAT, 2021 | TRAFFICKING IN PERSONS FOR THE PURPOSE OF ORGAN REMOVAL | Different concept |
| Ary and Maia, 2008 | Tráfico de seres humanos na sociedade internacional contemporânea: globalização, políticasmigratórias e os esforços multilaterais de combate | Different setting |
| Bechtel et. al., 2022 | Training Experiences of Emergency Department Providers in the Recognition of Child Trafficking. | Different concept |
| Powell et al., 2017 | Training US health care professionals on human trafficking: where do we go from here? | Different concept |
| World Health Organization, 2012 | Understanding and addressing violence against women: human trafficking | Different publication type |

| | | |
|---|---|---|
| Munro-Kramer et. al., 2022 | Understanding Health Facility Needs for Human Trafficking Response in Michigan. | Different concept |
| Chisolm-Straker et al., 2018 | Universal screening for trafficking in the emergency department: RAFT development and validation | Duplicate |
| United Nations 2021 | UNODC TOOLKIT For mainstreaming Human Rights and Gender Equality into criminal justice interventions to address trafficking in persons and smuggling of migrants | Different setting |
| Severini et. al., 2015 | Use of ancestry-informative markers as a scientific tool to combat the illegal traffic in human kidneys | Different population |
| Armstrong and Greenbaum, 2019 | Using Survivors' Voices to Guide the Identification and Care of Trafficked Persons by U.S. Health Care Professionals: A Systematic Review. | Different study design |
| Chisolm-Straker et. al., 2021 | Validation of a screening tool for labor and sex trafficking among emergency department patients | Duplicate |
| Basson, 2023 | Validation of the commercial sexual exploitation-identification tool (CSE-IT). Technical report. | Duplicate |
| Cano et. al., 2023 | Víctimas de trata de seres humanos: una realidad emergente en medicina forense | Different concept |
| Pocock et. al., 2021 | Victims or suspects? Identifying and assisting potentially trafficked fishermen: A qualitative study with stakeholders and first responders in Thailand. | Different setting |
| Riley, 2019 | When slavery hides in the symptoms - Are we ready to see it? | Different publication type |
| Tomsett et. al., 2025 | Tools for the identification of victims of domestic abuse and modern slavery in remote services: A systematic review. | Different concept |
| Twis et. al., 2024 | Beyond Victim Identification: A Practitioner's. Guide to Designing a Youth Anti-Sex Trafficking. Advocacy Program | Different concept |
| Shirazi et. al., 2024 | Human trafficking screening in Saskatoon Emergency Departments: What can be learned from high-risk patient presentations? | Different concept |
| Alnour et. al., 2022 | Global Practices and Policies of Organ Transplantation and Organ Trafficking | Different concept |
| Ambagtsheer et. al., 2016 | On Patients Who Purchase Organ Transplants Abroad | Different concept |
| Irish et. al., 2024 | International Travel for Organ Transplantation: A Survey of Professional Experiences and Attitudes Toward Data Collection and Reporting | Different concept |
| Mishra et. al., 2020 | mtDNA Analysis: A Valuable Tool to Establish Relationships in Live. Related Organ Transplants | Different concept |
| Hosey et. al., 2025 | Barriers to and facilitators of human trafficking screening in the healthcare setting: a scoping review protocol | Different publication type |
| Mangual et. al., 2024 | 54003 Role of the Dermatologist in Identifying and Advocating for Those Affected by Human Trafficking: A Needs Assessment in High Density Regions of Human Trafficking within the United States | Different publication type |
| Saucedo et. al., 2024 | Western Journal of Emergency Medicine: Integrating Emergency Care with Population Health | Different publication type |
| Roe-Sepowitz et. al., 2024 | PEARR tool training and implementation: building awareness of violence and human trafficking in a hospital system | Duplicate |
| The Advocated for Human Rights | Labor Trafficking Self-Assessment Card | Different setting |
| Missouri Hospital Association | Human Trafficking Toolkit: Guidance and Resources to Help Hospitals Combat Human Trafficking | Not available |
| Michigan Department of Health and Human Services, 2017 | HUMAN TRAFFICKING SCREENING TOOL – ONGOING CASES | Different setting |
| Michigan Department of Health and Human Services, 2017 | Human Trafficking of Children Protocol | Different setting |
| Macias-Konstantopoulos, 2018 | Adult Human Trafficking Screening Tool and Guide | Duplicate |

| | | |
|---|---|---|
| Labor International Office of United Nations, 2009 | Operational indicators of trafficking in human beings | Different setting |
| Hachey, 2017 | Identification and Management of Human Trafficking Victims in the Emergency Department | Different study design |
| Sonsiadek et. al. 2024 | Development of a Human Trafficking Flowsheet for Clinical Forensic Examiners | Duplicate |
| He et. al. 2020 | Use of Forensic DNA Testing to Trace Unethical Organ Procurement and Organ Trafficking Practices in Regions that Block Transparent Access to their Transplant Data | Different concept |
