## Appendix 2 for "Human Trafficking Detection in Health Care Settings: A Scoping Review"

#### Contend

### Tables

#### Table 1 Screening Tools Validation Studies

| Reference | Name | Population / Setting | Type of Validation / Evaluation of diagnosis properties | Outcome |
|---|---|---|---|---|
| Peterson, L.J. 2022 (1)<br><br>U.S. | A short, 6-question screening tool -Greenbaum et al. | Adolescents 11–17 years / EDs associated with the M. Health Fairview University of Minnesota Medical Center. | Diagnosis properties: Sensitivity, specificity and PPV | Of the 75 youth evaluated by FNE, no significant demographic differences were found between "positive Child Sex Trafficking (CST)" and "negative/at risk" groups, except for self-identified gender; 4 nonbinary individuals were in the "positive CST" group. Only the number of sex partners significantly differentiated the groups (p = 0.0094). The modified Short Screen for Child Sex Trafficking (SSCST) score (3+) failed to differentiate between groups, but the original scoring (threshold 2+) was more effective (p = 0.0295). The SSCST modification, awarding points for any number of partners, increased |

| | | | | |
|---|---|---|---|---|
| | | | | referrals but was unnecessary, as most CST cases in study period 3 had fewer than 5 partners or other risk factors.<br><br>**Performance Metrics:**<br>Sensitivity/Specificity of Secondary Questions in Youth with 3+ Positive Items on the SSCST:<br>Sensitivity (95% CI) = 72% (47–90%)<br>Specificity (95% CI) = 98% (91–100%)<br>PPV (95% CI) = 13/14 = 93% (66–100%)<br>NPV (95% CI) = 56/61 = 92% (82–97%). |
| Chisolm-Straker, M. 2021(2) | Rapid Appraisal for Trafficking (RAFT) | Adult patients (4127 ED patients) An emergency department | Validation: criterion validity and construct validity<br>Diagnosis properties: Sensitivity, specificity, ROC area | **Construct Validity**<br>Factor structure: A bifactor model showed the best fit in confirmatory factor analysis (CFA), supporting the hierarchical structure of RAFT.<br>Item selection: Testlet Response Theory confirmed that the four selected items had strong discrimination and covered key trafficking domains (labor exploitation, harm, sexual exploitation, and force).<br>Item consistency: The selected items reflected severe forms of exploitation and performed consistently across samples (NYC and Fort Worth) with minimal differential item functioning (DIF).<br><br>**Criterion Validity**<br>Reference standard: RAFT was validated against the Trafficking Victim Identification Tool (TVIT).<br>Performance metrics:<br>ROC Curve (AUC):<br>All sites: 0.8976<br>New York City: 0.8836<br>Fort Worth: 0.9430<br>Sensitivity and Specificity:<br>Derivation (NYC): 89% sensitivity, 74% specificity<br>Validation (Fort Worth): 100% sensitivity, 61% specificity<br>Combined (any positive RAFT item): 92% sensitivity, 72% specificity<br>Subgroup performance:<br>Sensitivity ranged from 75% to 96% across subgroups by gender, race/ethnicity, language of interview, and age, indicating robust performance in diverse populations.<br>Significant predictors of trafficking (logistic regression):<br>Forced work, threats at work, and payment for sex were statistically significant ($p < 0.01$).<br>Strongest predictor: Payment for sex (Odds Ratio = 24.47). |

| | | | | |
|---|---|---|---|---|
| Hurts, I. A. 2021 (3)<br><br>U.S. | A short, 6-question screening tool -Greenbaum et al. | Adolescents (12-17 years old) / An urban, regional, freestanding children's hospital that serves as a tertiary care level | Diagnosis properties: Sensitivity, specificity, ROC area | **Performance Metrics:**<br>The sensitivity and specificity of the screening tool varied according to the number of items answered affirmatively. With two items answered "yes," sensitivity was 84.6% (95% CI: 70.8–98.5) and specificity was 53.2% (95% CI: 46.1–60.4), with an ROC curve area of 0.6892. With 3 items, sensitivity was 80.8% (95% CI: 65.6–95.9) and specificity increased to 74.7% (95% CI: 68.5–81.0), with an ROC area of 0.7775. For 4 items, sensitivity decreased to 42.3% (95% CI: 23.3–61.3) while specificity increased to 91.4% (95% CI: 87.4–95.4), with an ROC area of 0.6685. Finally, with 5 items, sensitivity was 11.5% (95% CI: 0–23.8) and specificity reached 98.4% (95% CI: 96.6–100), with an ROC area of 0.5496. |
| Kaltiso, S.-A.O. 2021 (4)<br><br>U.S. | Human Trafficking Screening Tool | Adult patients (18 years and older) (26,974 patients were screening) / An urban emergency department | Feasibility and predictive accuracy | **Performance Metrics:**<br>Predictive questions: Eight questions were significantly associated with likely trafficking status, with odds ratios ranging from 1.15 to 7.83.<br>The most predictive question was: "Does anyone make you have any kind of sex for work/money?" (OR = 7.83).<br>AUC: Full 11-question model: AUC = 0.85.Reduced 7-question model: AUC = 0.83 (training set) and 0.82 (validation set).<br>Other Findings<br>False positives: Many of the 147 patients who screened positive but were not confirmed as trafficked had indicators of domestic violence (76 cases or 40.2% of positive screens).<br>Unknown outcomes: 61 patients (32.3%) with positive screens had incomplete or undocumented follow-ups. |
| Greenbaum, V.J. 2018 (5)<br><br>U.S. | A short, 6-question screening tool -Greenbaum et al. | Adolescents 12–18 years / pediatric healthcare facilities (108 participants)<br><br>Atlanta, Georgia, U.S | Diagnosis properties: Sensitivity, specificity and PPV and NPV and ROC area | **Performance Metrics:**<br>Sensitivity: 92% (with ≥2 positive responses).<br>Specificity: 73%.<br>PPV: 51%.<br>NPV: 97%.<br>The tool demonstrated excellent discriminative ability, with an AUC ROC of 0.97.<br><br>**Screening Effectiveness:**<br>A cutoff score of ≥2 positive answers effectively identified CSEC/CST victims. Odds of being a CSEC/CST victim were 22 times higher for those with ≥2 positive responses compared to those with fewer responses**.** |
| Greenbaum, V.J. 2018 (6) | A short, 6-question screening tool -Greenbaum et al. | Adolescents 11–17 years / Five PEDs, six child | Diagnosis properties: Sensitivity, specificity | **Performance Metrics:**<br>The screen had a sensitivity of 84.4% (95% CI: 75.3, 91.2) and specificity of 57.5% (95% CI: 53.8, 61.1) in the total sample; 83.3% sensitivity (95% CI: |

| | | | | |
|---|---|---|---|---|
| U.S. | | advocacy centers, and five teen clinics | | 51.6, 97.9) and 49.4% specificity (95% CI: 37.9, 60.9) in ED settings; 84.0% sensitivity (95% CI: 63.9, 95.5) and 61.4% specificity (95% CI: 56.2, 66.3) in CACs; and 84.9% sensitivity (95% CI: 72.4, 93.3) and 54.6% specificity (95% CI: 48.5, 60.7) in teen clinics. |
| Kaltiso, S.-A.O. 2018 (7)<br><br>U.S. | A short, 6-question screening tool -Greenbaum et al. | Adolescents (12-17 years old) / PED | Diagnosis properties: Sensitivity, specificity | **Performance Metrics:**<br>With a cutoff score of two positive answers the tool demonstrated a 90.9% (95% CI= 58.7%-99.8%) sensitivity, 53.1% (95% CI= 45.6% - 60.4%) specificity, 10.0% (95% CI= 5.0% – 17.6%) PPV, 99.0% (95% CI= 94.7% – 99.9%) NPV, 9.1% (IC 95% = 1.62%–37,7%) false-negative rate, and 46,9% (95% CI= 40.0%–53.9%) false-positive rate. Increase the cutoff value to three positive answers increase the specificity to a 75.5% (95% CI= 68.8%–81,4%) but decreased the sensitivity to ,8 (IC del 95% CI = 58.2%–97.7%). If a positive screen was defined as two positive items plus a positive answer to prior sexual activity, the specificity increased to 64,6% (95% CI = 57.4%–71,3%) while the sensitivity remained the same. |
| Basson, D 2017 (8)<br><br>U.S. | Commercial Sexual Exploitation-Identification Tool (CSE-IT) – version 2.0, WestCoast | Children and youth aged 10 and over (5,537 participants)<br><br>California, U.S | Validation: Criterion Validity, Construct validity, Reliability: | **Criterion Validity:**<br>Concurrent Validity: Established by comparing the CSE-IT with CAT/CSPI on exploitation outcomes, achieving a high correlation (92%). This shows how well the CSE-IT correlates with another measure of the same construct at the same time.<br>Convergent Validity: Demonstrated by comparing CSE-IT outcomes with mental health needs measured by CAT/CSPI. Youth with higher CSE-IT scores also had more severe mental health needs, supporting convergent validity as both tests align on related constructs.<br><br>**Additional Validity Checks:**<br>Face Validity: Verified through focus groups and interviews to ensure the tool appears appropriate for users.<br>Content Validity: Ensured by systematic reviews with experts to confirm it covers relevant information.<br>Utility Validity: Demonstrated by its usability in various service delivery contexts.<br><br>**Structural Validity:**<br>Confirmatory factor analysis: confirmed an 8-factor solution, indicating the tool's structural validity and theoretical coherence.<br><br>**Reliability:**<br>Internal Consistency: Cronbach’s alpha values for sub-scales indicate reliable measurements, with values mostly above 0.7. |

| | | | | |
|---|---|---|---|---|
| Mumma, B. 2017 (9)<br><br>U.S. | Screening survey to identify of victims of sex trafficking (the name of the tool is not mentioned) | 143 Female patients<br>age 18-40 years / A single academic ED<br><br>Sacramento, California, U.S. | Diagnosis properties: Sensitivity, specificity | **Performance Metrics:**<br>Sensitivity: The screening survey demonstrated a sensitivity of 100% (95% CI [74%-100%]), meaning it successfully identified all victims of sex trafficking who participated in the study.<br>Specificity: The survey showed a specificity of 78% (95% CI [70%-85%]), indicating it could accurately identify non-victims most of the time, though with some false positives.<br><br>**Comparison to Physician Concern**:<br>Sensitivity of physician concern: 40% (95% CI [12%-74%]).<br>Specificity of physician concern: 91% (95% CI [85%-95%]).<br>This indicates that while physicians were better at ruling out non-victims, the survey was significantly better at identifying actual victims.<br><br>**Key Screening Question**:<br>All confirmed victims (100%) answered "yes" to one specific question in the survey:<br>"Were you (or anyone you work with) ever beaten, hit, yelled at, raped, threatened, or made to feel physical pain for working slowly or for trying to leave?"<br>This finding suggests that this single question could potentially serve as an effective standalone screening tool. |
| Simich, L. 2014 (10)<br><br>U.S. | Trafficking Victim Identification Tool (TVIT)<br>VERA<br>National Institute of Justice | 180 potential trafficking victims, with additional analysis from 53 administrative case files / victim service organizations<br><br>California, Colorado, New York, Texas, and Washington in the U.S. | Validation: Criterion Validity, Construct validity, Discriminant and Convergent Validity, Predictive Validity Reliability | **Construct Validity:** Factor analysis confirmed that the questions measured key dimensions: abusive labor practices, physical harm, sexual exploitation, isolation, and coercion.<br><br>**Discriminant and Convergent Validity:** The scales effectively distinguished between trafficking victims and victims of other related crimes.<br>Moderate correlations between scales confirmed they measured distinct but related aspects.<br><br>**Criterion Validity:** High agreement between expert classifications and tool results, validating its accuracy in identifying victims.<br><br>**Predictive Validity:** Regression models showed 87% of the questions predicted trafficking cases overall.<br>Specific predictions: Labor trafficking: 71%. Sex trafficking: 81%.<br>The short version (16 questions) performed well with minimal loss of precision.<br><br>**Reliability:** Inter-rater: Consistent results between different evaluators.<br>Internal consistency: High correlations within scales, confirming reliable measurement of key concepts. |

AUC: Area Under the Curve, CAT/CSPI: Crisis Assessment Tool/Childhood Severity of Psychiatric Illness, CI: Confidence Interval, CSE-IT: Commercial Sexual Exploitation-Identification Tool (Tool), CST: Child Sex Trafficking, ED: Emergency Department, NPV: Negative Predictive Value, PED: Pediatric Emergency Department, PPV: Positive Predictive Value, ROC: Receiver Operating Characteristic, SSCST: Short Screen for Child Sex Trafficking, TVIT: Trafficking Victim Identification Tool, U.S: United States

#### Table 2 Toolkits for Identifying and Supporting Survivors of HT

| Reference | Detection focus | Target population | Application context | Content |
|---|---|---|---|---|
| **Adult Human Trafficking Screening Tool and Guide** (11)<br><br>Macias-Konstantopoulos, W. National Human Trafficking Training and Technical Assistance Center (NHTTAC), 2018<br><br>U. S. | HT in general | Adults | General healthcare settings | Toolkit that provides a screening tool to use in identifying adults that may have experienced sex or labor trafficking.<br>This Toolkit includes elements of a trauma-informed screening tool, considerations for administering the tool, agency practices to be implemented before use of the screening tool, ethical and safety considerations, and steps toward appropriate and meaningful referrals.<br><br>1. **Guide by NHTTAC**<br>a. **Before using this toolkit**<br>i. Take HT Training<br>ii. Establish an Internal Response Protocol<br>iii. Implement an Information and Referral Network<br>b. **Administering the Screening Tool**<br>i. Establishing a Relationship<br>ii. Identifying Trafficking Indicators<br>iii. Information and Referral<br>iv. Safety Planning<br>c. **Indicators of HT**<br>i. General Indicators<br>ii. Labor Trafficking<br>iii. Sex Trafficking<br>d. **Agency Considerations for Effective Use**<br>i. HT Policy: Establish a policy following survivor-centered, trauma-informed, and culturally responsive practices.<br>ii. Cross-sector Partnerships: Collaborate with local organizations to meet the complex needs of trafficking victims.<br>iii. Action Plan: Include a safety-oriented plan for positive screening cases, covering confidentiality, reporting, and privacy.<br>iv. Staff Training: Ensure staff are trained in screening administration, mandatory reporting, and recognizing trafficking signs.<br>v. Monitor and evaluate the effectiveness of the training and the impact of the plan of action, and revise procedures as needed for improvement. |

| | | | | |
|---|---|---|---|---|
| | | | | vi. Record Keeping and Confidentiality: Maintaining secure and confidential records.<br>The Toolkit provides Appendix sections with information on how to screen adult victims or those at risk of experiencing trafficking and screening flowcharts for adults at risk.<br><br>**2. Adult Human Trafficking Screening Tool (AHTST) by NHTTAC** |
| **Human Trafficking Response Protocol. A Toolkit for Hospitals**<br><br>Michigan Department of Health and Human Services-Division of Victim Services<br>2020 (12)<br><br>U. S. | HT in general | Applicable to different age groups | Hospital EDs | The aim of the Toolkit is to provide hospitals with a structured framework for identifying, assessing, and responding to HT cases. It equips healthcare professionals with the necessary tools and training to recognize red flags, deliver trauma-informed care, and coordinate with community resources.<br>**1. HT Assessment for Adults**<br>a. Purpose: Helps healthcare providers identify HT victims using a standardized, trauma-informed, and patient-centered approach.<br>b. Structure:<br>i. Screening Questions: Open-ended, non-threatening questions (e.g., "Can you leave your job if you want to?").<br>ii. Red Flags:<br>1. Physical: Trauma signs, malnourishment, STIs, branding/tattoos.<br>2. Behavioral: Fearfulness, avoidance of eye contact, dependency on another person.<br>3. Control: Accompaniment by a controlling individual, lack of personal documents.<br>c. Trauma-Informed Approach: Conducted privately, respecting autonomy, avoiding judgment, and prioritizing emotional well-being.<br>**2. HT Assessment for Children & Adolescents**<br>a. Purpose: Identifies child and adolescent trafficking victims with age-appropriate, non-judgmental screening.<br>b. Structure:<br>i. Screening Questions: Adapted to children's context (e.g., "Do you feel safe at home?").<br>ii. Red Flags:<br>1. Physical: Abuse signs, unexplained STIs, inconsistent injury explanations.<br>2. Behavioral: Withdrawn behavior, scripted stories, excessive loyalty to an older partner.<br>3. Situational: History of running away, presence of a controlling person, reports of sex or labor in exchange for necessities.<br>c. Trauma-Informed Approach: Private interviews, calm/supportive tone, building trust without pressuring disclosure.<br>. |
| **Human Trafficking Toolkit for Healthcare Professionals** (13) | HT in general | Applicable to different age groups | General healthcare settings | This toolkit is designed to support healthcare professionals in the identification, care, and referral of victims and survivors of human trafficking. It promotes a trauma-informed approach and includes practical guidance, validated screening tools, interdisciplinary response strategies, and adaptable protocols for healthcare settings. |

| | | | | |
|---|---|---|---|---|
| United Against Human Trafficking<br><br>U.S. | | | | 1. **Clinical and behavioral indicators** of trafficking in general medical, OB/GYN, and mental health contexts.<br>2. **Trauma-informed care strategies** for building trust, ensuring safety, and responding sensitively without retraumatizing patients.<br>3. **Validated screening tools**, including:<br>a. *Trafficking Victim Identification Tool* (Vera Institute)<br>*b. Commercial Sexual Exploitation Identification Tool (CSE-IT)*<br>*c. Quick Youth Identification Tool*<br>4. **Step-by-step response guidance**, including options for patient-led decisions, hotline use, and referrals—emphasizing full informed consent.<br>5. **Institutional protocol development**, with recommendations on:<br>a. Identifying responsible staff<br>b. Building referral networks<br>c. Differentiating responses for adults and minors<br>d. Complying with HIPAA and reporting laws<br>6. **Culturally competent interpretation**, emphasizing trained interpreters familiar with trauma and trafficking contexts.<br>7. **Connection to local services** (primarily in Houston), with national applicability via referral directories and hotlines. |
| Identifying and Assisting Victims of Human Trafficking (14)<br><br>American Hospital Association<br><br>U.S. | HT in general | Applicable to different age groups | General healthcare settings | This toolkit provides resources for hospital and health care personnel to identify, assess, and support individuals who may be victims of sex or labor trafficking. It includes trauma-informed strategies, recommended practices, training resources, legal guidance, and patient-centered protocols for appropriate response and referral.<br>**1.** **Building Awareness of Human Trafficking Among Staff:** Provides foundational knowledge on sex and labor trafficking, with emphasis on trauma-informed, victim-centered approaches. Includes awareness videos, survivor stories, infographics (e.g., "10 Red Flags"), and multilingual resources.<br>**2. Staff Training for Identifying and Helping Victims of Human Trafficking: Offers webinars and tools to train clinical and non-clinical staff on:**<br>a. Recognizing trafficking indicators<br>b. Using screening tools (e.g., PEARR Tool)<br>c. ICD-10-CM coding for trafficking cases<br>d. Ensuring safe, ethical, and legal responses<br>Also includes models for developing internal protocols and safe clinics.<br>3. **Support Organizations That Offer Help:** Promotes partnerships with national and local organizations such as HEAL Trafficking, SOAR (HHS), Polaris, Blue Campaign, and OVC. Includes referral directories and guidance on creating response networks.<br>**4.** **Tools, Data and Reports:** Provides downloadable tools: red flag checklists, patient materials, clinical flowcharts, and social media toolkits. Includes guidance on data coding, safety planning, and case documentation. Features hospital best practices and implementation examples. |

| | | | | |
|---|---|---|---|---|
| **NHA Human Trafficking Toolkit**<br><br>Nebraska Hospital Association (15)<br><br>U. S. | HT in general | Applicable to different age groups | General healthcare settings | The aim of the Toolkit is to provide healthcare professionals with the knowledge, tools, and protocols necessary to identify, respond to, and intervene in cases of HT.<br>The Toolkit includes<br>**1. Recognizing Warning Signs**<br>a. General signs<br>b. Behavioral indicators<br>c. Physical indicators<br>d. Indicators of labor trafficking<br>e. Indicators of sex trafficking<br>**2. Clinical Decision Trees**<br>a. Provides **structured flowcharts** for identifying and responding to trafficking cases.<br>b. Separate **decision trees for adults and minors** that outline steps for medical intervention, legal reporting, and referral to advocacy services.<br>**3. Screening and Assessment Tools**<br>a. HTI Labs Screening Tool: A digital platform that assists in recognizing trafficking victims.<br>b. PEARR Tool (Provide, Explain, Ask, Respect, Respond): A trauma-informed approach for engaging with suspected victims.<br>c. ICD-10 Billing Codes: Standardized medical billing codes for documenting HT cases.<br>**4. Hospital Policies and Best Practices**<br>a. Examples from Nebraska hospitals on implementing trafficking response protocols.<br>b. Guidelines for developing institutional policies to ensure consistent and effective intervention.<br>**5. Referral Programs and Resources**<br>a. Lists adult and minor referral programs across Nebraska, including domestic violence shelters, advocacy centers, and forensic nursing teams.<br>b. Provides contact details for Nebraska's Child Advocacy Centers for handling minor trafficking cases.<br>c. Shares awareness materials, such as help cards, silent notification tools, and posters for discreet victim support.<br>**6. Next Steps and Call to Action**<br>a. Encourages hospitals to integrate training, policy development, and community collaboration to combat trafficking.<br>b. Provides contact information for state and national trafficking hotlines. |
| **Protocol Toolkit for Developing a Response to Victims of Human** | HT in general | Applicable to different age groups | General healthcare settings | This is a Toolkit designed to help healthcare professionals develop and implement protocols to identify and assist victims of HT in healthcare settings. This document promotes an interdisciplinary approach based on trauma-informed care, offering detailed steps for creating protocols, integrating with existing policies, and continuing staff training. The HEAL Toolkit includes: |

| | | | | |
|---|---|---|---|---|
| **Trafficking in Health Care Settings** (16)<br><br>Baldwin., et al, 2017<br>HEAL Toolkit<br><br>U. S. | | | | 1. **Protocol toolkit for developing a response to victims of HT**<br>a. **Steps for protocol development.**<br>i. Identify community multidisciplinary responders<br>ii. Engage non-medical community stakeholders<br>iii. Engage medical stakeholders<br>iv. Understand HT and health<br>v. Create and convene an interdisciplinary protocol committee<br>vi. Develop a multidisciplinary treatment and referral plan<br>b. **Response protocol for identifying patients at risk for trafficking:**<br>i. Decide who will screen patients and whether the screening will be limited to high-risk patients or include all patients<br>ii. Guidelines for interviewing high-risk patients<br>iii. Strategies for interviewing patients alone<br>iv. Safety considerations<br>v. Multidisciplinary treatment and referral plan<br>vi. Strategies for working with minor patients<br>vii. Responding to patients who decline assistance<br>viii. Documentation procedures<br>ix. Guidelines for forensic examination<br>x. External reporting procedures:<br>c. **Education and training key points related to screening include:**<br>i. Types of specialized training: HT, covering sex and labor trafficking; trauma-informed service delivery to minimize re-traumatization during screening; commercial Sexual Exploitation of Children (CSEC); motivational interviewing to facilitate open communication during screening<br>ii. Basic understanding for all staff: Creating a safe environment for the patient during screening; maintaining cultural competence, confidentiality, and a nonjudgmental attitude; and understanding the general risk factors and potential indicators of trafficking<br>iii. Target departments for training |
| **Toolkit to Combat Trafficking in Persons** (17)<br><br>United Nations Office on Drugs and Crime, 2008<br><br>U.S | HT in general | Applicable to different age groups | General healthcare settings | Toolkit Prevent, Suppress and Punish Trafficking in Persons, Especially Women and Children, supplementing the United Nations Convention against Transnational Organized Crime. The toolkit presents:<br>1. **Indicators of trafficking:**<br>a. General indicators<br>b. Children<br>c. Sexual exploitation<br>d. Labour exploitation<br>e. Domestic servitude<br>f. Begging and petty crime<br>2. **Trafficking indicators card** (United States Immigration and Customs Enforcement): The card prominently displays a hotline for reporting suspicious activity and also provides succinct information on: |

| | | | | |
|---|---|---|---|---|
| | | | | a. The differences between trafficking in persons and smuggling of migrants<br>b. Trafficking indicators<br>**3. Health-care providers' tool for identifying victims**<br>a. Contains suggested screening questions |

AHTST: Adult Human Trafficking Screening Tool, CSEC: Commercial Sexual Exploitation of Children/ Commercially Sexually Exploited Children, ED: Emergency Department, HEAL: Health, Education, Advocacy, and Linkage; HHS: Health and Human Services; HT: Human Trafficking, HTI: Human Trafficking Interview, ICD-10: International Classification of Diseases, NHTTAC: National Human Trafficking Training and Technical Assistance Center, OVC: Office for Victims of Crime; PEARR: Provide privacy, Educate, Ask, Respect, and Respond (Tool); SOAR: Stop, Observe, Ask, Respond; STI: Sexually Transmitted Infection, U.S.: United States.

#### Table 3 Guidance and Protocols for Identifying and Supporting Survivors of HT

| Reference | Scope and purpose | Detection focus | Target population | Intended audience | Application context | Methods | Content |
|---|---|---|---|---|---|---|---|
| **Assessing for Human Trafficking**<br><br>Human Trafficking Collaborative University of Michigan<br>2021 (18)<br><br>U. S. | This document provides a comprehensive framework for healthcare providers to screen and assess patients for potential HT situations. It outlines logistics for screening, including who should be screened, who should conduct the screening, appropriate times for screening, and the importance of ensuring a safe environment during the process. | HT in general | Applicable to different age groups | Healthcare providers, including doctors, podiatrists, dentists, chiropractors, clinical psychologists, optometrists, nurse practitioners, nurse-midwives, and clinical social workers. | General healthcare settings | Guidance<br>Non-systematic review | It focuses on 3 themes:<br>**1. Screening Logistics**: Guidelines on who to screen, who should conduct the screening, when to screen, and how to ensure a safe environment for screening.<br><br>**2. Clinical Indicators**: Lists of general indicators, as well as those specific to labor and sex trafficking, to assist healthcare providers in identifying potential trafficking victims.<br><br>**3. Screening Questions**: Specific questions designed to elicit information about potential trafficking situations.<br>- General Life Situation:<br>- Can you leave your job or living situation when you want to?<br>- Has anyone threatened you or your family if you tried to leave?<br><br>- Work Conditions and Compensation:<br>- Are you being paid fairly for your work?<br>- Has your employer taken your passport, ID, or other documents?<br>- Personal Freedom and Control:<br>- Do you feel safe where you live and work? |

| | | | | | | | |
|---|---|---|---|---|---|---|---|
| | | | | | | | - Has anyone forced you to do something you didn't want to do?<br>These questions are designed to be asked in a private setting, ensuring that no one else is present during the interview. |
| **Building a Child Welfare Response to Child Trafficking Handbook**<br><br>The Center for the Human Rights of Children at Loyola University Chicago 2011 (19)<br><br>U.S. | To equip state and private child welfare agencies with the necessary tools, policies, and best practices to identify and respond to child trafficking cases effectively. It provides screening tools, case management strategies, legal protections, and referral resources to ensure child victims receive appropriate care and services. | HT in general | Children and adolescents | Law enforcement, legal professionals, healthcare providers, and educators. | Illinois Department of Children and Family Services (IDCFS) | Report | It consists of four components:<br>**1. Identification and Investigation**<br>a. Defines child trafficking and distinguishes it from human smuggling.<br>b. Provides indicators for detecting sex and labor trafficking of minors.<br>c. Includes interview guidelines for law enforcement and caseworkers.<br>**2. Introduces screening tools:**<br>a. Rapid Screening Tool (RST) for quick victim identification.<br>**b.** Comprehensive Screening and Safety Tool (CSST) for risk assessment**.**<br>**3. Case Management**<br>a. Provides caseworker templates:<br>i. New Client Checklist<br>ii. Child-Trafficking Informed Consent Form<br>iii. Tripartite Assessment for evaluating needs<br>**b.** Covers best practices for mental health, safety planning, and legal support**.**<br>**4. Legal Protections and Advocacy**<br>a. Explains federal and state laws relevant to child trafficking.<br>b. Outlines immigration options for foreign-born victims (e.g., T visas).<br>c. Discusses victims' rights in the legal system, including access to restitution.<br>**5. Resources and Referrals**<br>a. Lists national and local organizations offering assistance.<br>b. Includes contacts for law enforcement, shelters, health services, and advocacy groups.<br>**6. Integrating Trafficking Response into Child Welfare** |

| | | | | | | | |
|---|---|---|---|---|---|---|---|
| | | | | | | | a. Highlights the Illinois Safe Children Act (2010), which:<br>i. Decriminalized juvenile prostitution.<br>ii. Ensured child sex trafficking victims are treated as victims, not criminals.<br>b. Demonstrates how child protection services can apply trafficking response protocols. |
| **Caring for Trafficked Persons**<br><br>International Organization for Migration (IOM)<br>2009 (20)<br><br>Switzerland | This document is a guidance for Health Providers offers practical, non-clinical guidance for healthcare professionals to understand HT, recognize its health consequences, and provide ethical, trauma-informed, and culturally appropriate care. It highlights the physical, psychological, and social health risks of trafficking and includes 17 action sheets covering key areas such as trauma-informed care, health assessments, mental health support, sexual and reproductive health, patient confidentiality, and safe referrals. | HT in general | Applicable to different age groups | Health care professionals | General healthcare settings | Guidance<br>Non-systematic review | The guide consists of the following components**:**<br>**1. Understanding HT and Its Health Impacts**<br>a. Explanation of forms of exploitation: sexual trafficking, forced labor, servitude, organ trafficking.<br>b. Key health consequences: injuries, infectious diseases, malnutrition, mental health issues (PTSD, anxiety, depression).<br>**2. Guiding Principles for Healthcare Providers**<br>a. Do no harm and ensure patient safety.<br>b. Confidentiality in handling patient information.<br>c. Trauma-informed approach to avoid re-traumatization.<br>d. Respect for cultural diversity and effective communication with interpreters.<br>e. Informed consent in all medical interventions.<br>**3. Practical Care Guidelines**<br>a. Comprehensive health assessment addressing physical, psychological, and sexual health needs.<br>b. Mental health care, including trauma, depression, and PTSD support.<br>c. Sexual and reproductive health services, such as STI treatment, contraception, and care for sexual violence survivors.<br>d. Specialized care for children and vulnerable populations**.**<br>**4. Ethical and Legal Considerations** |

| | | | | | | | |
|---|---|---|---|---|---|---|---|
| | | | | | | | a. Patient privacy and data protection.<br>b. Safe referrals to shelters, legal aid, and support services.<br>The guide also includes:<br>**1. Flowchart for Victim Identification and Response**<br>a. Guides healthcare providers through **key screening steps**.<br>b. Covers **confidentiality, informed consent, and referral pathways**.<br>c. Helps assess risk and determine **next steps for victim support**.<br>**2. Indicators of HT**<br>a. **Physical signs**: Injuries, malnutrition, poor hygiene, occupational hazards.<br>b. **Psychological signs**: PTSD, depression, anxiety, dissociation.<br>c. **Behavioral signs**: Scripted responses, avoidance of authorities, fear.<br>d. **Situational signs**: Lack of personal documents, restricted movement, excessive working hours, debt bondage. |
| **Child Sex Trafficking and Commercial Sexual Exploitation: Health Care Needs of Victims**<br><br>Greenbaum, J Committee on Child Abuse and Neglect, 2013–2014, 2015 (21)<br><br>U.S. | This document addresses the critical health care needs of victims of child sex trafficking and commercial sexual exploitation. It provides valuable insights and guidelines for healthcare professionals to better support and treat these vulnerable individuals. | Sex trafficking | Children and Adolescents | Health care professionals | General healthcare settings | Guidance Non-systematic review | This article provides:<br>1. **Risk Factors for CSEC**<br>a. Age of Vulnerability<br>b. High-Risk Populations<br>c. Environmental and Societal Factors:<br>**2. Victim Identification and Evaluation**<br>a. Healthcare Settings.<br>b. Common Medical Issues<br>c. Challenges in Identification<br>**3. Indicators of HT**<br>a. Behavioral Indicators<br>b. Physical Indicators<br>c. Situational Indicators<br>**4. Questions to Ask Potential Victims**<br>a. Exchange of Sex for Needs<br>b. Involvement with Others<br>c. Sexual Imagery<br>**5. Recommendations** |

| | | | | | | | |
|---|---|---|---|---|---|---|---|
| | | | | | | | a. Evaluation Approach:<br>Building Rapport<br>Safety and Confidentiality Trauma-Informed Care<br>Medical and Diagnostic Focus<br>6. Conclusions and Guidance |
| **Child sexual exploitation: improving recognition and response in health settings**<br><br>Academy of Medical Royal Colleges, 2014 (22)<br><br>UK | The purpose of this report is to make recommendations to the medical Royal Colleges and Faculties to help:<br>• Raise professional awareness of the indicators of sexual exploitation.<br>• Support health care professionals in communicating with and engaging young people in this situation.<br>• Ensure that health care professionals feel equipped to refer sexually exploited children in a safe and appropriate manner to local services for assistance. | Sex trafficking | Children and adolescents suspected of or at risk of victimization by sexual exploitation (up to 18 years of age) | Health care professionals | General healthcare settings | Guidance: The working group included representation from a number of medical colleges and schools and received input from other disciplines and health groups | The guide references and includes tools for identifying victims of sexual exploitation<br>**1. Vulnerability factors for CSE**<br>2. **Possible warning signs** of CSE (drawn from CCSEGG interim report, 2012)<br>**3. Identifying child sexual exploitation in health settings**<br>a. PHYSICAL HEALTH presentations of CSE<br>4. **Implementation strategies**<br>Five key components were identified to improving the response of health professionals to child sexual exploitation:<br>• Training<br>• Awareness<br>• Recognition<br>• Response<br>• Supervision and support<br>5. **Recommendations:**<br>Curricula Review<br>Generic Capabilities Review-Caldicott Principles<br>Local Safeguarding Protocols-Multi-Agency Approaches<br>New Partnerships<br>Support and Supervision<br>Public Health Collaboration<br>Quality Improvement Review |
| **Commercial Sexual Exploitation of Children (CSEC) Protocol for San Luis Obispo County** | Outlines the policies, procedures, and collaborative efforts of various agencies to address and support youth | Sex trafficking | Children | All Community Partners that directly work with youth, including Emergency Medical | Multiple settings | Protocol | San Luis Obispo County Commercial Sexual Exploitation of Children (CSEC) Protocol<br>1. **Medical services responses** designed to support victims of commercial sexual exploitation include:<br>a. Child Welfare Services (CWS)<br>b. Suspected Abuse Response Team (SART) |

| | | | | | | | |
|---|---|---|---|---|---|---|---|
| CSEC Collaborative Response Team, 2017 (23)<br><br>U.S. | who are victims of commercial sexual exploitation.<br>The protocol aims to place CSE youth in protective environments that offer trauma-specific therapeutic programming to stabilize them during this critical time. | | | Services/ Paramedic, Mental Health, Nurses and Doctors, Public Health | | | c. RISE<br>d. San Luis Obispo County Behavioral Health<br>2. **County Policy:** A victim-centered approach aims to help sexually exploited youth heal and transition into adulthood. The protocol focuses on placing these youth in protective environments with therapeutic programming tailored to their trauma.<br>3. **Governance:** The Children's Services Network (CSN) and a Multi-Agency Steering Committee oversee the implementation of this protocol, ensuring ongoing collaboration and effectiveness through regular reviews.<br>4. **Memorandum of Understanding (MOU):** The MOU highlights the collaborative efforts between law enforcement, service providers, and the community to address HT and support victims.<br>5. **Information Sharing and Confidentiality**<br>6. **Training and Prevention**<br>7. **Identification and Risk Factors**: The document provides tools and guidelines for screening and assessing youth at risk of or currently experiencing exploitation, with specific risk factors identified.<br>8. **Intervention:** Detailed procedures for multidisciplinary team meetings and immediate response protocols are outlined to ensure timely and coordinated support for the youth.<br>9. **Ongoing Support** |
| **Commercially sexually exploited children (CSEC): Screening tool user guide.**<br><br>CSEC Collaborative Response Team, 2015 (24) | Guide to interpret each of the tool items | Sex trafficking | Children | Not reported | Multiple settings | User manual | It serves as a manual explaining how to use, interpret, and apply the tool effectively.<br>**Content:**<br>1. Provides structured guidelines for identifying at-risk minors.<br>2. Offers example questions for interviews.<br>3. Outlines key risk indicators related to CSEC. |

| | | | | | | | |
|---|---|---|---|---|---|---|---|
| U.S. | | | | | | | 4. Explains a victim-centered, trauma-informed approach.<br>5. Emphasizes the importance of confidentiality and appropriate referrals.<br>**Application:**<br>1. Guides professionals in healthcare, social services, and juvenile justice on integrating the tool into their assessments.<br>2. Promotes inter-agency collaboration for a coordinated response. |
| **Committee on Health Care for Underserved Women**<br><br>Committee on Health Care for Underserved Women<br>2024 (25)<br><br>U.S. | To provide guidance for healthcare professionals, particularly obstetricians and gynecologists, on how to identify, respond to, and assist victims of HT. | HT in general | Applicable to different age groups | Healthcare professionals | General healthcare settings | Report | The guide includes:<br>1. **Types of trafficking.**<br>2. **At-risk populations**.<br>3. **Key indicators:**<br>a. Physical – Injuries, malnutrition, signs of abuse.<br>b. Behavioral – Fear, avoidance, scripted responses.<br>c. Situational – No control over documents, unpaid labor, accompanied by a controlling person.<br>4. **Screening questions:** Assess work conditions, personal freedom, coercion, and safety (must be done privately).<br>5. **Healthcare role:** trauma-informed approach.<br>6. **Resources & next steps**:<br>a. National Human Trafficking Hotline for referrals.<br>b. Display discreet resource materials in clinics.<br>c. Encourage staff training for better victim identification. |
| **Domestic Minor Sex Trafficking Intervene Identifying and Responding to America's Prostituted Youth** | This guide focuses on DMST, offering tools and training to identify, support, and rehabilitate victims. It explains the dynamics of trafficking, | HT in general | Applicable to different age groups | Health care professionals | Child Advocacy Centers (CACs). Social Services and Mental Health Settings. Community Organizations | Guidance | The guide consists of:<br>**Section I – The Practitioner Guide**<br>The Practitioner Guide trains first responders on DMST, helping them recognize victimization patterns and respond effectively.<br><br>**Section II – The Intake Tool** |

| | | | | | | | |
|---|---|---|---|---|---|---|---|
| Shared Hope International, 2010 (26)<br><br>U.S. | including recruitment tactics, methods of control, and the severe psychological, physical, and emotional impacts on victims. The guide emphasizes accurate identification to prevent mislabeling and provides a trauma-informed, strengths-based approach for practitioners. | | | | | | A trauma-informed, strengths-based system designed to identify DMST victims while minimizing re-traumatization. It structures invasive questions carefully and includes follow-up questions to maintain emotional stability and improve recall. Not all sections are meant for every practitioner.<br><br>The Intake Tool aims to effectively identify potential victims of DMST through a trauma-informed and strengths-based approach, minimizing the risk of re-traumatization.<br>**Implementation Methodology**:<br>Two-Tier System:<br>- Tier 1: Consists of preliminary questions to quickly identify signs or indicators of trafficking. This level is designed to be used by any professional who may have initial contact with a victim.<br>- Tier 2: Comprises a deeper and more detailed assessment intended for professionals with specialized training in trafficking intervention, such as social workers, psychologists, or healthcare personnel.<br>**Trauma-Informed and Strengths-Based Approach:**<br>Questions are structured to avoid directly triggering traumatic memories.<br>When addressing sensitive topics, questions focus on the victim's strengths or positive aspects to help restore emotional stability |
| **Framework for a Human Trafficking Protocol in Healthcare Settings**<br><br>National Human Trafficking Resource Center (NHTRC)<br>2010 (27)<br><br>U.S. | No information | HT in general | Applicable to different age groups | Healthcare providers | General healthcare settings | Protocol | Components of the Protocol:<br>1. **Identification of Red Flags**<br>a. Lists common behavioral, physical, and situational indicators of trafficking.<br>b. Helps healthcare providers detect signs of coercion, medical neglect, and psychological distress.<br>2. **Engagement and Safety**<br>a. Emphasizes immediate intervention in high-risk situations.<br>b. Recommends the use of professional interpreters to prevent interference from traffickers. |

| | | | | | | | |
|---|---|---|---|---|---|---|---|
| | | | | | | | c. Guides providers on discussing available options with the patient while maintaining confidentiality.<br>3. **Referral and Collaboration**<br>a. Advises connecting victims with local social services and community resources.<br>b. Encourages partnerships with anti-trafficking organizations and crisis response teams for complex cases.<br>4. **Long-Term Planning**<br>a. Recommends the development of hospital-specific protocols to standardize trafficking responses.<br>b. Suggests compiling a resource list of support services to facilitate rapid assistance for victims. |
| **Guia para desenvolver uma resposta às vítimas de tráfi co de pessoas em ambientes de assistência à saúd**<br><br>HEAL Trafficking 2023 (28)<br><br>Brazil | A multidisciplinary protocol for identifying and assisting trafficking victims (an adaptation from the manual developed by the NGO HEAL Trafficking) | HT in general | Applicable to different age groups | Health care professionals | General healthcare settings | Guidance Adapted to Brazil based on the manual developed by HEAL Trafficking. The guide's development involved collaboration with experts in HT and representatives from security, justice, and migrant protection sectors. | The protocol's components as outlined in the guide include:<br><br>1. Process of Identification<br>2. Qualified Listening<br>3. Safety Considerations<br>4. Multidisciplinary Follow-up and Referral Plan<br>5. Training and Capacity Building<br>6. Integration with Social Services<br>7. Monitoring and Evaluation |
| **Hiding in Plain Sight**<br><br>Hughes, DM<br>2003 (29)<br><br>U.S. | This guide focuses on identifying victims of sexual trafficking as defined by the Trafficking Victims Protection Act of | Sex trafficking | Applicable to different age groups | Social workers and healthcare providers, | General healthcare settings | Narrative review | Components of the Guide<br>1. **Definition of Trafficking**<br>a. Sex trafficking<br>b. Severe forms of trafficking<br>c. Distinction between foreign-born and domestic victims<br>2. **Indicators of HT** |

| | | | | | | | |
|---|---|---|---|---|---|---|---|
| | 2000 (TVPA). It provides practical strategies for recognizing and assisting victims, with a particular emphasis on those trafficked for commercial sex acts. | | | | | | a. Examples of force<br>b. Indications of force<br>c. Examples of coercion<br>d. Examples of fraud<br>3. **Where and How to Find Victims**<br>a. Commercial sex venues<br>b. Public advertisements<br>c. Emergency healthcare settings<br>d. Immigrant communities<br>e. Areas with transient male populations<br>4. **Who Should Report Cases**<br>a. Mandated reporters<br>b. Hotlines & contacts<br>c. Challenges in law enforcement. |
| **Human Trafficking: Care and Response** (30)<br><br>Virginia Hospital & Healthcare Association / Boyett, R.<br><br>U. S. | This guide aims to train healthcare professionals to identify, respond to, and care for victims of HT through a trauma-informed and survivor-centered approach, providing tools, best practices, protocols, and resources to ensure their safety and access to services, while promoting awareness and collaboration within healthcare systems. | HT in general | Applicable to different age groups | Health care professionals | General healthcare settings | Guidance Non-systematic review | Components of the Guide<br>1. **Human Trafficking Task Force**: Overview of the task force's mission and role.<br>2. **Executive Summary & Key Points**: Introduction to the issue and key takeaways for healthcare professionals.<br>3. **Defining HT**: Explanation of trafficking types, including sex and labor trafficking.<br>4. **Scope of the Issue**: Prevalence and impact of trafficking.<br>5. **Child & Adult Trafficking**: Distinctions and unique vulnerabilities of each group.<br>6. **Trafficking Mechanisms**: How traffickers recruit, control, and exploit victims.<br>7. **Indicators in Healthcare Settings**:<br>a. Red flags<br>b. Medical signs<br>c. Behavioral indicators of trafficking.<br>8. **Myths vs. Facts**: Addressing common misconceptions about HT.<br>9. **Buyers & Perpetrators**: Profiles of those who exploit trafficking survivors.<br>10. **Health Outcomes**: Physical and psychological consequences of trafficking.<br>11. **Identification & Response Tools**: |

| | | | | | | | |
|---|---|---|---|---|---|---|---|
| | | | | | | | a. Guidelines for screening<br>b. Interviewing<br>c. Responding to trafficking cases.<br>12. **Mandatory Reporting Laws**: Legal obligations for healthcare professionals in Virginia.<br>13. **Resources & Training**: Additional materials, training programs, and support networks for professionals. |
| **Human Trafficking: Guidebook on Identification, Assessment, and Response in the Health Care Setting** (31)<br><br>The Massachusetts General Hospital (MGH) Human Trafficking Initiative and the Massachusetts Medical Society Committee on Violence Intervention and Prevention / Alpert, E. J.<br><br>U.S. | The objective of this guide is to educate healthcare providers about HT and to provide resources for patient referral and ongoing professional education. It aims to enhance healthcare professionals' ability to identify, assess, and respond to HT cases, while contributing to global efforts in intervention and prevention. The guide offers “red flags” and questions for identifying potential victims, and references existing tools like the Vera Institute and Ohio Task Force screening tools, but clarifies that these tools lack | HT in general | Applicable to different age groups | Health care professionals | General healthcare settings | Guidance Non-systematic review | Components of the Guide<br>1. **Definitions and Types:** Explains forms of trafficking (sexual, labor, organ) and distinguishes trafficking from smuggling.<br>2. **Health Effects**: Describes physical, mental, reproductive, and developmental consequences.<br>3. **Assessment:** Provides "red flags," guidelines for medical history, physical examination, and documentation.<br>4. **Victim Care:** Focuses on disclosure, safety planning, and case management.<br>5. **Legal Aspects**: Addresses legal protection and immigration options for victims.<br>6. **Collaboration:** Promotes interdisciplinary work with external organizations.<br>7. **Self-Care and Resources**: Offers strategies for healthcare providers' well-being and useful references. |

| | | | | | | | |
|---|---|---|---|---|---|---|---|
| | sufficient evidence of effectiveness. | | | | | | |
| **Human Trafficking: A Guide to Identification and Approach for the Emergency Physician**<br><br>Shandro, J, 2016 (32)<br><br>U.S. | This article outlines the clinical approach to the identification and treatment of a potential victim of HT in the ED. | HT in general | Applicable to different age groups | Emergency practitioners | Emergency settings | Guidance<br>Non-systematic review | Components of the Guide<br>1. **Red flags** and signs that indicate a patient in the ED may be a victim of HT.<br>2. **Interviewing Tips**<br>3. **Key components of ED and institutional protocols** for caring for trafficking survivors.<br>4. **List of clinical indicators** that should arouse suspicion for trafficking<br>5. **Clinical priorities** for caring for a victim of trafficking<br>Institutional contacts (social work, forensic examiners, security) to help care for a victim of trafficking<br>6. **Institutional security plan** for identified victims of trafficking<br>7. **Local mandatory reporting laws**<br>8. **Local HT resources** for referral<br>9. **Local forensic examiner information** and guidelines for referral<br>10. **Local and** national law enforcement contact information and guidelines for referral<br>11. **Author recommendations** |
| **Human Trafficking: Clinical Assessment Guideline**<br><br>Leslie, J, 2018 (33)<br><br>U.S. | Review the HT victim identification process for health care settings. | Sex or labor trafficking | Applicable to different age groups | HCPs | Hospital EDs | Guidance<br>Non-systematic review | Components of the Guide<br>1. **Warning signs** from other publications<br>a. Physical warning signs<br>b. Other warning signs<br>c. Mental health<br>2. **Screening questions** from other publications<br>3. **Follow-Up Questions** to Ask About Living/Work Conditions from other publications<br>4. **Physical Screening** from other publications<br>5. **14-question survey screening tool** by Mumma et al., 2017<br>6. **National Human Trafficking Resource Center (NHTRC)**<br>a. Indicators of labor trafficking<br>b. Indicators of sex trafficking |

| | | | | | | | |
|---|---|---|---|---|---|---|---|
| | | | | | | | c. NHTRC's Recommendations for Assessments<br>7. **Recommendation by the author** |
| **Human Trafficking Response Program Shared Learnings Manual**<br><br>Dignity Health<br>2019 (34)<br><br>U.S. | The PEARR Tool is a key component of Dignity Health's Abuse, Neglect, and Violence policy.<br>Dignity Health developed the PEARR Tool, in partnership with HEAL Trafficking and Pacific Survivor Center, to help guide social workers, nurses, chaplains, and other health professionals on how to provide victim assistance to patients in a trauma-informed manner. | HT in general | Applicable to different age groups | Healthcare professionals | General healthcare settings | Report | Components of the Approach<br>1. **Privacy and Confidentiality**<br>a. Prioritizes ensuring a safe environment for the patient by minimizing external influence.<br>b. Encourages private conversations to allow victims to disclose information without fear.<br>2. **Education and Awareness**<br>a. Recommends providing general information on HT to all patients.<br>b. Promotes a non-stigmatizing approach by integrating trafficking awareness into routine care discussions.<br>3. **Screening and Inquiry**<br>a. Advocates for using open-ended, neutral questions to explore potential trafficking situations.<br>b. Advises avoiding pressure or confrontation to ensure patient comfort and autonomy.<br>4. **Respect and Response**<br>a. Emphasizes respecting the patient's decisions, even if they choose not to seek help immediately.<br>b. Encourages trauma-informed interventions, including referrals to shelters, legal aid, and healthcare services. |
| **Identification of Victims/ Persons 'At-Risk' of Trafficking in Human Beings**<br><br>UNICEF, 2022 (35)<br><br>Switzerland | Practical Guide for Frontline Responders/<br>General/<br>can be used as a practical tool with information on what Trafficking in Human Beings is, | HT in general | Cases of child trafficking/ children 'at-risk' of trafficking | Frontline Responders Border and immigration officers;<br>Front-line police officials;<br>Child protection authorities; | Multiple settings | Guidance Methods not reported | Components of the Guide<br>List several strategies:<br>1. **International and National Legal Framework on THB**<br>2. **Indicators** of HT<br>3. **Special indicators** for Border Police (and other relevant stakeholders) that a child may have been trafficked/'at-risk' of being trafficked |

| | | | | | | | |
|---|---|---|---|---|---|---|---|
| | with a list of indicators, and recommended questions for interviewing trafficked persons/ those 'at-risk' of trafficking.<br>As training material on identification of trafficked victims/ persons who may be 'at-risk' of trafficking.<br>As a concise guide for child-friendly communication with practical techniques and facilitators. | | | Social workers;<br>g Staff of Blue Dots Hubs;<br>Other relevant government and non-government service providers | | | 4. **Some specific indicators** and guiding questions for all frontline responders<br>a. Recruitment and Migration Experience<br>b. Means of Control used by Traffickers<br>5. **Key Processes and Guidelines** for Identifying and Communicating with Trafficking Victims<br>**a. Chronology of the Identification Process**<br>**b. Informed Consent and Legal Protections**<br>c. **Initial Screening Interview**: Before the Interview; During the Interview; and Referrals and Further Steps.<br>6. **Techniques for Child-Friendly Communication**<br>a. Best Interest of the Child<br>b. Child-Sensitive Approach<br>c. Victim-Centric Approach<br>d. Practical Techniques |
| **Identifying and supporting victims of modern slavery: guidance for health staff**<br><br>Department of Health & Social Care, 2015 (36)<br><br>UK | Identifying and supporting victims of modern slavery: guidance for health staff produced by the Department of Health / Modern slavery (HT) / It is not a detection tool, it provides indicators for suspected trafficking and general recommendations. | HT in general | Applicable to different age groups | Health Staff | General healthcare settings | Guidance Methods not reported | Components of the Guide<br>1. **Signs of trafficking**<br>2. **Possible health care issues**<br>3. **Recommendations** including:<br>a. Suspect that a person is a victim<br>b. Ask further questions and get additional information and support<br>c. What you should do next<br>d. Communication to possible victims<br>e. Referral |
| **Identifying Victims of Human Trafficking: What to Look for in a Healthcare Setting** | List potential red flags and indicators that medical providers may see in a patient who | HT in general | Not reported | Anyone in a healthcare setting – from clerical staff to lab technicians, nursing staff, | General healthcare settings | Report. | Components of the Guide<br>1. **Red Flags and Indicators**<br>a. General Indicators of HT<br>b. Labor Trafficking Indicators<br>c. Sex Trafficking Indicators |

| | | | | | | | |
|---|---|---|---|---|---|---|---|
| National Human Trafficking Resource Center, 2016 (37)<br><br>U.S. | may be a victim of HT. | | | ambulatory care, radiology staff, security personnel, case managers, and physicians. | | | d. Health Indicators and Consequences of HT<br>e. Physical Health Indicators<br>f. Mental Health Indicators<br>g. Social or Developmental Indicators<br>2. **Conducting Assessments with Potential Victims**<br>3. **Key Recommendations** |
| **London Safeguarding Trafficked Children Guidance**<br><br>London Safeguarding Children's Board 2011 (38)<br><br>UK | Provides guidance to professionals and volunteers from all agencies in safeguarding and promoting the welfare of trafficked and exploited children. This guidance is linked to the London Safeguarding Trafficked Children Toolkit 2011. | HT in general | Children and adolescents | Professionals and volunteers from all agencies | Multiple settings | Guidance supplementary to, the London Safeguarding Children Board's London Child Protection Procedures. Methods not reported | Components of the Guide<br>1. **Introduction:** Overview of the purpose and importance of safeguarding trafficked children.<br>2. **Definitions:** Explanation of key terms, including HT and child exploitation**.**<br>3. **Principles:** Core principles guiding agencies in identifying and responding to trafficked children**.**<br>4. **Problem of Child Trafficking:** Reasons, recruitment methods, control mechanisms, and impacts on children.<br>5. **Identification of Trafficked Children:** Indicators, obstacles to self-identification, and professional responsibilities.<br>a. Physical<br>b. Behavioral signs<br>c. Situational signs<br>6. **Common Assessment Framework (CAF):** Use of structured assessment to evaluate risk and needs.<br>a. A structured tool to evaluate the child's situation, risks, and needs.<br>b. Encourages inter-agency collaboration to ensure a comprehensive assessment.<br>7. **Children at Risk of Harm:** Referral processes and local authority response mechanisms**.**<br>8. **National Referral Mechanism (NRM):** Multi-stage process for identifying and assisting trafficked children.<br>9. **Working with Trafficked Children:** Best practices, including legal, medical, and psychological support. |

| | | | | | | | |
|---|---|---|---|---|---|---|---|
| | | | | | | | 10. **Vulnerable Groups:** Special considerations for missing children and those in care**.**<br>11. **Information Sharing**: Importance of inter-agency cooperation to ensure child protection**.**<br>**Role of Local Safeguarding Boards:** Responsibilities in coordinating efforts to protect trafficked children |
| **Practice Guide for Intake and Investigative Response to Human Trafficking of Children**<br><br>Connecticut Department of Children and Families, 2015 (39)<br><br>U. S. | The tool is designed to guide the identification, assessment, and response to child trafficking cases in Connecticut, U.S. Provides comprehensive protocols for intake, medical and mental health evaluations, safety planning, and service coordination. | HT in general | Children and adolescents who are victims or at risk of HT | Professionals and stakeholders directly involved in responding to and managing cases of child trafficking (Social workers, HART (Human Anti-Trafficking Response Team) liaisons, Law enforcement agencies, Healthcare professionals, Community service providers, Legal and immigration professionals) | Multiple settings | Methods not reported | Components of the Guide<br>1. **Human Trafficking Screening Tool:** Used by social workers and healthcare providers to identify victims or those at risk of trafficking.<br>2. **Decision Maps:** Guides to determine risk levels (confirmed victim, high risk, or at risk) and appropriate interventions.<br>3. **Medical Assessment Protocols:**<br>a. Physical, sexual, substance use, and dental health evaluations.<br>b. Trauma-sensitive procedures for assessment and treatment.<br>4. **Behavioral Health Assessments:**<br>a. Trauma evaluations and mental health assessments.<br>b. Development of safety and treatment plans, including therapy.<br>c. Safety Planning: Tailored plans addressing risks like running away, re-encounters with traffickers, and online safety.<br>5. **Support Service Coordination:**<br>a. Access to advocacy, mentoring, legal services, and community-based programs.<br>b. Job training and educational consultations.<br>6. **Monitoring and Follow-Up:**<br>a. 90-day case monitoring by HART liaisons.<br>b. Use of the Human Trafficking Monitoring Tool for progress tracking. |

| | | | | | | | |
|---|---|---|---|---|---|---|---|
| | | | | | | | 7. **Training and Education:** Training sessions for professionals and community education campaigns. |
| **Recognizing Child Trafficking as a Critical Emerging Health Threat** (40)<br><br>Peck et al., 2021<br><br>U.S. | This white paper reframes child trafficking from a criminal justice issue to a pressing public health crisis. It is aimed at equipping pediatric healthcare providers (HCPs) with evidence-based, trauma-informed, and culturally responsive strategies to identify, respond to, and prevent child trafficking in clinical settings. | Sex or labor trafficking | Children and adolescents who are victims or at risk of HT | Pediatric healthcare professionals, including pediatricians, family medicine physicians, nurses, nurse practitioners, physician assistants, social workers, mental health providers, and emergency department staff who care for minors. It is also relevant for medical trainees and educators involved in pediatric or adolescent health. | Pediatrich healthcare settings | Methods not reported | The guide presents:<br>**Conceptual Framework:**<br>Positions child trafficking as a public health issue, not just a criminal justice matter. Addresses both sex and labor trafficking of minors. Highlights the healthcare system as a critical intervention point for unidentified victims.<br>1. **Identified Risk Factors:**<br>a. History of childhood trauma, neglect, or abuse.<br>b. Involvement in the foster care system.<br>c. LGBTQI+ identity.<br>d. Economic instability or forced migration.<br>2. **Health Impacts:**<br>a. Physical: STIs, unwanted pregnancy, injuries, substance use.<br>b. Mental: PTSD, depression, suicidal ideation, dissociation.<br>3. **Common Clinical Presentations:**<br>a. Frequent visits for unresolved or vague health issues.<br>b. Controlling companions, reluctance to speak, lack of documents.<br>c. Inconsistent stories or signs of fear and distress.<br>4. **Clinical Recommendations:**<br>a. Use a trauma-informed, victim-centered approach.<br>b. Ask open-ended, nonjudgmental questions.<br>c. Ensure confidentiality and obtain informed consent.<br>d. Engage culturally competent, professional interpreters.<br>5. **Practical Tools Included:**<br>a. Sample open-ended screening questions.<br>b. Risk indicator checklists for pediatric environments. |

| | | | | | | | |
|---|---|---|---|---|---|---|---|
| | | | | | | | c. Tables outlining trafficking warning signs in clinical settings.<br>**6. Calls to Action:**<br>a. **Individual level:** Ongoing training for providers.<br>b. **Organizational level:** Development of clear screening and response protocols.<br>c. **Systemic level:** Investment in research, policy development, and multisector coordination. |
| **Resource Guide on Human Trafficking for Title X Family Planning Clinics** (41)<br><br>HHS Office of Population Affairs (OPA) Title X Family Planning Program<br><br>U.S. | This guide, developed by the U.S. Office of Population Affairs (OPA), is tailored for Title X family planning clinics to enhance the identification, care, and referral of potential victims of human trafficking, particularly sex trafficking, within reproductive health settings | HT in general | Applicable to different age groups | Healthcare providers working in Title X-funded reproductive health settings, including physicians, nurses, medical assistants, and social workers. | Reproductive health settings | Guidance Methods not reported | The guide is organized into eight sections:<br><br>1. **Basics of Human Trafficking:** Defines key concepts (e.g., AMP model), legal definitions, myths, risk factors, and indicators of trafficking, with a focus on healthcare contexts.<br>2. **Core Tools for Providers:** Outlines guiding principles, survivor-informed approaches, screening tools, victim needs assessments, and intersections with homelessness, domestic violence, and mandatory reporting.<br>3. **Victim-Centered, Trauma-Informed Care:** Describes care models, therapeutic interventions, safety planning, and HIPAA considerations when working with victims.<br>4. **Staff Care:** Emphasizes cultural competence, unconscious bias, and prevention of clinician burnout and vicarious trauma.<br>5. **Community Connections:** Offers guidance on collaborating with community and faith-based organizations, task forces, and legal/law enforcement partners, including rural and Indigenous communities.<br>6. **Federal Resources:** Lists anti-trafficking programs and tools from agencies like HHS, DOJ, State, and others.<br>7. **Awareness Materials:** Provides posters, brochures, infographics, and other printable outreach materials. |

| | | | | | | | |
|---|---|---|---|---|---|---|---|
| | | | | | | | 8. **Hotlines:** Includes contact information for national and federal hotlines addressing human trafficking, runaway youth, and labor violations. |
| **Reference Guide on Protecting the Rights of Child Victims of Trafficking in Europe**<br><br>UNICEF, 2006 (42)<br><br>Switzerland | The Guidelines focus on the steps needed to protect and assist anyone under 18 who is believed to have been trafficked, and to make decisions about their future. Serves as an implementation book for the Guidelines as it gives information about the steps and procedures that constitute 'good practice' in the protection and assistance of child victims of trafficking. As such, the Guide is a practical tool for policy makers and practitioners from government, non-governmental and international organizations responsible for protecting and assisting child victims of trafficking across Europe. Endorsed by the Stability Pact Task Force on | HT in general | Children: Anyone under 18 who is believed to have been trafficked | Policy makers and practitioners from government, non-governmental and international organizations responsible for protecting and assisting child victims of trafficking across Europe | Multiple settings | Guidance Reference guide In 2003, UNICEF created "Guidelines for Protection of the Rights of Child Victims of Trafficking in South Eastern Europe." These Guidelines, endorsed by the Stability Pact Task Force, are based on international standards. | The UNICEF Guidelines cover 11 specific aspects concerning trafficking of children:<br>1. Identification of children as victims of trafficking.<br>2. Appointment of a guardian for each trafficked child.<br>3. Questioning by the authorities.<br>4. Referral to appropriate services and inter-agency coordination.<br>5. Interim care and protection.<br>6. Regularization of a child's status in a country other than their own.<br>7. Individual case assessment and identification of a durable solution;<br>8. Implementing a durable solution, e.g., possible return to a child's country of origin.<br>9. Access for children to justice.<br>10. Protection of the child as a victim and potential witness; and<br>11. Training for government and other agencies dealing with child victims.<br><br>Includes the **"Guidelines for Conducting Forensic Interviews with Child Victims of Trafficking**"<br>1. General principles<br>a. Pro-active identification measures<br>b. Presumption of age<br>2. Who is responsible for taking action?<br>3. Guidelines for Conducting Forensic Interviews with Child Victims of Trafficking<br>a. Professionals Involved<br>b. Key Objectives of Interviews<br>i. Identify the Child<br>ii. Determine Legal Status<br>iii. Understand the Child's Experience<br>iv. Gather Evidence of Crimes |

| | | | | | | | |
|---|---|---|---|---|---|---|---|
| | Trafficking for Southeastern Europe and adopted by member states (2003). | | | | | | v. Support the Child's Emotional Well-being<br>c. Principles for Interviewing Trafficked Children<br>d. Special Considerations Trauma Awareness<br>e. Practical Interview Techniques<br>f. Types of Questions to Ask<br>i. General Open-Ended Questions<br>ii. Specific Questions (Adapted for Age and Circumstances):<br>iii. Recruitment<br>iv. Migration<br>v. Working Conditions<br>g. Tools and Materials<br>h. Indicators of HT<br>i. Physical and Behavioral Signs<br>ii. Situational Indicators |
| **Safeguarding Children and Young People from Sexual Exploitation in Leicester, Leicestershire and Rutland (LLR)**<br><br>Safeguarding Children Board (LSCB) of Leicester, Leicestershire, and Rutland, 2016 (43)<br><br>UK | This guidance provides a framework for preventing, identifying, and addressing child sexual exploitation (CSE) in Leicester, Leicestershire, and Rutland, UK. It emphasizes multi-agency collaboration to safeguard children under 18 through prevention strategies, early identification of risks, tailored interventions, and prosecution of perpetrators. The document prioritizes treating | Sex trafficking | Children and young people under the age of 18 who are at risk of, or affected by, sexual exploitation, including cases involving grooming, coercion, trafficking (both domestic and international), and other forms of exploitation linked to sex trafficking. | All professionals and agencies involved in safeguarding children, including law enforcement (police), children's social care workers, health services (nurses, GPs, mental health professionals), education staff (teachers, school leaders), voluntary agencies, and other organizations working with children and families. | Multiple settings | Guidance Methods not reported | The guide highlights the importance of early identification, prevention, and intervention in addressing child sexual exploitation (CSE), emphasizing that a multi-agency collaboration is essential for an effective response.<br>Included Tools and Strategies:<br>1. **Risk Factors & Warning Signs**<br>a. Grooming: Establishing control over a child by giving them gifts, money, affection, or manipulating their trust.<br>b. Vulnerabilities: Poverty, social exclusion, prior abuse history, low self-esteem, being in care. Family history of substance misuse or domestic violence.<br>c. Warning Signs:<br>i. Secretive behavior, staying out late, associating with older individuals.<br>ii. Unexplained money, mobile phones, or expensive items.<br>iii. Missing school or being frequently absent. |

| | | | | | | | |
|---|---|---|---|---|---|---|---|
| | sexually exploited children as victims, not criminals, and focuses on disrupting exploitation while ensuring their protection and recovery. | | | | | | iv. Multiple mobile phones, unexplained injuries, involvement in petty crime.<br>d. High-Risk Indicators:<br>i. Frequent missing episodes.<br>ii. Older, controlling partner.<br>iii. Entering unknown vehicles.<br>iv. Signs of physical/emotional abuse.<br>2. **Multi-Agency Approach:**<br>Collaboration is key: Various agencies must work together (social care, police, health services, education, voluntary sector).<br>3. **Managing Cases**<br>a. Strategy Discussion: A multi-agency review of cases to assess risk and plan intervention.<br>b. Multi-Agency CSE Meetings: Convened for high/moderate-risk cases.<br>c. Team Around the Child (TAC/TAF).<br>d. Confidentiality and Information Sharing.<br>4. **Risk Assessment & Intervention**<br>a. Risk Assessment Tool: Classifies children into At Risk, Medium Risk, and High Risk categories.<br>b. Intervention Pathway:<br>i. At Risk: Prevention through education and family involvement.<br>ii. Medium Risk: Multi-agency safety plans, disruption tactics.<br>iii. High Risk: Immediate action needed, police investigation, possible prosecution.<br>c. Working with Victims:<br>i. Requires long-term intervention and trusted relationships.<br>ii. Support networks should be built around the child. |

| | | | | | | | |
|---|---|---|---|---|---|---|---|
| | | | | | | | iii. Care-takers and professionals should monitor risky behavior (e.g., social media use, phone activity, visitors).<br>5. **Disrupting & Prosecuting Perpetrators**<br>a. Identifying Perpetrators.<br>b. Disrupting Perpetrator Behavior<br>c. Gathering Evidence<br>d. Victim & Witness Support<br>Also provide: CSE Risk Assessment Tool, Social Care Flowchart, Guidelines for Young People Over 18. |
| **Safeguarding children who may be trafficked**<br><br>Gweithdrefnau Diogelu Cymru Wales Safeguarding Procedures 2021 (last update) (44)<br><br>UK | This practice guide provides additional information about safeguarding responses when a child may have been trafficked. It should be used in conjunction with the Wales Safeguarding Procedures. There are some issues which are common across safeguarding practice guides and some which are specific to the safeguarding issue being considered. | HT in general | Children and adolescents | Practitioners working with children (up to the age of 18). This includes those working in early years, social care, education, health, the police, youth offending and youth, community and family support services (including the third sector) and foster care and residential care. | Multiple settings | Guidance Methods not reported | Components of the Guide<br>1. **Indicators**<br>a. Things that children may say and ways they may behave<br>b. Physical indicators of exploitation<br>c. Indicators related to the movement of a child from one place to another<br>d. Indicators of exploitation and control by another person<br>e. Indicators related to documentation and personal details<br>f. Further risk indicators<br>g. Indicators to be alert to in health settings<br>2. **Approaches to care** with which these cases should be addressed. It also explains relevant aspects of HT and explains the particularities of child trafficking in the UK. It provides guidance on how to make referrals to other services and post-identification care. |
| **Safeguarding children who may have been trafficked; Practice guidance** | Non-statutory good practice guidance for agencies in England which are likely to encounter, or have referred to them, children and | HT in general | Children and adolescents who may have been trafficked | Practice nurse, health visitors, hospital staff, maternity staff, adult mental health and child and adolescent | Multiple settings | Guidance Methods not reported | Contains recommendations including:<br>1. **Role of specific agencies and services** including health services<br>**2. Identifying trafficked children**<br>3. **Possible indicators** that a child may have been trafficked<br>a. At port of entry |

| | | | | | | | |
|---|---|---|---|---|---|---|---|
| Department for education, HM Government, 2011 (45)<br><br>UK | young people who may have been trafficked. It is intended to help agencies safeguard and promote the welfare of children who may have been trafficked. | | | mental health services practitioners | | | b. Whilst resident in the UK (in addition to those listed above)<br>4. **Specific action during an initial assessment** |
| **Screening for Human Trafficking Guidelines for Administering the Trafficking Victim Identification Tool (TVIT)**<br><br>Vera Institute of Justice 2014 (46)<br><br>U.S. | It consists of screening questions that help identify victims of HT and ensure they receive necessary protection and services. It is noted that the tool should complement, not replace, specialized training and professional practices. Negative responses do not rule out victimization, as trauma can affect responses. | HT in general | Applicable to different age groups | Victim service agency staff and other social service providers, who will administer the Trafficking Victim Identification Tool (TVIT). Law enforcement, health care and shelter workers will also find it helpful in improving trafficking victim identification | Multiple settings | User manual | Components of the guide<br>1. **Purpose & Scope:** Provides guidelines for administering the Trafficking Victim Identification Tool (TVIT) and best practices for identifying trafficking survivors.<br>2. **Interviewing Techniques:** Includes recommendations on building trust, ensuring confidentiality, and understanding trauma effects.<br>3. **Considerations for Law Enforcement**: Emphasizes a victim-centered approach and coordination with support services.<br>4. **Trafficking Victim Identification Tool (TVIT):** Contains both long and short versions of the screening tool for assessing potential trafficking situations.<br>5. **Frequently Asked Questions**: Clarifies common concerns about screening, identifying victims, and using the tool effectively.<br>6. **Training & Resources**: Lists additional materials, training opportunities, and legal frameworks related to HT.<br>7. **Definitions & Legal Context**: Outlines U.S. legal definitions of HT and federal protections available to survivors. |
| **Screening to Identify Commercially Sexually Exploited Children; A Guide for Implementing the Commercial Sexual Exploitation –** | Guide that provides recommendations for implementing the Commercial Sexual Exploitation–Identification Tool | Sex trafficking | Children and youth aged 10 and older, regardless of gender, ethnicity, culture, sexual | Staff with a variety of professional backgrounds including HCPs | Multiple child-serving systems, including child welfare, juvenile justice, schools, residential, mental health, medical, | Guide of a tool. The WestCoast Children's Clinic developed the Commercial Sexual Exploitation – | The guide presents:<br>1. **Commercial Sexual Exploitation–Identification Tool (CSE-IT) by WestCoast Children's Clinic**<br>a. Domains and questions (1. Housing and caregiving. 2. Prior abuse or trauma. 3. Physical health and appearance. 4. |

| | | | | | | | |
|---|---|---|---|---|---|---|---|
| **Identification Tool (CSE-IT) in Youth-Serving Organizations**<br><br>Haley, Basson, & Lang. WestCoast Children's Clinic, 2017 (47)<br><br>U.S. | (CSE-IT) to improve early identification of commercially sexually exploited children. | | orientation, health status, socioeconomic background, or behavior. Screening should also be extended to individuals outside the recommended age range if there are signs of exploitation or risk factors. | | and homeless services | Identification Tool (CSE-IT) in 2014. | Environment and exposure. 5. Relationships and personal belongings. 6. Signs of current trauma. 7. Coercion. 8. Exploitation)<br>b. Scoring Instructions<br>c. Guide for Implementing the Commercial Sexual Exploitation – Identification Tool<br>2. **Developing a Screening and Response Protocol**<br>a. Key Components of the Protocol<br>b. Response to Trafficking Indicators<br>c. Documentation<br>d. Training and Technical Assistance<br>e. Implementation and Continuous Improvement |
| **Spotting the Signs – CSE proforma**<br><br>Rogstad, K. (The Faculty of Forensic & Legal Medicine) 2014 (48)<br><br>UK | Spotting the Signs, funded by the Department of Health, allows sexual health professionals to use a standardized approach to pick up on the warning signs of CSE in all its forms. It is designed to be integrated into existing sexual and social history taking frameworks. Spotting the Signs provides a framework to support conversations with young people around CSE linked to latest research and evidence bases. | Sex trafficking | Children and Adolescents | Sexual health professionals | Sexual health services | Report.<br>The proforma was written by Dr Karen Rogstad of BASHH and Georgia Johnston of Brook, and was developed with the support of a multi-agency advisory board and working group, including focus groups of young people from across the UK. The proforma was piloted in a range of services including GUM clinics, specialist young people's services and General Practice. | The guidance provides questions to help practitioners identify a young person's circumstances or behaviours that may be cause for concern and indicate the young person's needs.<br><br>Three resources are available on the website for identifying potential victims of sexual exploitation.<br>1. Spotting the Signs – CSE proforma A4<br>2. Spotting the Signs – CSE proforma A3<br>3. Spotting the Signs – foldout leaflet<br><br>The tool has the following screening sections:<br>1. Education<br>2. Family Relationships<br>3. Friendships<br>4. Relationships<br>5. Consent<br>6. Sexual Health<br>7. Professional analysis<br><br>And presents the results of a pilot test to evaluate its usefulness and use in 23 institutions. |

| | | | | | | | |
|---|---|---|---|---|---|---|---|
| **The Anatomy of Human Trafficking: Learning About the Blues: A Healthcare Provider's Guide**<br><br>Stevens, M. 2016 (49)<br><br>U.S. | This article explores the scope of the problem, definitions, types, and elements of HT. The roles of clinicians, particularly ED nurses and advanced practice nurses, in screening and identifying those at risk are examined. | Sex or labor trafficking | Not specified | Clinician, including ED, primary care, and forensic nurses as well as advanced practice nurses | General healthcare settings | Narrative review | This document consists of a narrative review that provides indicators and recommendations for the detection of HT. With regard to detection, it indicates:<br>1. **Red Flags for HT**<br>a. Physical<br>b. Environment<br>c. Poor Physical Health<br>d. Psychological<br>e. Poor Mental Health or Abnormal Behavior<br>f. Lack of Control<br>2. **Screening Tools:**<br>The Trafficking Victim Identification Tool (TVIT) by the Vera Institute of Justice |
| **The Commercial Sexual Exploitation of Children: The Medical Provider's Role in Identification, Assessment, and Treatment**<br><br>American Professional Society on the Abuse of Children (APSAC) 2013 (50)<br><br>U.S. | These guidelines provide medical professionals with an overview regarding the current understanding of the commercial sexual exploitation of children. | Sex trafficking | Children and adolescents (under 18 years of age) victims of commercial sexual exploitation | Medical Providers | General healthcare settings | Guidance Methods not reported | 1. **Possible Indicators of CSEC**<br>2. **Signs** that child is being controlled (domineering person accompanying child)<br>3. **Questions** for CSEC identification<br>4. **Tips for Interviewing** CSEC Patients<br>5. **Commercially sexually exploited children (CSEC) screening procedure guideline**<br>a. A list of Potential Questions for Medical Interview of Possible CSEC Victim<br>i. Reproductive Health<br>ii. Prior Sexual Victimization<br>iii. Prior Violence<br>iv. Prior Inflicted Injuries<br>v. Other injury:<br>vi. Anogenital trauma:<br>vii. Prior Confinement<br>viii. Emotional Health<br>ix. Drugs and Alcohol including CRAFFT Screen (Center for Adolescent Substance Abuse Research)<br>x. Additional Questions |
| **The IOM Handbook on Direct Assistance for Victims of Trafficking**<br><br>International Organization for | Provides guidance and advice necessary to effectively deliver a full range of assistance to | HT in general | Applicable to different age groups | Not specified if health care professionals but is intended for all instances involving the | Not specified | Guidance Methods not reported | Chapter 2 Screening of Victims of Trafficking: Presents a formula to enable organizations to better distinguish between the different crimes of trafficking in human beings and people smuggling and outlines a methodology for the screening and identification of individuals seeking assistance as trafficking victims. |

| | | | | | | | |
|---|---|---|---|---|---|---|---|
| Migration (IOM), 2007 (51)<br><br>Switzerland | victims of trafficking from the point of initial contact and screening up to the effective social reintegration of the individuals concerned. | | | referral of trafficking victims to service delivery organizations including health authorities | | | 1. **Limitations of the Screening Process** and key points to consider<br>2. **Key Indicators**<br>3. **Recommendations** including:<br>a. Screening interview and special considerations for interviewing minors<br>b. Victim Response and Treatment<br>c. Using the Screening Interview Form<br>Also provides appendix sections with:<br>1. **Ethical Principles** in Interviewing and Caring for Trafficked Persons<br>2. **Interview Checklist**<br>a. Conditions<br>b. Explanation<br>c. Final Points Before Beginning the Interview<br>d. Key Interview Questions:<br>i. Recruitment Phase<br>ii. Transportation Phase<br>iii. Exploitation Phase<br>iv. Decision-Making |
| **The London Child Sexual Exploitation Operating Protocol**<br><br>London Safeguarding Children Board, 2015 (52)<br><br>UK | The protocol provides a structured multi-agency response to protect children from sexual exploitation. It defines strategies for identification, intervention, prevention and investigation to safeguard children at risk. It also establishes a framework for law enforcement, social services, health care and community organizations to | Sex trafficking | Children and young people under the age of 18 who are at risk of, or affected by, sexual exploitation, including cases involving grooming, coercion, trafficking (both domestic and international), and other forms of exploitation | Social workers, healthcare providers, law enforcement, and community partners. | Multiple settings | Protocol. Methods not reported | The protocol includes:<br>1. **Identifying and Challenging CSE**<br>a. Uses the acronym SAFEGUARD to recognize warning signs.<br>b. Daily contact between police and other agencies to identify cases.<br>c. Multi-agency meetings ensure intelligence-sharing and coordinated responses.<br>2. **Reporting Suspicions of CSE**<br>a. Procedures for documenting CSE suspicions in police databases.<br>b. Categorization of investigations based on risk levels.<br>3. **Support for Victims and Families**<br>a. CSE affects not only the victim but also their family. |

| | | | | | | | |
|---|---|---|---|---|---|---|---|
| | effectively coordinate responses. | | linked to sex trafficking. | | | | b. Collaboration with organizations like Barnardo's and PACE (Parents against Child Exploitation).<br>c. Overview of coercion methods used by perpetrators.<br>4. **Prevention Strategies**<br>a. Identifies high-risk groups.<br>b. Education and awareness campaigns in schools and communities.<br>c. Mapping of high-risk areas and training businesses in the nighttime economy (Operation Makesafe).<br>5. **Intervention Strategies**<br>a. Strategies to reduce the vulnerability of at-risk children.<br>b. Promotion of positive family relationships and preventive sex education.<br>c. Risk assessments through health and child welfare agencies.<br>6. **Disruption Strategies**<br>a. Use of Child Abduction Warning Notices and legal measures against perpetrators.<br>b. Patrolling of "hotspot" locations where exploitation occurs.<br>c. Surveillance of criminal networks using technology and police intelligence.<br>7. **Investigation Strategies**<br>a. Proactive investigations to dismantle exploitation networks.<br>b. Use of recorded interviews to collect evidence.<br>c. Application of the National Referral Mechanism for child trafficking cases.<br>8. **Outcomes Framework for CSE**<br>a. Success evaluation based on: Reduction in child disappearances. Increased |

| | | | | | | | |
|---|---|---|---|---|---|---|---|
| | | | | | | | prosecution rates of perpetrators.<br>Improved access to support services.<br>9. **Communication**<br>a. Strategies to enhance public awareness of CSE.<br>b. Collaboration with the media to report cases sensitively.<br>c. Use of social media to reach potential victims. |
| **The PEARR Tool: Trauma-informed approach to victim assistance in health care settings.**<br><br>Common Spirit Health, HEAL Trafficking, Pacific Survivor Center, 2019 (53)<br><br>U.S. | A trauma-informed approach designed to assist healthcare professionals in providing care to patients who may be affected by abuse, neglect, or violence, such as HT. | HT in general | Applicable to different age groups | Healthcare professionals | General healthcare settings | User manual | The scope includes guidelines on how to educate, ask, respect, and respond to patients in a culturally and developmentally sensitive manner. It provides strategies for maintaining privacy, educating patients, asking the right questions, and respecting their choices while ensuring their safety and connecting them with appropriate resources.<br>**The tool is structured around four key steps:**<br>1. Provide Privacy<br>2. Educate<br>3. Ask<br>a. General Inquiry<br>b. Safety Concerns<br>4. Respect & Respond<br><br>Also provides:<br>1. **Risk Factors** for HT<br>2. **Potential Indicators** of Victimization<br>3. **Resources for Assistance** |
| **The Role of the Nurse in Combating Human Trafficking** (54)<br><br>Sabella, 2011<br><br>U.S | This article aims to raise awareness among nurses about human trafficking, helping them recognize potential victims in clinical settings and respond safely and effectively. It outlines key | HT in general | Applicable to different age groups | Healthcare professionals | General healthcare settings | Guidance Non-systematic review | This article provides an overview of human trafficking, describes how to recognize signs that a person is being trafficked and how to safely intervene, and offers a resource list.<br>1. Clinical Case: Story of a trafficking victim whose signs were missed in the ED.<br>2. Definitions and Types of Trafficking: Sex trafficking, labor trafficking, debt bondage, child trafficking, sex tourism. |

| | | | | | | | |
|---|---|---|---|---|---|---|---|
| | indicators of trafficking, provides guidance on safe intervention, and emphasizes the critical role nurses play in identifying and supporting victims. | | | | | | 3. Prevalence and Risk Factors: Global and U.S. scope; poverty, gender inequality, and false promises as drivers.<br>4. Legal Framework: Overview of the Trafficking Victims Protection Act (TVPA) and victim services.<br>5. Health Impacts: Common physical and psychological consequences for trafficked individuals.<br>6. Barriers to Identification: Victims' fear, language barriers, coercion, and lack of awareness.<br>7. Red Flags in Clinical Settings: Behavioral and physical indicators; presence of controlling companions; inconsistencies.<br>8. Recommended Clinical Responses: Private interviews, neutral interpreters, trauma-informed questions, referral to NHTRC.<br>9. Role of Nurses and Educators: Need for training, policy development, and curricular integration.<br>10. Resources and Follow-up: List of support organizations; update on Elis's case and recovery. |

CACs: Child Advocacy Centers, CAF: Common Assessment Framework, CCSEGG: Child Sexual Exploitation in Gangs and Groups, CRAFFT: C (Car) – Riding with someone under the influence, R (Relax) – Using substances to relax or fit in, A (Alone) – Using substances alone, F (Forget) – Memory loss while using, F (Family/Friends) – Being advised to cut down; CSE: Child Sexual Exploitation/ Commercially Sexually Exploited, CSEC: Commercial Sexual Exploitation of Children/ Commercially Sexually Exploited Children, CSE-IT: Commercial Sexual Exploitation-Identification Tool, CSN: Children's Services Network, CSST: Comprehensive Screening and Safety Tool, CWS: Child Welfare Services, DMST: Domestic Minor Sex Trafficking, ED: Emergency Department, GUM: Genitourinary Medicine, HART: Human Anti-Trafficking Response Team, HEAL: Health, Education, Advocacy, and Linkage, HT: Human Trafficking, IDCFS: Illinois Department of Children and Family Services, IOM: International Organization for Migration, LLR: Leicestershire and Rutland, LSCB: Safeguarding Children Board of Leicester, MOU: Memorandum of Understanding, NGO: Non-Governmental Organization, NHTRC: National Human Trafficking Resource Center, NRM: National Referral Mechanism, OPA: Office of Population Affair,  PACE: Parents Against Child Exploitation, PEARR: Provide privacy, Educate, Ask, Respect, and Respond (Tool), PTSD: Post-Traumatic Stress Disorder, RICE:RST: Rapid Screening Tool, SART: Suspected Abuse Response Team, STI: Sexually Transmitted Infection, TAC/TAF: THB: Trafficking in Human Beings, TVIT: Trafficking Victim Identification Tool, TVPA: Trafficking Victims Protection Act, UNICEF: United Nations Children's Fund, UK: United Kingdom, U.S.: United States.

### Table 4 Indicators of HT

| Category | Subcategory | Types of indicators | Sexual Trafficking | Labour Trafficking | Other types of trafficking | General trafficking | Children and adolescents | Adults | Studies where the indicator is exposed |
|---|---|---|---|---|---|---|---|---|---|
| Physical Indicators | General | Appearance-Related Indicators (loss of hair, tattoos or other forms of "branding," Severe weight loss, malnutrition and dehydration, skin problems, dental/oral problems) | | | | X | X | X | (13,25,35,37,40,54–61) |
| | | Cardiovascular/respiratory conditions that appear to be caused or worsened by stress, such as: Arrhythmia, High blood pressure, Acute Respiratory Distress | | | | X | X | X | (9,31,51,62) |
| | | Neurological conditions: Traumatic brain injury, Headaches or migraines, Unexplained memory loss, Vertigo of unknown etiology, Insomnia, Difficulty concentrating | | | | X | X | X | (13,18,37) |
| | | Injuries or impairments typical of certain jobs or control measures | | X | | | X | X | (17,40,44) |
| | | Gastrointestinal conditions that appear to be caused or worsened by stress, such as: Constipation, Irritable bowel syndrome | | | | X | X | X | (13,18,37) |
| | | Physical abuse, drugs/alcohol | | | | X | X | X | (13,39,40) |
| | | Communicable and non-communicable diseases (e.g. TB, hepatitis) | | | | X | X | X | (37,62) |
| | | Other health issues: Effects of prolonged exposure to extreme temperatures, Effects of prolonged exposure to industrial or agricultural chemicals, Somatic complaints | | | | X | X | X | (18,37) |

| | | | | | | | | | |
|---|---|---|---|---|---|---|---|---|---|
| | Sexual | Physical and sexual abuse (vaginal / anal pain, Chronic pelvic pain) | X | | | | X | X | (13,38,40,44,61,62) |
| | | Presence of STIs or symptoms of STIs | X | | | | X | X | (13,37,38,44,53,58–61) |
| | | Pregnancy indicators (first medical consultation after 24 weeks pregnant), unintended pregnancy, and multiple pregnancies or miscarriages | X | | | | X | X | (37,44,45,48,59,60,62) |
| | | Reproductive issues: Genitourinary issues, Forced or pressured abortions, Genital trauma, Sexual dysfunction, Retained foreign body | X | | | | X | X | (37,40) |
| | Labour | Physical indications of working (For example overly tired in school, indications of manual labour – condition of hands/skin, backaches) | | X | | | X | | (44) |
| | | Occupational-type injuries or physical ailments linked to their work | | X | | | X | X | (55,63) |
| | Indicators of medical or physical neglect | Untreated medical conditions | | | | X | X | X | (42,53–56,63) |
| | | Unexplained or untreated illnesses | | | | X | X | | (42,54) |
| | | Signs of poor hygiene | | | | X | X | | (13,53,54,62) |
| | | Malnourished | | | | X | X | X | (13,38,40,44,54) |
| | | Patient does not have health insurance and/or pays with cash | | | | X | X | X | (2,13,17,44,53,60) |
| | Indicators of exploitation and | Be subjected to violence or threats of violence against themselves or | | | | X | X | X | (17) |

| | | | | | | | | | |
|---|---|---|---|---|---|---|---|---|---|
| | control by another person | against their family members or loved ones | | | | | | | |
| | | Patient was threatened with a weapon | | | | X | X | X | (59) |
| **Mental Health Indicators and Behavioral signs** | General | Self-harm, including attempted suicide or Suicidal ideation, Self-harming behaviors | | | | | X | X | (37,44,45,62) |
| | | Signs of depression, or anxiety | | | | | X | X | (13,35,37,42,44,54,60,62) |
| | | Non-specific post-traumatic stress disorder or Psychological – indications of trauma or numbing | | | | | X | | (37,44,55,60,62,63) |
| | | Low cognitive functioning and concerns about thought processes | | | | | X | X | (59) |
| | | Intrusive Symptoms: Nightmares, Flashbacks. Emotional and Behavioral Responses: Lack of emotional responsiveness, feelings of shame or guilt, hostility. Attachment and Social Difficulties: Attachment disorders, withdrawal, fear, sadness. Dissociative Symptoms: Depersonalization, derealization, dissociative disorders, distortions in perception of time, detachment from self or reality. | | | | | X | X | (37,62) |
| | Behavioral Sings | Show fear or anxiety | | | | | X | X | (13,37,54,62) |
| | | Submissive, tense, nervous/paranoid or Hypervigilant | | | | | X | X | (13,17,19,32,35,37,42,48,53,55–58,60,63) |
| | | Irritable/aggressive behaviour | | | | | X | X | (18,40,53,55–58,63) |
| | | Patient appears to be “in crisis” or continual crying | | | | | X | X | (21,33,66,79,81,83,88,99) |
| | | Poor concentration or memory, unsociable behaviour | | | | X | X | | (37,62) |

| | | | | | | | | | |
|---|---|---|---|---|---|---|---|---|---|
| | | Uncooperative behavior (like resistant to assistance or questions) | | | | X | X | X | (59,60) |
| | | Avoiding eye contact | | | | X | X | X | (44) |
| | | Exhibits self-assurance, maturity and self-confidence not expected in a child of such age | | | | | X | | (32,53,57,62) |
| | | Fearful of employer or supervisor | | X | | | | X | (25,37,57–60,62) |
| | | Patient was in a hurry/expressed they had been at the ED too long | | | | X | X | X | (44,61,64) |
| | Sexual | Mental, physical and sexual trauma | X | | | | X | X | (11) |
| | | Shows promiscuity | X | | | | X | | (59) |
| Situational Indicators | General | Entering or leaving vehicles driven by unknown adults | | | | X | X | | (44) |
| | | Adults loitering outside the child's usual place of residence | | | | X | X | | (55,63) |
| | | Persistently missing, staying out overnight or returning home late with no plausible explanation | | | | X | X | | (38,44) |
| | | Reports from reliable sources suggest likelihood of sexual exploitation, including being seen in places known to be used for sexual exploitation | X | | | | X | | (38,44) |
| | | Come from a place known to be a source of human trafficking | | | | X | X | | (38,40,44) |
| | | Is one among a number of unrelated children found at one address | | | | X | X | | (38,44) |
| | | Being found in exploitative settings such as mines, sweatshops, brothels, or places associated with illegal activities strongly indicates trafficking due to their connection with exploitation (3,88) | | X | | | | X | (17,35) |

| | | | | | | | | | |
|---|---|---|---|---|---|---|---|---|---|
| | | Is living or working in a location with high security measures (e.g. opaque or boarded-up windows, bars on windows, barbed wire, security cameras, etc.) | | X | | | | | (17,51) |
| | | Abnormal work hours; no breaks or vacations | | X | | | | X | (56) |
| | | Was recruited through false promises concerning the nature and conditions of their work | | X | | | X | X | (11) |
| | | Unusual hours/regular patterns of child leaving or returning to the residence which indicates probable HT | | X | | | X | | (56) |
| | | Child returning afterhaving been missing, looking well cared for despite no known base | | | | X | X | | (38,44) |
| | | Claims to have been in the country for years but hasn't learnt the local language(s) or culture | | | | X | X | X | (38,44) |
| | | Not knowing what country they are in or the location of house where live | | | | X | X | X | (44,54) |
| | | Is not aware of their location, the current date, or time | | | | X | X | X | (35) |
| | Related to the movement from one place to other | Be reported missing by their employer even though they are still living in their employer's house | | | X | | | X | (36,54,59,65) |
| | | Travel in groups with persons who are not relatives | | | | X | X | X | (17) |
| | | Moves frequently or talk about travelling to other locations | | | | X | X | X | (9) |
| | | Changing their migration story, evasiveness, denial, minimizing the situation, telling exactly the same story as other migrants from the same area or | | | | X | X | X | (55,63) |

| | | | | | | | | | |
|---|---|---|---|---|---|---|---|---|---|
| | | sharing an inconsistent and scripted history | | | | | | | |
| | | Patient was not local to the area | | | | X | X | X | (35) |
| | | Accompanying adult previously made multiple visa applications for other children/ acted as the guarantor for other children’s visa applications | | | | | X | | (59) |
| | Indicators related to documentation | Not in possession of their passport or other travel or identity documents, as those documents are being held by someone else | | | | X | X | X | (38,40,44,54) |
| | | Irregularity with migration status (entered country illegally, inconsistencies in the description of the migratory journey, visa arranged by someone other than themselvesor their family) | | | | X | X | X | (17,32,35,37,56,57,61) |
| | | False documentation or genuine documentation that has been altered or fraudulently obtained | | | | X | X | X | (17,61) |
| | | The child says they have a different name or other personal details than those in the passport | | | | | X | | (38,44) |
| | Personal Indicators | Patient reports illicit drug use or substance abuse (Note: maybe forced) | | | | X | X | X | (35) |
| | | Has money, expensive clothes, mobile phones or other possessions without plausible explanation | | | | X | X | X | (37,59,60,62) |
| | | Offering to pay cash, advance cash payments | | | | X | | X | (38,44,54) |
| | Sexual Signs | High number of sexual partners | X | | | | X | X | (57) |
| | | Patient history of physical- or sexual-abuse victimization | X | | | | X | X | (58,60,62) |
| | | Significantly older partner | X | | | | X | | (60) |
| | | Evidence of controlling or dominating relationships | X | | | | X | X | (38,44,60) |

| | | | | | | | | | |
|---|---|---|---|---|---|---|---|---|---|
| | | (excessive concerns about pleasing a family member, romantic partner, or employer) | | | | | | | |
| | Behavioral Signs | Acts as if they were instructed by someone else or the person appears to be under the control and supervision of someone who never leaves the person alone. | | | | | X | X | (33,54) |
| | | Reacts with unusually fearful or anxious behavior at any reference to “law enforcement” | | | | X | X | X | (17,44) |
| | | Refused to speak with/distrust of law enforcement | | | | X | X | X | (33) |
| | | Is afraid of revealing their immigration status/ deportation | | | | | X | X | (17,59) |
| | | Looks intimidated and behaves in a way that does not correspond with behaviour typical of children their age | | | | | X | | (17,61) |
| | | Patient uses terminology indicative of sex work (e.g., pimp, escort, player, the life, turn out) | X | | | | X | X | (17,35) |
| | | Often runs away | | | | | X | | (60) |
| | Social Indicators | Has no friends of their own age outside of work | | X | | | X | | (55,63) |
| | | Lives as gang members, with adults who are not their parents | | | X | | X | | (17) |
| | | Lives with members of their gang | | | X | | | X | (17) |
| | | There is evidence that suspected victims have been involved in begging or in committing petty crimes in another country | | | X | | | | (17) |
| | | Impaired social skills | | | | | X | X | (17) |
| | | Social isolation | | | | | X | | (37) |
| | Domestic Servitude | Has no private space | | | X | | | X | (66) (101) |
| | | Sleeps in a shared or inappropriate space | | | X | | | X | (17) |

| | | | | | | | | | |
|---|---|---|---|---|---|---|---|---|---|
| | | Is only given leftovers to eat | | | X | | | X | (17) |

STI: Sexually Transmitted Infection, TB: Tuberculosis.

#### Table 5 Implementation Strategies for Identifying and Supporting Survivors of HT

| Reference | Type of strategy | Category and subcategory | Strategy description | Target audience/ Setting | Strategy outcomes |
|---|---|---|---|---|---|
| Farrel, E. 2024 (67)<br><br>U.S. | Educational strategy for healthcare personnel | **Training and support for healthcare workers**<br>Formal training: the strategy includes structured education on trafficking detection, trauma-informed care, and effective teaching methodologies. | The objective of the strategy is to develop an interprofessional community of educators trained in HT detection and response through a structured educational program. The strategy consists of a train-the-trainer model that integrates social cognitive, constructivist, and experiential learning theories, utilizing interactive methods such as case studies, survivor testimonials, and reciprocal teaching activities. It is supported by technology-enabled engagement tools, including WhatsApp and Flipgrid, to foster collaboration and sustain long-term knowledge sharing. | Healthcare and related professionals, such as physicians, nurses, social workers, advanced practice providers, psychologists, public health professionals, legal and justice personnel, and educators. | **Knowledge and Skill Improvement**<br>Significant increases in Stop, Observe, Ask, Refer (SOAR) framework understanding (+1.64 pre/post delta, $p<0.0001$).<br>Improved ability to recognize trafficking indicators (+1.04 pre/post delta, $p<0.0001$).<br>Enhanced competency in developing trafficking response protocols (+1.27 pre/post delta, $p<0.0001$).<br><br>**Sustained Learning Impact (3-month follow-up)**<br>Continued use of SOAR framework (+4.38 cohort-level delta).<br>Increased application of adult learning principles (+4.15 delta).<br>Strengthened protocol development in home organizations (+4.00 delta).<br><br>**Community of Practice Growth**<br>Active WhatsApp group with 2,199 unique posts (2020-2023).<br>27 new training modules created since 2021.<br><br>**Long-term Institutional Impact**<br>59,755 clinicians trained by program graduates as of 2023.<br>Expansion to regional in-person courses and continued virtual training. |
| Olivieri, S. 2024 (68)<br><br>U.S. | Educational strategy for healthcare personnel | **Training and support for healthcare workers**<br>Formal training: the strategy focuses on a structured and formal educational module aimed at nurses and nurse practitioners in emergency and urgent care settings, with the goal of | The objective of the study is to evaluate the effectiveness of an online educational module on HT in increasing the knowledge and confidence of emergency and urgent care nurses and nurse practitioners in New York. The strategy consisted of a pretest/post-test design using an | Nurse practitioners / EDs and urgent care centers | The study results showed a significant improvement in participants' knowledge and self-confidence in identifying and managing HT victims in urgent and emergency care settings:<br><br>**Knowledge Improvements:**<br>Ability to assess danger in trafficking cases increased from 2.15 to 3.24 ($z = -2.08$, $P = .04$). |

| | | | | | |
|---|---|---|---|---|---|
| | | enhancing their ability to identify and manage HT cases. | asynchronous online module covering key topics on identifying, assessing, and treating trafficking victims, supported by recognized research and resources. | | Recognition that trafficking is not limited to women and girls in prostitution (z = -3.00, P < .01).<br>Awareness that trafficking affects over 20 million people annually (z = -3.34, P < .01).<br>Identification of associated symptoms, like PTSD (z = -3.50, P < .01) and chronic headaches (z = -3.39, P < .01).<br>Recognition of victims' difficulties in reporting their situation (z = -3.95, P < .01).<br><br>**Confidence Gains:**<br>Increased confidence in asking about exploitative situations (z = -2.36, P = .02).<br>Improved ability to make appropriate referrals, especially for child victims (z = -1.87, P = .05).<br>Enhanced confidence in taking action, such as notifying authorities (z = -2.09, P = .04).<br>Reduced perception of lacking sufficient training (z = -2.28, P = .02). |
| Rajaram, S. S. 2024 (69)<br><br>U.S. | Educational strategy for healthcare personnel | **Training and support for healthcare workers**<br><u>Formal training:</u> The workshop is a structured educational program aimed at improving healthcare workers' knowledge and skills in identifying and responding to HT. | The study aims to assess the impact of a 3-hour educational workshop on HT in enhancing nurses' knowledge and skills to identify and treat victims of this crime.<br>The intervention consisted of an in-person workshop that was live-streamed and recorded for later availability as a webinar. The content of the workshop included an introduction to HT, its health impact, identification and assessment strategies, and response and follow-up methods. The workshop was delivered by a forensic nurse with expertise in the field and a public health faculty member, both of whom are experts in HT. | Nurses / Academic and healthcare setting | **Knowledge Improvement:**<br>Nurses' perceived adequacy of knowledge about HT increased from a baseline score of 2.47 (SD = 0.76) to 4.27 (SD = 0.69) after the workshop, with a statistically significant change (p < 0.001).<br><br>**Ability to Define HT:**<br>Their ability to define HT and identify vulnerability factors improved from a pretest mean of 2.47 (SD = 0.89) to 4.69 (SD = 0.79) (p < 0.001).<br><br>**Health Impact Understanding:**<br>Nurses' understanding of the health impact of HT increased from 2.76 (SD = 1.03) to 4.62 (SD = 0.81) (p < 0.001).<br><br>**Identification and Response:**<br>Nurses showed significant improvement in their ability to discuss HT identification, assessment strategies, and follow-up procedures.<br>**Impact of Prior Training:** Nurses with no prior training experienced the most significant improvement, while those with prior training also benefited from the workshop. |

| | | | | | |
|---|---|---|---|---|---|
| | | | | | **Intended Practice Changes:** 100% of participants expressed their intention to apply the knowledge gained, though barriers like patient compliance and lack of resources were cited. |
| Shue-McGuffin, K. 2024 (70)<br><br>U.S. | Educational strategy for healthcare personnel | **Training and Support for Healthcare Workers**<br><u>Formal training:</u> The study explicitly involved structured training through the HOPE Training modules, designed to educate Family Nurse Practitioner (FNP) students about HT detection and management. The modules covered trafficking indicators, trauma-informed care, and culturally sensitive approaches. | The study evaluated the effectiveness of the HOPE Training program in enhancing the knowledge, attitudes, and confidence of Family Nurse Practitioner (FNP) students in identifying and managing HT victims, incorporating a mixed-method design. It was conducted in two phases: an online education intervention using HOPE Training modules with pretest and post-test evaluations, followed by interprofessional simulations and focus groups to explore student's perceptions and readiness. | Family Nurse Practitioner students / University of North Carolina at Charlotte<br><br>North Carolina, U.S. | **Knowledge Improvement:**<br>Pretest mean score: 69.5.<br>Post test mean score: 89.2.<br>Statistical significance: Knowledge scores increased significantly across all participants, with a p-value of $<0.001$.<br><br>**Confidence Gains:**<br>**Confidence Levels (Pre-intervention vs. Post-intervention)**<br>Initial confidence:<br>62.5% reported no confidence in their knowledge of HT.<br>50% felt no confidence in identifying trafficking victims.<br>56.3% felt no confidence in implementing interventions.<br>62.5% lacked confidence in offering appropriate resources.<br>Post-simulation self-confidence:<br>Mean scores ranged from 4.8 to 5.0 (out of 5.0), indicating high confidence in mastering the simulation content and applying it to clinical practice.<br><br>**Satisfaction with Simulation:**<br>All students rated satisfaction items at the maximum score of 5.0 (out of 5.0), reflecting high approval of the teaching methods, materials, and simulation design.<br>Students described the experience as "informative," "eye-opening," and "helpful." |
| Briggs, MR 2023 (71)<br><br>U.S. | Educational strategy for healthcare personnel | **Training and support for healthcare workers**<br><u>Formal training:</u> The training is specifically designed for obstetrics/gynecology (OBGYN), emergency medicine (EM), and family medicine (FM) residents to improve their knowledge and confidence in caring for trafficking victims. | The objective of this study was to develop and evaluate a sex trafficking education program designed for obstetrics/gynecology (OBGYN), emergency medicine (EM), and family medicine (FM) residents. The program aimed to increase residents' knowledge and confidence in caring for individuals involved in sex trafficking. | OBGYN, EM, and FM residents / Academic medical center | **Knowledge Improvements:**<br>There was a statistically significant increase in residents' knowledge immediately after the education session, which persisted five months later, although it slightly decreased over time. Knowledge increased from a mean of 5.92 (pre-session) to 7.71 (post-session) and remained higher at 7.25 after 5 months.<br><br>**Confidence Gains:**<br>Confidence in identifying and screening for sex trafficking victims, providing appropriate care, and |

| | | | | | |
|---|---|---|---|---|---|
| | | | The strategy involved a 45-minute evidence-based education session that included a PowerPoint presentation and case discussions. It covered the identification and treatment of sex trafficking victims, trauma-informed care, and local and national resources. The education session was delivered both in-person and virtually. Surveys (pre-, post-, and 5 months after) were used to assess changes in residents' knowledge, confidence, and perceived barriers to care. A focus group was also conducted to gather further insights from participants. | | discussing the topic with patients significantly increased after the education session, but this confidence slightly decreased five months post-session.<br><br>**Encounters with Victims:** More residents reported having encountered sex-trafficked individuals after the education session (75% post-session, 85% after 5 months).<br><br>**Barriers to Care:** Before the education, the most common barriers were a lack of training, awareness, and organizational guidelines. After the session, these barriers were less frequently reported, with "lack of awareness" and "delicate subject matter" remaining as significant obstacles.<br><br>**Satisfaction:** A majority of residents were satisfied with the training and strongly agreed that sex trafficking education should be a standard part of medical education. |
| Cavey, W. 2023 (72)<br><br>U.S. | Educational strategy for healthcare personnel | **Training and Support for Healthcare Workers**<br>Formal training: a structured educational program on identifying and treating HT victims, focusing on trauma-informed care. | A quasi-experimental design with a 20-minute educational intervention available through YouTube. The intervention included a presentation on HT related topics such as definitions, types, vulnerability factors, health impact, victim identification and assessment, as well as response and follow-up recommendations. Participants completed a self-efficacy measurement survey before and after the intervention. | Healthcare professionals / Online platform | The educational intervention significantly increased healthcare professionals' confidence in identifying and treating HT victims.<br>**Overall improvement in self-efficacy:**<br>Before the intervention: $M = 2.6427$ ($SD = 0.4888$).<br>After the intervention: $M = 1.9758$ ($SD = 0.3467$), with a significant difference ($t(30) = 7.1888$, $p < .001$).<br>**Survey results:**<br>Preparation to ask about HT: Before $M = 2.79$, after $M = 1.90$ ($p < .001$).<br>Responses to victims saying "yes": Before $M = 2.83$, after $M = 1.86$ ($p < .001$).<br>Support during the interview: Before $M = 2.38$, after $M = 1.86$ ($p < .01$).<br>Access to resources for victims: Before $M = 2.41$, after $M = 1.72$ ($p < .01$). |
| Kelly, K 2023 (73)<br><br>U.S. | Educational strategy for healthcare personnel | **Training and support for healthcare workers**<br>Formal training and interprofessional education: The program involved training medical residents and other healthcare professionals in an | A HT training program for medical residents, specifically through the Medical Safe Haven (MSH) model, which provides trauma-informed care (TIC) for victims of trafficking. This program was implemented at | Medical residents / Family medicine residency clinic | A significant increase in resident confidence in all aspects evaluated after completing the training. Residents reported an increase in their ability to identify trafficking victims, understand the effects of trauma, and provide trauma-informed care. |

| | | | | | |
|---|---|---|---|---|---|
| | | interdisciplinary manner , providing structured education on HT and trauma-informed care (TIC).<br><u>Educational materials:</u> Educational resources were distributed, and simulations and case studies were used to reinforce the concepts learned. | Dignity Health Family Medicine residency sites in California.<br><br>The educational strategy consisted of three training sessions for the medical residents:<br>Human Trafficking Training (HTMSH): Focuses on recognizing and managing trafficking victims.<br>TIC Training: Training on the effects of trauma on health and how to provide patient-centered medical care.<br>Medical-Patient Interaction Training (PPE): Training on how to interact safely and effectively with victims. | | **Confidence Gains:** Increased confidence in identifying and treating trafficking victims, with significantly higher scores after the training.<br><br>**Awareness:** On physical indicators of trafficking victims and the importance of trauma-informed care.<br>Resident testimonies indicating that the MSH program helped them feel more comfortable discussing trafficking and better prepared to interact with those experiencing HT.<br><br>Most residents indicated that they planned to incorporate TIC models into their future practice, and some wanted to replicate the MSH model in their future workplaces. |
| Marcinkowski, B. 2022 (74)<br><br>U.S.<br><br>**Scoping review** | Educational strategy for healthcare personnel | **Training and support for healthcare workers**<br><u>Formal training:</u> It addresses how educational strategies reduce knowledge gaps, increase confidence, and provide practical tools such as simulations and assessment algorithms integrated into workflows. | This scoping review systematically evaluates existing literature on the screening, identification, and intervention for adult sex trafficking victims in EDs, aiming to identify knowledge gaps and improve understanding of current practices. The review highlights educational strategies designed to improve healthcare professionals' ability to identify and support victims of sex trafficking in EDs. These strategies include brief workshops, interactive role-play scenarios, online training modules, and mandatory education sessions for ED staff. | Healthcare Professionals/ EDs<br><br>U.S. (e.g., Washington, D.C., Baltimore, South Carolina, Pennsylvania) and one study in Alberta, Canada. | **Knowledge**<br>Online and in-person sessions improved knowledge of trafficking indicators. Knowledge retention was observed three months post-training.<br><br>**Identification**<br>Training and screening tools identified 38 victims in five months in one study.<br><br>**Role-Specific Training**<br>Tailored programs enhanced EMS, nurses, and residents' response capabilities.<br><br>**Simulation Learning**<br>Role-play and case-based exercises improved practical skills.<br><br>**Clinical Integration**<br>Training sessions encouraged using standardized tools and communication strategies. |
| Missouri Hospital Association (75)<br><br>U.S. | Educational strategy for healthcare personnel | **Training and support for healthcare workers**<br><u>Formal Training:</u> The 14 educational modules offer structured training on trafficking | This course provides training to hospital staff to identify and care for HT victims, and provides tools to develop appropriate policies and procedures in Missouri hospitals. | Healthcare professionals/ EDs, urgent care centers, and other frontline healthcare settings. | There are no reported results of its application, nor statistics on the number of people who have accessed it, nor evaluations of its effectiveness. |

| | | | | |
|---|---|---|---|---|
| | | indicators, trauma-informed care, and screening tools. | The course consists of 14 online education modules, available *on-demand*, with a practical and multidisciplinary approach. It uses evidence-based detection tools, survivor testimonies, and training in trauma-informed care. Additionally, it covers regulatory compliance and collaboration with support agencies and law enforcement. Tools and strategies included:<br>Human Trafficking Toolkit – Developed by the Hospital Human Trafficking Taskforce to assist hospitals in creating policies and procedures.<br>Online Education Modules – 14 modules covering identification, screening, trauma-informed care, and reporting.<br>Screening Protocols – Evidence-based tools for identifying trafficking victims among minors and adults.<br>Trauma-Informed Care Framework – Strategies to minimize re-traumatization and improve victim engagement.<br>Law Enforcement Reporting Guidelines – Training on when and how to report trafficking cases.<br>Type of Trafficking Addressed:<br>Commercial Sexual Exploitation of Children (CSEC)<br>Sex Trafficking (adults and minors)<br>Labor Trafficking (various industries, including agriculture, domestic work, and service sectors) | Missouri, U.S. |

| | | | | | |
|---|---|---|---|---|---|
| Murphy, M. C. 2022 (76)<br><br>U. S. | Educational strategy for healthcare personnel | **Training and support for healthcare workers**<br>Formal training: The intervention involved structured education, both online and in-person, to enhance nurses' competencies in identifying HT victims using detection tools and a trauma-informed approach.<br><br>**Information and Communication Technology (ICT)**<br>Health Information Systems: The intervention utilized an EMR system that was already in place at the hospital. The victim screening tool was integrated into the pre-existing safety assessment system of the EMR, allowing nurses to systematically assess patients for potential trafficking victims during their visits in the ED. This integration represents the use of health information systems to manage patient information and support the identification of trafficking victims. | The project implemented two key interventions: HT-specific education and the use of a safety screening form to help identify potential victims. The education was delivered through an asynchronous online learning module via the hospital's existing Learning Management System (LMS). The module included a three-minute introductory video, "Faces of Human Trafficking," created by the U.S. Department of Justice's Office for Victims of Crime. After the video, participants viewed a narrated PowerPoint presentation covering topics such as defining HT, debunking common myths, identifying victim and trafficker characteristics, understanding why victims seek medical care, using the new safety screening tool, and outlining procedures for handling suspected trafficking cases | Emergency room nurses / General ED | The results showed a significant improvement in nurses' self-efficacy regarding the identification of HT victims. Pre- and post-intervention survey data, analyzed using a paired t-test, revealed an increase in the mean self-efficacy scores, with a significant difference between pre- ($M = 2.26$, $SD = 0.216$) and post- ($M = 3.417$, $SD = 0.133$) intervention scores ($p = 0.0002$).<br><br>**Identification**<br>The improvement in self-efficacy was significant for each of the six items on the scale used to assess nurses' confidence in identifying and managing trafficking victims. This included greater confidence in the ability to identify victims, differentiate between child abuse and sex trafficking, and appropriately use the screening tool. |
| Talbott, et al., 2022 (77)<br><br>**Scoping review**<br><br>U.S. | Educational strategy for healthcare personnel | **Training and support for healthcare workers**<br>Formal training: This scoping review focuses on educational resources and training aimed at healthcare professionals on labor and organ trafficking. | This scoping review examines the availability, content, and gaps in U.S.-based educational resources for healthcare professionals related to labor trafficking and organ trafficking. The review aimed to assess whether existing materials are adequate, accessible, and applicable for training clinicians and health trainees to recognize and respond to these forms of trafficking<br>Thematic Areas Explored:<br>1. Definition of Labor Trafficking. | U.S.-based healthcare professionals across disciplines (e.g., physicians, nurses, emergency staff), including some resources for students and residents. | Reviewed 37 resources, including journal articles, manuals, toolkits, presentations, and online modules.<br>**Key Findings:**<br>86.5% defined labor trafficking.<br>64.9% described indicators or warning signs.<br>48.6% addressed prevention strategies.<br>43.2% included child labor.<br>Only 21.6% addressed organ trafficking.<br>Few resources discussed race/ethnicity (8.1%) or included trainee-focused content (8.1%).<br>24.3% offered Continuing Medical Education (CME) credit.<br><br>**Gaps Identified:** |

| | | | | | |
|---|---|---|---|---|---|
| | | | 2. Indicators and Warning Signs.<br>3. Prevention Strategies.<br>4. Child Labor Trafficking: Inclusion of information specific to minors affected by labor trafficking.<br>5. Organ Trafficking (THBOR): Content addressing the trafficking of human beings for the purpose of organ removal.<br>6. Racial and Ethnic Considerations.<br>7. Trainee-Focused Content.<br>8. CME Accreditation: Whether the resources offer Continuing Medical Education (CME) credits. | | Lack of comprehensive resources covering both labor and organ trafficking.<br>Scarcity of content on organ trafficking indicators and legal frameworks.<br>Minimal incorporation of race, ethnicity, and structural determinants of vulnerability.<br>Limited availability of concise, user-friendly tools tailored to clinicians' time constraints. |
| International Organization for Migration<br>2021 (78)<br><br>Switzerland | Educational strategy for healthcare personnel | **Training and support for healthcare workers**<br>Formal training: This is a virtual course for healthcare providers available in English and Spanish, which can be accessed at any time. It is provided by the International Organization for Migration (IOM) through a web page that can be freely accessed from different countries. | This course provides healthcare providers and other professionals with practical, non-clinical guidance on understanding HT. It equips them to recognize and respond to trafficking situations while ensuring a safe and trauma-informed approach to healthcare.<br>This is a virtual course available in English and Spanish, accessible year-round at any time.<br>The course consists of the following modules:<br>Introduction<br>Module 1: A brief introduction to HT<br>Module 2: Health consequences for victims of HT<br>Module 3: Trauma-informed care<br>Module 4: The role of the healthcare provider | Healthcare providers from different countries | There are no reported results of its application, nor statistics on the number of people who have accessed it, nor evaluations of its effectiveness. |

| | | | | | |
|---|---|---|---|---|---|
| Lee, H. 2021 (79)<br><br>U.S. | Educational strategy for healthcare personnel<br><br>**The Learn to Identify and Fight Trafficking (LIFT) training** | **Training and support for healthcare workers**<br>Formal training: The strategy of the study focused on a structured training program for healthcare professionals on how to identify and manage trafficking victims, using an educational approach that includes both didactic content and active discussions. The training also covered trauma-informed and culturally sensitive approaches, aligning with this subcategory. | The objective was to measure the impact of this CME-accredited training on healthcare professionals' knowledge and attitudes about HT before and after attending the course.<br>The Learn to Identify and Fight Trafficking (LIFT) training program curriculum aimed to improve healthcare professionals' knowledge on recognizing HT victims and providing appropriate care. The training covered the scope and prevalence of trafficking, warning signs, trauma-informed care, and available resources for victims.<br>The study involved 17 LIFT training sessions held across various U.S. cities. Healthcare providers who attended the training completed a pre-test, post-test (1-week after), and follow-up test (6 months later). The tests assessed participants' knowledge and attitudes on HT. | Healthcare professionals/ General healthcare settings | **Knowledge**<br>The study found that the LIFT training significantly improved healthcare professionals' knowledge about HT, increasing from 54.7% to 84.5% one week after the training. Attitudes also improved from 49.4% to 71.0% immediately after the course. At 6 months, attitudes remained stable, but knowledge decreased to 50%. This suggests a short-term improvement in both areas, with a decline in knowledge over time. |
| The CMDA Commission on Human Trafficking, 2020 (80)<br><br>U.S. | Educational strategy for healthcare personnel | **Training and support for healthcare workers**<br>Formal training: An online educational course designed to equip healthcare professionals with foundational knowledge about human trafficking, including clinical indicators, appropriate responses, and trauma-informed care principles. The module is self-paced and available through the Christian Medical & Dental Associations (CMDA) learning platform. It represents a scalable and accessible tool to raise awareness and improve provider readiness in clinical settings. | This online, self-paced course is designed to train healthcare professionals in the identification, care, and comprehensive response to victims of human trafficking, both domestic and international.<br>It consists of 12 modules covering clinical, legal, ethical, and spiritual aspects, with specific content tailored to low-resource settings and trauma-informed care.<br>Key topics include:<br>- Identifying signs and symptoms of trafficking in patients<br>- Medical evaluation of victims of sexual and labor trafficking<br>- Coordination with law enforcement, child protection | Healthcare providers (physicians, nurses, dentists, and physician assistants) | There are no reported results of its application, nor statistics on the number of people who have accessed it, nor evaluations of its effectiveness. |

| | | | | | |
|---|---|---|---|---|---|
| | | | services, shelters, and forensic teams<br>- Use of trauma-informed approaches and multidisciplinary care models<br>- Mental health care and spiritual support for survivors<br>- Protocol development, referral systems, and follow-up care<br>- Public health perspectives and biblical foundations for anti-trafficking response | | |
| Fraley, H.E. 2020 (81)<br><br>U.S.<br><br>**Systematic review** | Educational strategy for healthcare personnel | **Training and support for healthcare workers**<br>Formal training: the study focuses on structured educational interventions designed to increase awareness, knowledge, and skills of healthcare providers (HCPs) in identifying and addressing HT. | The literature review aimed to identify existing HT educational interventions targeting HCPs and evaluate their effectiveness. It included peer-reviewed, English-language original research studies published between January 1, 2000, and September 1, 2018, excluding earlier studies due to the increased global awareness following the adoption of the 2000 UN Protocol to Prevent, Suppress and Punish Trafficking in Persons. The U.S. federal definition of HCPs was used, encompassing physicians, dentists, chiropractors, nurses, clinical psychologists, and social workers. Search terms included HT, healthcare providers, commercial sexual exploitation, educational intervention, awareness, attitudes, knowledge, children, youth, victims, and prevention, providing a systematic approach to assessing the reach and effectiveness of educational efforts. | Healthcare providers / Multiple settings | **Increased Knowledge and Awareness:**<br>All studies reported significant improvements in knowledge, awareness, and confidence in identifying trafficking victims.<br>Attitudes toward victims shifted from negative perceptions to recognizing them as victims of trafficking.<br><br>**Behavioral Impact:**<br>One study found increased awareness did not translate into more referrals of trafficking victims.<br><br>**Duration of Interventions:**<br>Shorter sessions (25 minutes) were as effective as longer ones (60 minutes) in improving knowledge and awareness.<br><br>**Instrument Evaluation:**<br>Most studies reported content validity for their instruments.<br>Only one study reported high internal consistency ($\alpha > 0.9$). |
| Scannell, M. 2020 (82) | Educational strategy for | **Training and support for healthcare workers** | The strategy is the SASH course, a sexual assault simulation designed to train emergency staff to identify | Healthcare professionals, specifically the | **Knowledge**<br>The results of the SASH course showed a significant increase in participants' knowledge and perceived |

| | | | | | |
|---|---|---|---|---|---|
| U.S. | healthcare personnel | Formal training: The SASH course is a structured training program that teaches ED healthcare workers how to identify trafficking indicators and use a trauma-sensitive approach.<br><br>. | victims of sex trafficking. It includes theoretical lessons, evidence collection practice, simulations with actors, and a debriefing session which promotes a trauma-informed approach. | nursing staff / ED of an urban hospital | competence in identifying HT victims. Specifically, pre- and post-course tests were administered, with the following results:<br>Average pre-test score: 64.27<br>Average post-test score: 81.60<br>Statistical significance level: P = 0.00023<br>These data demonstrate a significant improvement in participants' knowledge about HT detection. Additionally, a few months after the course, one of the newly trained nurses identified a trafficking victim in the ED, applying what was learned during the training. |
| Donahue, S. 2019 (83)<br><br>U.S. | Educational strategy for healthcare personnel<br><br>Implementation of a HT survivor response tool | **Training and support for healthcare workers**<br>Formal training: The online educational module (HTEmergency.com) is a structured training module that equips healthcare workers with the necessary tools to identify and treat trafficking victims in EDs.<br><br>**Community and interagency collaboration**<br>Referral systems: The project references the National Human Trafficking Hotline as a resource for referral, implying a referral system for victims. | The objective of this project was to educate ED staff on the identification and treatment of HT victims and to develop and implement a screening tool with care guidelines for those identified as potential victims of trafficking.<br><br>The strategy involved an evidence-based online training module (HTEmergency.com) that included a PowerPoint presentation, case studies, and identification and treatment guidelines. The module covered the definition of HT, red flags, at-risk populations, screening questions, and health implications.<br>A step-by-step care guideline was provided, including an assessment tool and a reference to the National Human Trafficking Hotline. | Healthcare personnel / EDs | The training effectively increased ED staff's confidence and knowledge regarding HT, which is essential for improving the identification and care of trafficking victims in the ED setting.<br><br>**Training Impact:**<br>89% of participants had not received prior training on HT.<br>The training module significantly improved confidence in identifying (from 4/10 to 7/10) and treating (from 4/10 to 8/10) HT victims.<br>93% of participants reported a comprehensive understanding of HT after the education (up from less than half before).<br>96% of participants found the module useful in their work setting.<br><br>**Increased Screening and Confidence:**<br>After the training, staff were more likely to screen patients for HT and felt more confident in their ability to identify and treat trafficking victims.<br>The project also included a practical, readily accessible screening tool (a laminated page) to assist ED staff in identifying potential victims. |
| Berishaj, K. 2018 (84)<br><br>U.S. | Educational strategy for healthcare personnel | **Training and support for healthcare workers**<br>Formal training and interprofessional education: The conference addressed these aspects by training nurses on how to identify trafficking victims and | The intervention consisted of a 4-hour educational conference titled "Human Trafficking 101: A Practical Conference on Understanding the Issues and Responding to the Epidemic." The conference was delivered by HT experts with backgrounds in nursing, | Registered nurses (RNs) / Public university | **Knowledge:**<br>The results of the study showed that the educational intervention significantly improved nurses' knowledge and beliefs about HT. There was an improvement in knowledge about the difference between labor and sex trafficking, trafficking-related laws, and how to identify and assist victims. |

| | | | | | |
|---|---|---|---|---|---|
| | | use available resources for their support. | law, and criminal justice. The objectives were to teach nurses to identify signs of trafficking, understand the differences between labor and sex trafficking, recognize relevant state and federal laws, and learn about available resources to assist victims. | | **Confidence Gains:**<br>Confidence in the ability to identify victims and manage the available resources to support them also increased.<br><br>**Attitudes:**<br>Attitudes towards trafficking also improved, especially regarding the understanding that trafficking can happen in any community and the belief that nurses can make a difference in the fight against trafficking. However, no significant change was observed in the intention to be actively involved in the fight against trafficking. |
| Liverseed, G 2018 (58)<br><br>U.S. | Educational strategy for healthcare personnel<br><br>Implementation of a HT survivor response protocol. | **Interventions directed at health workers**<br>**Training and support for healthcare workers**<br><u>Formal training:</u> The implementation of a 30-minute educational webinar aligns with this subcategory, as it aimed at improving the clinicians' knowledge and practices.<br><br><u>Clinical Practice Guidelines:</u> The introduction of a formal HT response protocol for clinicians also aligns with this subcategory, as it provides structured guidance on managing trafficking cases.<br><u>Continuous Quality Improvement</u>: The iterative process of assessing preparedness and modifying practices aligns with continuous quality improvement approaches, aiming to improve clinician practices over time.<br><br>**Interventions directed at health organizations**<br><u>Organizational Culture:</u> Although not explicitly stated, the intervention sought to change the | A quality improvement project implemented in a reproductive healthcare organization to evaluate the impact of a HT response protocol on clinicians' preparedness to identify, assess, and respond to trafficking victims. Below are the requested details:<br>The main strategy was the implementation of a HT response protocol in a reproductive healthcare setting. This protocol was introduced to clinicians through a 30-minute educational webinar. The protocol included indicators of victimization, recommended screening questions, and steps to follow if a clinician suspected a patient was a trafficking victim. The goal was to increase clinician awareness and preparedness to identify and respond to HT in reproductive health services. | Clinicians / Reproductive Health Settings / General healthcare settings | **Increased Clinician Preparedness:**<br>A statistically significant increase was observed in clinicians' self-reported preparedness to identify, assess, and respond to HT victims across all evaluated areas.<br>Specific areas that showed improvements included:<br>Identifying indicators of HT.<br>Helping victims assess their safety and readiness to change.<br>Asking appropriate trafficking-related questions during consultations.<br>Responding adequately to trafficking disclosures.<br>Using a trauma-informed approach when interacting with victims.<br>Creating safety plans and referring victims to appropriate services.<br>Fulfilling state reporting requirements for HT.<br>These improvements were statistically significant with p-values $< 0.05$.<br><br>**Increased Frequency of HT Screening**:<br>Clinicians showed a significant increase in the frequency of asking screening questions about HT when warning signs were present. Areas that showed improvements included:<br>High number of sexual partners.<br>Inconsistent or scripted history.<br>Multiple pregnancies or abortions.<br>Accompanied by a controlling person.<br>Multiple sexually transmitted infections.<br>Hyper-vigilance or subordinated demeanor. |

| | | | | | |
|---|---|---|---|---|---|
| | | organizational approach to HT, suggesting a shift in organizational culture to incorporate better identification and response practices.<br><u>Processes of Local Consensus:</u><br>The development and implementation of the HT response protocol might involve consensus within the organization regarding how to handle cases of trafficking. | | | Discrepancy between history and clinical presentation.<br>Several somatic symptoms arising from stress.<br>These increases were also statistically significant ($p < 0.05$).<br><br>**Protocol Usage:**<br>94% of clinicians reported that the HT response protocol was available in their clinics, though only 17.6% considered it to be widely used. This suggests that while the protocol was in place, its use in clinical practice was not fully widespread.<br>4. Referral Resources for Trafficking Victims:<br>82.4% of clinicians reported that they felt they had adequate referral resources for trafficking victims at their clinics. This indicates an improvement in available resources to support victims, although a small percentage of clinicians still felt the resources were insufficient. |
| Egyud, A 2017 (85)<br><br>U.S. | Educational strategy for healthcare personnel<br><br>Evaluation of the implementation of a screening tool for HT | **Training and support for healthcare workers**<br><u>Formal training</u>: The project involved multidisciplinary training (nurses, physicians, security personnel, etc.) on using screening tools and rescue protocols.<br><br><br>**Communication Technology (ICT):**<br><u>Health information systems</u>: The electronic screening tool integrated into the EMR system allowed for systematic patient evaluations and recording of red flag signs. | The project implemented a treatment algorithm and screening tool for identifying HT victims in the ED. The strategy consisted of a multidisciplinary approach, involving nurses, physicians, security personnel, social services, and other hospital staff. Key components included:<br>Education: Mandatory training was provided for ED staff (nurses, physicians, registration staff, security, etc.), which included training on trafficking victim screening tools, medical red flags (such as urinary tract infections, pelvic pain, suicide attempts, etc.), and available resources for victim rescue.<br>Screening Tool and Notification System: An electronic screening tool was integrated into the hospital's EMR system for victim detection through medical red flags and a silent notification system (using a | Healthcare staff / ED | The results showed that education and the implementation of the screening tool significantly improved the staff's ability to identify victims of trafficking. Over 5 months of follow-up, the results were:<br><br>**Identification**<br>Identification of Victims: A total of 38 potential trafficking victims were identified, with 20 identified through medical red flags (53%) and 18 through the silent notification system (47%).<br><br>**Acceptability**<br>Interventions Accepted: 5 patients accepted intervention. Of these, 1 was identified as a true trafficking victim after a suicide attempt, and the others were identified through other forms of abuse (domestic violence, sexual abuse, etc.).<br><br>**Staff Engagement**<br>97% of participants in the training expressed willingness to change their practices, and 74% perceived that the training improved their competence in identifying trafficking victims. |

| | | | | | |
|---|---|---|---|---|---|
| | | | blue dot on urine samples from patients).<br><br>Rescue Planning: In cases where a victim was identified, a huddle was held with the healthcare team to coordinate rescue actions, including notifying the appropriate law enforcement and government agencies. | | |
| Grace, A. 2014 (86)<br><br>U.S. | Educational strategy for healthcare personnel | **Training and support for healthcare workers**<br>Formal training: The strategy is based on a structured formal training through a standardized educational presentation that provides healthcare providers with the necessary knowledge about HT, clinical signs, and referral options. | The objective of this study was to assess whether an educational presentation increased ED providers' recognition of HT victims and enhanced their knowledge of resources available to manage cases of HT. The strategy involved a randomized controlled trial in which 20 of the largest EDs in the San Francisco Bay Area were randomly assigned to either an intervention or a delayed intervention comparison group. The intervention consisted of a standardized educational presentation about HT, including background information, clinical signs, and referral options. The presentations were delivered by law enforcement and medical professionals and were available in both short (25 minutes) and long (60 minutes) versions. | Healthcare professionals/ EDs | A brief educational intervention effectively increased ED providers' knowledge and self-reported ability to recognize and manage cases of HT.<br><br>**Knowledge and Recognition Improvements:**<br>The intervention group showed a significant increase in self-rated knowledge about HT, with an average increase of 1.42 points compared to a decrease of -0.15 in the comparison group ($P < 0.001$).<br>The proportion of participants who knew who to call for potential HT victims increased from 7.2% to 59% in the intervention group, while it remained unchanged at 15% in the comparison group ($P < 0.01$).<br>The proportion of participants who suspected that a patient was a victim of HT increased from 17% to 38% in the intervention group, while it remained unchanged at 10% in the comparison group ($P < 0.01$).<br><br>**Attitudes T owards HT Knowledge:**<br>There was no significant change in how participants rated the importance of knowledge about HT for their profession, but knowledge of HT and recognition of its signs improved significantly in the intervention group compared to the delayed intervention group. |
| Ahn, R. 2013 (86)<br><br>U.S<br><br>**Systematic review** | Educational strategy for healthcare personnel | **Training and Support for Healthcare Workers:**<br>Formal Training: The study emphasizes the need for structured training programs for healthcare professionals, focusing on the identification and management of trafficking victims**.** The | The study aimed to review educational resources on HT for healthcare professionals to enhance awareness, victim identification, and treatment within healthcare settings while providing recommendations for curriculum development. This was achieved through a literature | Healthcare professionals / General healthcare settings<br><br>Canada and the UK | **Resources identified:**<br>A total of 27 educational resources on HT and healthcare were reviewed, primarily published after 2003. These included practical tools such as guides, manuals, protocols, online courses, academic articles, informational sheets, and issue briefs.<br><br>**Topics covered:** |

| | | | | | |
|---|---|---|---|---|---|
| | | importance of tailoring educational strategies to specific healthcare settings, such as EDs or specialized clinics, is highlighted. | review of peer-reviewed and gray literature from databases like PubMed and EMBASE, using a structured keyword search to identify English-language resources designed specifically for healthcare audiences. | | Definitions and scope of HT.<br>Physical and psychological health consequences for victims.<br>Indicators for victim identification.<br>Medical treatment based on cultural sensitivity and trauma-informed care.<br>Referral to complementary services, such as housing, legal assistance, and additional medical care.<br>Legal aspects, including proper documentation and patient confidentiality.<br>Safety for victims and healthcare professionals.<br>Prevention, although addressed only minimally.<br>Target audience: Most resources were designed for a general audience of healthcare professionals, although some targeted specific groups such as ED staff, psychotherapists, and nurses.<br><br>**Limited evaluation:** None of the reviewed resources were rigorously evaluated in terms of their impact on healthcare professionals' behaviors or the identification of trafficking victims.<br><br>**Lack of prevention guidance**: Very few resources provided concrete recommendations for healthcare professionals to actively participate in preventing HT. |
| Chisolm-Straker, M. 2012 (87)<br><br>U.S. | Educational strategy for healthcare personnel | **Training and support for healthcare workers**<br>Formal training: This is a structured and didactic training program focused on the identification and treatment of trafficking victims in the emergency setting.<br><br>. | To assess the knowledge and confidence levels of emergency care providers in identifying and treating HT victims before and after receiving a specific educational workshop on the topic.<br>The study included a 20-minute training intervention with a narrative approach, using a real case of an unidentified trafficking victim who presented in an ED. This didactic session explained the signs emergency staff should recognize, how to intervene safely, and recommended clinical treatment guidelines for trafficking victims.<br>The training was conducted in four | Healthcare professionals / EDs | The vast majority, 97.8%, indicated they had not received formal training on the clinical presentation of trafficking victims, and 95% had not received training on the appropriate treatment for these victims.<br><br>**Changes After Training:**<br>Following the 20-minute educational intervention, 90.3% of participants reported feeling confident or very confident in their ability to define HT, a significant improvement from the initial 19.2%.<br>Confidence in the ability to identify a victim increased to 53.8% after the training, compared to 4.8% before the intervention.<br>Similarly, confidence in treating a trafficking victim rose to 56.7% after training, compared to 7.7% prior. |

| | | | | | |
|---|---|---|---|---|---|
| | | | institutions, and participants completed questionnaires before and after the intervention to assess changes in their knowledge and confidence | | The session was well-received: 93.3% of participants found the training useful, 76% felt it was well-organized, and 71.2% rated it as thorough.<br><br>**Participant Feedback:**<br>Feedback on the session was uniformly positive, reflecting an increased understanding of the topic's importance.<br>Several participants highlighted their previous lack of awareness about HT and its relevance in the emergency care context.<br>Requests were also received to repeat the session for emergency nursing staff and social workers, indicating widespread interest in continued training on this topic. |
| **Analysis of Practices in Healthcare Settings** | | | | | |
| Balasa, R. 2024 (88)<br><br>Canada | Analysis of practices for identifying child victims of sex trafficking in pediatric healthcare settings | **Contextual adaptation and customization**<br><u>Setting-specific adaptation:</u><br>The study focuses on adapting practices to the specific context of PEDs, accounting for the unique challenges of high workload, lack of privacy, and the pediatric patient demographic.<br><u>Barrier and facilitator assessment:</u><br>Barriers like lack of tools, training, and time are discussed, along with the need for tailored interventions to address these obstacles.<br>**Organizational strategies**<br><u>Organizational culture:</u><br>The study underscores the need for organizational support to foster a culture conducive to identifying trafficking victims, including leadership training and supportive policies. | The study aimed to examine healthcare providers' practices in identifying child sex trafficking in Ontario PEDs, addressing challenges such as the lack of formal screening tools and exploring effective methods within a trauma- and violence-informed framework. Using a qualitative descriptive design as part of a larger mixed-methods framework, the study conducted semi-structured interviews with 12 healthcare providers from Ontario pediatric EDs between March and September 2023. Thematic analysis, guided by intersectionality theory, was employed to understand the interrelated identities and power dynamics influencing trafficking identification practices. | Healthcare providers / PEDs<br><br>Ottawa, Toronto, Hamilton, and London.<br>Canada. | **Profile of Patients Identified as CEST:**<br>Predominantly white female adolescents between 12 and 17 years old, although the underrepresentation of marginalized groups such as Indigenous and Black girls was recognized.<br>Vulnerability factors include low socioeconomic status, adverse childhood experiences, and involvement in the child welfare system.<br><br>**Common Reasons for ED Visits:**<br>Physical conditions such as sexually transmitted infections, pregnancies, traumatic injuries, and sequelae of sexual assaults.<br>Mental health issues such as suicidal ideation, self-harm, and trauma-related disorders.<br><br>**Identification Practices:**<br>Registered nurses were the primary identifiers due to their close and continuous interaction with patients.<br>The most effective strategies included trauma- and violence-informed care approaches, private interviews, and trust-building with patients.<br><br>**Barriers to Identification:**<br>Lack of standardized screening tools.<br>High workload in EDs, lack of privacy in triage, and scarcity of specific training. |

| | | | | | |
|---|---|---|---|---|---|
| | | | | | Underrepresentation of marginalized populations in urban pediatric services. |
| Eickhoff, L. 2023 (89)<br><br>U.S.<br><br>Systematic review | To evaluate practices for identifying adult victims of HT | **Outcome evaluation and continuous improvement**<br>Usage data monitoring: Tracking data on the frequency and effectiveness of detection tools to evaluate and improve identification practices.<br>Patient impact assessment: Measuring patient outcomes, such as successful referrals, to assess the practical impact of the detection tools**.** | The study aimed to identify best practices for detecting sex trafficking among adults in U.S. EDs, comparing multifaceted screening systems to standardized questions. Using an integrative review, 11 articles published between 2016 and 2021 were selected from databases like PubMed and CINAHL. Inclusion focused on peer-reviewed studies in adult ED settings, excluding pediatric and non-ED research. Articles were appraised with the Johns Hopkins model, identifying four themes: education, protocols, legal considerations, and multidisciplinary teamwork. Ethical approval was not required. | Healthcare providers / Adult ED<br><br>Baltimore, Maryland, U.S. | **Screening Tools**: Multifaceted systems, including silent flag methods like the "Blue Dot Campaign" improve detection over standardized question lists.<br><br>**Provider Education**: Training on trafficking indicators and trauma-informed care enhances detection and patient trust.<br><br>**Protocols:** Tailored protocols and tools like RAFT perform better than no system or basic screening.<br><br>**Legal and Ethical Considerations**: Objective documentation is crucial; reporting varies by state, requiring adult patient consent.<br><br>**Multidisciplinary Approach**: Collaboration among healthcare workers, social workers, and legal experts improves outcomes.<br><br>**Barriers:** False negatives and lack of training hinder effective detection. |
| Hulick, J. 2022 (90)<br><br>U.S. | Analysis of current screening practices for sexual exploitation in EDs | **Contextual adaptation and customization**<br>Setting-specific adaptation: The study examines the implementation of screening tools in EDs, acknowledging the fast-paced environment and the need to adapt tools and approaches to fit within existing ED workflows.<br>Barrier and facilitator assessment: The participants identify several barriers to effective screening, such as time constraints, lack of training, and the absence of clear protocols.<br><br>**Organizational strategies**<br>Organizational culture: The study examines the organizational | The study aims to examine current screening practices for sexual exploitation in EDs in hospitals in Western Washington and assess the readiness of nurses and facilities to implement a standardized identification system.<br><br>The methodology used was qualitative and exploratory, divided into two stages: first, a literature review to identify tools and warning signs in detecting exploited individuals in healthcare settings; and second, in-depth interviews with nursing leaders and frontline nurses in three urban hospitals. These interviews sought to understand their current practices, the | Nurses / ED | **Role Clarity:** Nurses reported confusion about their roles in identifying and managing trafficked individuals, with unclear responsibilities across the healthcare team.<br><br>**Time Constraints:** The fast-paced ED environment makes it difficult for nurses to establish rapport with patients, limiting their ability to effectively screen for trafficking.<br><br>**Clinical Judgment**: Nurses rely on informal screening based on clinical judgment and patient behavior, as there are no specific protocols for trafficking detection.<br><br>**Need for Decision Pathways:** Nurses emphasized the need for standardized algorithms or decision pathways to improve identification and care for trafficked patients. |

| | | | | | |
|---|---|---|---|---|---|
| | | culture in EDs regarding the detection of trafficking victims, including the absence of formal protocols and screening tools, as well as the desire to improve care quality. | acceptance of existing screening questions, and their perception of the usefulness of a standardized tool | | **Missed Opportunities**: The lack of training and formal processes leads to missed opportunities in identifying trafficking victims.<br><br>**Support for Screening Tools**: Nurses agreed on the necessity of a standardized screening tool to better identify and manage cases of trafficking in the ED. |
| Ram, S. 2022 (57)<br><br>U. S.<br><br>**Systematic review** | Analysis of clinicians' knowledge, attitudes, and behaviors regarding the identification and assistance of trafficking-in-persons in healthcare settings | **Organizational strategies**<br>Organizational culture: This analysis contributes to building a supportive organizational culture by identifying gaps in knowledge and attitudes, promoting leadership training, and fostering an environment conducive to effective detection and support for HT.<br>Quality improvement: The systematic review and assessment of clinician practices align with the goal of continuous improvement. The evaluation of knowledge and behaviors supports iterative cycles to monitor, refine, and enhance the implementation of trafficking detection tools. | This study aimed to examine primary care clinicians' knowledge, attitudes, and behaviors in identifying and assisting HT in healthcare settings, considering cultural influences and barriers. Using a systematic review guided by PRISMA, the authors searched PubMed, Medline Plus, and CINAHL (2016–2021). Inclusion criteria focused on quantitative, English-language, peer-reviewed articles addressing clinicians' roles with HT. Of 130 initial articles, 10 met the criteria, with data extraction and synthesis independently conducted by the authors. | Health care professionals / Primary care and clinical settings<br><br>Miami, Florida, U.S. | **Clinician Knowledge:** Many clinicians lacked sufficient knowledge to identify effectively.<br>Training was identified as a key factor in improving recognition of HT indicators and providing appropriate resources.<br><br>**Clinician Attitudes:** Limited confidence and preparedness were common among clinicians, contributing to reluctance in pursuing identification or intervention. Training protocols improved confidence and preparedness significantly post-implementation.<br><br>**Clinician Behaviors:** Behavioral changes, such as increased use of screening tools and communication strategies, were noted after training programs. Educational interventions led to a higher likelihood of clinicians identifying HT and referring them for appropriate services.<br><br>**Barriers to Identification:** Lack of training and resources in healthcare settings impeded effective detection and assistance. Cultural and behavioral factors, including stigma and fear of disclosure, further limited HT identification.<br><br>**Health Effects of HT**: HT presented with physical injuries (e.g., broken bones, chronic pain, sexually transmitted infections) and mental health issues (e.g., depression, anxiety, PTSD). Cultural nuances influenced HT decision-making and interactions with healthcare providers, necessitating culturally competent care.<br><br>**Recommendations:** Mandatory training programs for clinicians on trafficking indicators and trauma-informed care. Development and implementation of standardized |

| | | | | | |
|---|---|---|---|---|---|
| | | | | | protocols to improve clinician readiness and resource availability. Inclusion of cultural sensitivity in training to address barriers and improve patient-clinician trust. |
| Albright, K. 2020 (91)<br><br>U.S.<br><br>**Systematic review** | Analysis of barriers and facilitators for the identification and care of child victims of trafficking in healthcare entities | **Contextual adaptation and customization**<br>Barrier and facilitator assessment: This study emphasizes the identification of specific challenges, such as resource limitations and language barriers, and the adaptation of implementation methods to address these issues efficiently and effectively. | This systematic review aimed to summarize facilitators, barriers, and recommendations for providing medical and mental health care to trafficked children globally, focusing on English-language peer-reviewed studies published since 2010. It included 29 articles meeting criteria such as focusing on individuals under 18 years old and examining trafficking-related health services, barriers, or facilitators.. Data were systematically extracted and analyzed using qualitative content analysis, categorizing findings into five domains: survivors, individual healthcare providers, healthcare organizations, other agencies, and societal structures. Reliability was ensured through inter-reviewer and inter-coder consensus. Findings were reported based on their locus of control, offering actionable insights for improving care. | Healthcare professionals / Pediatric healthcare centers | **Facilitators of Care:** Identified 45 facilitators of healthcare service delivery for trafficked children, cited 140 times across the reviewed studies. Most facilitators fell under the domains of healthcare providers (e.g., trauma-informed training) and healthcare organizations (e.g., availability of interpreters and age-appropriate communication tools).<br><br>**Barriers to Care:** Identified 118 barriers, cited 174 times, including: Lack of provider training and awareness. Survivors' fear of judgment, confidentiality breaches, or being misunderstood.<br>Structural challenges like geographic inaccessibility or limited clinic hours.<br><br>**Recommendations for Improvement**: A total of 52 distinct recommendations, cited 100 times, focusing on: Expanding trauma-informed and culturally sensitive training for providers. Enhancing access through better resource allocation and service coordination. Developing child-specific, multidisciplinary care models.<br><br>**Domains of Change:** Most facilitators, barriers, and recommendations were within the control of healthcare providers and organizations, highlighting these groups' pivotal roles. Broader societal and structural issues (e.g., stigma, systemic inequities) were also noted but less emphasized.<br><br>**Global Perspective:** Findings reflect healthcare challenges and solutions for trafficked children worldwide, with applicability across diverse healthcare settings. |
| Peck, J.L. 2020 (92)<br><br>U.S. | Analysis of practices in pediatric healthcare settings for identifying child | **Organizational strategies**<br>Organizational culture: This analysis contributes to fostering a culture that supports trafficking detection through improved policies, leadership, and | This study aimed to examine primary care clinicians' knowledge, attitudes, and behaviors regarding the identification and assistance of HT in healthcare settings, addressing cultural influences and barriers. | Pediatric healthcare providers / Clinical and healthcare settings | **Clinician Knowledge and Awareness**: Clinicians demonstrated low awareness and limited knowledge about identifying HT. Lack of training was a significant barrier to effective recognition and intervention. |

| | | | | | |
|---|---|---|---|---|---|
| **Systematic review** | victims of trafficking | awareness-building in pediatric healthcare settings.<br><u>Quality improvement</u>: By reviewing and assessing current practices, this strategy supports continuous improvement processes aimed at refining the use of detection tools and enhancing overall effectiveness. | Using a systematic review guided by PRISMA, the authors searched PubMed, Medline Plus, and CINAHL (2016–2021). Of 130 initial articles, 10 quantitative, English-language, peer-reviewed studies focused on clinicians' roles in identifying and assisting HT were included, with data extraction and synthesis conducted independently by the authors. | Dallas, Texas. | **Attitudes and Confidence**: Many clinicians lacked confidence in their ability to identify and assist HT victims. Training improved clinicians' preparedness and confidence in handling HT-related cases.<br><br>**Barriers to Identification**: Cultural biases, unconscious stigmas, and misconceptions about trafficking hindered accurate identification. Limited access to resources and inadequate organizational support further impeded intervention efforts.<br><br>**Training Impact:** Evidence-based training programs significantly improved knowledge, skills, and confidence among healthcare providers. Trauma-informed and culturally responsive approaches were recommended for effective training.<br><br>**Recommendations**: Development of standardized clinical guidelines and screening tools. Integration of HT-related training into healthcare curricula and continuing education programs. Emphasis on multidisciplinary collaboration and organizational support to enhance detection and care. |
| Armstrong, S. 2019 (59)<br><br>U.S. | Analysis of the response of hospitals for the identification and care of trafficking victims | **Organizational Strategies**<br><u>Organizational Culture:</u><br>The study explores the absence of organizational policies and protocols in South Carolina hospitals, emphasizing the need for leadership training and fostering a supportive environment for victim identification.<br>**Contextual Adaptation and Customization**<br><u>Barrier and Facilitator Assessment:</u> By identifying barriers such as limited staff training, lack of resources, and low prioritization of trafficking issues, as well as potential facilitators like leadership engagement, the study underscores | The study aimed to evaluate the preparedness of South Carolina hospitals to identify and care for trafficked individuals, establishing baseline data and identifying unmet needs. It employed a qualitative descriptive design with stratified purposive sampling based on reported trafficking cases from the National Human Trafficking Hotline's 2016 heat map. Data were collected through structured telephone interviews with ED directors/managers and analyzed using qualitative and content analysis to uncover patterns and themes. | ED directors or managers/ Urban, suburban, and rural hospitals<br><br>South Carolina, U.S. | South Carolina hospitals were largely unprepared to address HT, with most lacking formal policies, protocols, or training for healthcare professionals. While 72.2% of participants believed trafficking occurred in their area, only 22.2% had cared for a confirmed victim, mostly related to sex trafficking. Identification relied on patient disclosures and behavioral indicators, but responses to suspected cases were inconsistent and ad hoc. Key barriers included lack of resources, knowledge, and prioritization. Few hospitals partnered with local organizations, and safety concerns for victims and staff were common. Overall, hospitals demonstrated significant gaps in readiness, highlighting the need for improved training, protocols, and community collaboration. |

| | | | | | |
|---|---|---|---|---|---|
| | | the importance of assessing and addressing these factors to improve implementation. | | | |
| Dols, J. 2019 (93)<br><br>U.S. | Analysis of Strategies to Identify HT Victims in Hospitals | **Contextual Adaptation and Customization**<br>Barrier and Facilitator Assessment: The study identifies barriers such as lack of staff training, misconceptions about trafficking indicators, absence of standardized tools, and legal limitations (e.g., privacy laws). It also acknowledges facilitators, including staff interest in improving practices and the existence of external tools that can be adapted.<br>**Organizational Strategies**<br>Organizational Culture: The study reflects a gap in organizational culture regarding awareness and prioritization of HT screening. Limited staff education and a lack of systematic approaches indicate that identifying and intervening in trafficking cases is not yet embedded in the core culture of these healthcare organizations. However, interest among leaders suggests potential for cultural shifts with proper interventions. | The study aimed to explore and document the strategies employed in EDs across 47 South Texas counties for identifying, assessing, and intervening in cases of HT. Using a descriptive survey design, researchers developed a 23-question survey based on literature and ED practices, targeting ED leaders. Data was collected through online tools, emails, and phone interviews, with implied consent through survey completion. The analysis centered on the methods and outcomes of screening practices for HT victims. | ED leaders/ EDs in 47 counties, including urban, suburban, and rural areas.<br><br>Texas, U.S. | **Survey Response:** Out of 99 EDs surveyed, 27 (27.3%) ED leaders responded.<br><br>**Screening for Adults:** 11 EDs (40.7%) screened adults for HT. Most used safety-related questions during triage to identify potential victims. 59.3% of EDs did not formally screen adults for trafficking. No EDs identified new adult trafficking victims in 2017.<br><br>**Screening for Children:** 10 EDs (37.0%) screened children for trafficking. Screening methods often mirrored those used for adults, including general safety assessments. 63.0% of EDs did not screen children specifically for trafficking. One ED identified 10 child trafficking victims in 2017, all referred by external agencies.<br>**Barriers:** Lack of standardized protocols and validated tools for trafficking victim identification.<br>Limited education and training on HT for ED staff.<br><br>**Actions Upon Identification:** Common actions included reporting to police (30.8%) or consulting social workers (15.4%). Few EDs provided referrals to shelters or community resources.<br><br>**Challenges:** Variability in screening practices across EDs. Limited awareness and misconceptions about trafficking indicators among staff.<br><br>**Opportunities Identified**: High interest among ED leaders in improving HT screening and intervention protocols. |
| Franklin, A. 2018 (94)<br><br>UK | Analysis of Strategies to Identify HT Victims in Hospitals | **Organizational strategies,**<br>Quality improvement: Evaluation cycles to monitor and enhance the use of detection tools, aligning with continuous improvement processes. | The study explores the development and use of tools and checklists for assessing the risk of child sexual exploitation (CSE) in the UK, aiming to evaluate their effectiveness in identifying potential victims, understand their application | Professionals from various agencies involved in safeguarding children/ social services, law enforcement, | **Lack of solid evidence**: The tools used are not supported by rigorous research nor validated to measure their effectiveness in identifying CSE victims.<br><br>**Variability in tools:** There is a wide diversity of tools with inconsistent indicators and a lack of standardization in their application. |

| | | | | | |
|---|---|---|---|---|---|
| | | | across agencies, and provide recommendations for improvement. Using a rapid evidence assessment to review existing literature on risk and protective indicators and conducting surveys and interviews with 42 professionals from fields such as social care, police, healthcare, and education, the study highlights the strengths, limitations, and practical use of these tools in safeguarding children and mitigating risks. | healthcare, education, and voluntary organizations.<br><br>England and Wales. | **Confusion between risk and harm:** Tools mix indicators of actual harm with risk factors, potentially leading to inadequate responses for children already being exploited.<br><br>**Reliance on scoring systems:** Some tools depend on scoring methods that do not always reflect actual risk, overlooking broader contextual information.<br>**Need for professional judgment:** While useful, tools should not replace professional expertise; a balance between both is necessary.<br><br>**Improvement proposals**: Recommendations include standardizing tools, promoting multi-agency evaluations, incorporating detailed narratives, and avoiding victim-blaming approaches.<br><br>**Limited training:** Professionals require further education on the complexity of CSE and the proper use of tools. |
| Long, E. and Dowdell Eb. 2018 (95)<br><br>UK | Analysis of Perceptions of HT and Its Identification Among Emergency Nurses | **Organizational strategies**<br><u>Organizational culture:</u><br>The recognition by nurses of the need to identify HT victims and the lack of specific policies suggests a necessary strategy to build an organizational culture that supports detection. This includes the need for organizational policies and leadership that foster an environment conducive to addressing this issue. | This study explored emergency nurses' perceptions of HT victims, including victims of violence and prostitution, and how these views influence their identification and care. Using a descriptive qualitative design, the researchers conducted semi-structured interviews with 10 registered nurses in an urban ED, analyzing the audio-recorded and transcribed data thematically through manual coding and independent reviews to ensure credibility. | 10 Bachelor of Science in Nursing (BSN) degree / An urban academic emergency<br><br>Philadelphia, Pennsylvania, U.S. | **HT awareness:** Nurses recognized the existence of HT in their patient population but lacked experience in screening or knowingly treating victims.<br><br>**Stereotypes of trafficking victims**: Victims were perceived as predominantly young, female, and foreign-born, influenced by media portrayals.<br><br>**Screening for interpersonal violence (IPV):** Nurses consistently screened for IPV and felt confident in their ability to identify such cases, unlike HT victims.<br><br>**Perceptions of victims**: IPV victims were described as "sad and grieving" or emotionally fragile. Prostitutes were perceived as "hard and tough," with little recognition of potential trafficking involvement.<br><br>**Lack of education**: Participants reported no formal training on HT, relying on IPV-related education, which was insufficient for addressing trafficking-specific needs. |

| | | | | | |
|---|---|---|---|---|---|
| | | | | | **Resource gaps**: Nurses highlighted a lack of specific resources for trafficking victims, particularly during night shifts when social workers were unavailable.<br><br>**Desire for education**: Participants expressed interest in additional training to better identify and care for HT victims. |
| Stoklosa, H. 2016 (60)<br><br>U.S. | Analysis of protocols used in healthcare entities | **Organizational Strategies**<br>Organizational Culture: Through the use of protocols, a culture that supports the detection of HT can be created.<br><br>**Contextual Adaptation and Customization**<br>Setting-Specific Adaptation: Protocols need to be adapted to the specific needs of different healthcare settings, such as EDs or pediatric care.<br>Barrier and Facilitator Assessment: Reviewing protocols allows organizations to identify potential barriers (e.g., staff workload, lack of training) and facilitators (e.g., leadership support, availability of resources) for successful implementation and adjust the approach accordingly. | The aim of the study was to characterize and assess the protocols for identifying, treating, and referring victims of HT in U.S. healthcare institutions. It analyzed 30 protocols from 19 states and 2 national organizations.<br>The strategies used included collecting protocols from hospitals, clinics, and healthcare entities through respondent-driven sampling. The protocols provided various indicators to identify HT, such as physical or sexual abuse, dependency on others, and medical symptoms. Additionally, they offered guidance on how to act in suspected trafficking cases, including contact information for local organizations and the definition of HT. | Healthcare professionals / Healthcare institutions | The findings demonstrate the varied inclusion of indicators and resources across protocols, highlighting areas where more comprehensive guidance could be implemented to improve the identification and treatment of HT victims.<br><br>**Indicators of HT:**<br>Physical or Sexual Abuse: Present in 73% of protocols.<br>Medical Symptoms: Found in 70%, including bruises (57%) and malnutrition (60%).<br>Dependence on Another Person: Noted in 70%, including lack of control over identification (63%).<br><br>**Communication Signs:** Seen in 70%, with 50% mentioning inconsistencies in stories.<br>Mental Health**:** 63% noted depression and PTSD signs.<br>Sexual History: 63% included factors like STI history (53%).<br>Housing & Appearance Indicators: 60% and 47%, respectively.<br>Technology Indicators: Present in 20%, such as explicit photos.<br><br>**Information and Guidance:**<br>Resources and Hotlines: 83% provided local contacts.<br>Definition of HT: 60% defined it; 50% explained trafficking types.<br>Mandatory Reporting: 67% addressed laws, especially for youth.<br>Screening and Clinician Guidance: 67% provided screening questions, 57% listed involved clinicians.<br><br>**Missing Information:**<br>Screening Accompanying Individuals: 80% omitted this.<br>Focus on EDs: 20% were emergency-specific. |

| | | | | | |
|---|---|---|---|---|---|
| | | | | | Child Indicators: 33% included children .<br>Follow-up Guidance: 33% mentioned safety plans, 23% follow-up. |
| Beck, M. E. 2015 (96)<br><br>U.S. | Analysis of Perceptions and Knowledge on the Identification of Trafficking Victims in Healthcare Settings | **Outcome Evaluation and Continuous Improvement**<br>Usage data monitoring: The study evaluates the effectiveness of training by comparing outcomes (e.g., knowledge and confidence) between trained and untrained providers, aligning with monitoring and evaluation practices.<br>**Contextual Adaptation and Customization**<br>Barrier and facilitator assessment: Barriers to the identification and care of trafficking victims by healthcare workers are evaluated. | The study aimed to identify knowledge gaps and training needs among medical providers regarding pediatric victims of sex trafficking (ST), emphasizing the importance of training to address their specific needs and overcome barriers to recognition and response. Using a survey distributed to physicians, nurses, social workers, and other providers in urban, suburban, and rural hospitals and clinics in southeastern Wisconsin, the research collected demographic data, assessed clinical vignettes, and analyzed knowledge and perceptions about ST through statistical methods to uncover trends and correlations. | Medical providers / multiple hospitals and medical clinics in urban, suburban, and rural locations.<br><br>southeastern Wisconsin, U.S | **Knowledge Gaps:** 63% of participants had never received training on identifying or assisting victims of sex trafficking. Only 48% correctly identified a minor as a trafficking victim in a clinical vignette. 42% adequately distinguished between a trafficking case and child abuse. The main barriers to recognizing victims were lack of training (34%) and lack of awareness (22%).<br><br>**Training Background:** Participants with prior training were significantly more likely to: Recognize trafficking as a local issue (68% vs. 45%; $p \leq 0.001$). Encounter victims in their clinical practice (75% vs. 49%; $p \leq 0.001$). Feel confident in identifying victims (10 points vs. 8 points average knowledge score; $p \leq 0.001$). No participants from primary care or urgent care clinics reported having received training.<br><br>**Actions Taken with Victims:** 69% of respondents who identified victims contacted child protective services or local police. Only 14% of medical providers, compared to 45% of social workers, contacted national hotlines or referred victims to specialized services.<br><br>**Myths and Misconceptions:** 85% knew trafficking does not require movement across borders. 90% understood that initial consent does not negate a victim's trafficking status. 10% incorrectly labeled a minor as a "prostitute" instead of a trafficking victim.<br><br>**Identified Barriers:** Major obstacles to identifying and responding to victims included: Lack of training (47% among untrained participants). Lack of awareness about the issue. Absence of clear organizational policies and standardized protocols.<br><br>**Confidence and Competence**: Participants with higher confidence in identifying victims scored significantly better on knowledge questions ($p \leq 0.001$). |

| | | | | | |
|---|---|---|---|---|---|
| Lumpkin, C.L. and Taborda, A. 2017 (97)<br><br>U.S. | Interview with survivors to improve identification and referral processes | **Patient engagement and empowerment**<br>Patient-reported indicators | A survey was conducted with survivors to assess victims' access to health services and the ability of staff to identify and refer them to other services, in order to improve healthcare training processes.<br>A structured questionnaire of 20 questions was developed for the purpose of this study, titled The Identification and Referral in Health Care Settings survey. | Survivors of labor and sex trafficking | **Respondents:** 55 individuals, with 54% survivors of sex trafficking, 42% labor trafficking, and 4% both.<br>Trafficking Duration: 61.8% trafficked for 1-5 years, 20% for over 5 years; 24 were trafficked in California.<br><br>**Age at Trafficking:** 63.6% trafficked as adults, 18.2% as minors, and 18.2% as both.<br>Healthcare Access: 64% accessed healthcare during trafficking, with community clinics being the most common.<br><br>**Identification by Providers:** 96.7% were never received information on trafficking from healthcare providers; only one person was identified by healthcare providers.<br><br>**Provider's Role**: 64.3% believe healthcare providers can help identify and refer victims.<br><br>**Improvement Suggestions:** Screening questions, resources, supportive behavior, and offers to contact authorities.<br><br>**Screening Questions**: 81% were never asked suggested screening questions; key helpful questions differed by trafficking type.<br><br>**Additional Needs:** Suggested more questions for minors and noted behaviors that may indicate trafficking. |
| Family Violence Prevention Fund In Partnership with the World Childhood Foundation, 2005 (98)<br><br>U.S. | Analysis of the experiences and needs of HT survivors and identification of barriers, facilitators, and opportunities for improving identification and care in healthcare systems | **Organizational strategies:**<br><u>Organizational culture:</u> The strategy aims to build an organizational culture that supports trafficking detection through training healthcare professionals, implementing policies, and raising awareness within healthcare systems. | The report analyzes the data that emerged from interviews with 21 victims of trafficking who were brought to the U.S. to serve as unpaid domestic and sex workers, unpaid restaurant helpers, sweatshop workers, and in one case, as a wife forced into a servile marriage.<br>This study focused on understanding the health care needs and rights of trafficking victims, identifying if the health care setting is appropriate for screening and intervention, and exploring public policy | Healthcare professionals, Government agencies and social organizations, Police and security forces, Researchers and academics / Healthcare systems | **Limited access to medical care:** Victims had limited contact with healthcare providers, often controlled by traffickers, which prevented meaningful communication.<br><br>**Barriers to victim identification:** Victims faced isolation, fear, and manipulation, making it difficult for them to recognize they were trafficked.<br><br>**Need for healthcare sector training:** Many healthcare professionals lack training to identify trafficking victims, highlighting the need for specific protocols.<br><br>**Physical and psychological impact:** Victims suffer from severe health issues, including STDs, malnutrition, and |

| | | | | | |
|---|---|---|---|---|---|
| | | | opportunities to improve health care for trafficked women and children. | | emotional trauma, especially minors who struggle to recognize their situation.<br><br>**Effective interventions:** Clear intervention protocols, such as separating victims from traffickers, could have enabled identification and assistance.<br><br>**Key recommendations:** The study recommends awareness programs, improved healthcare protocols, and better coordination between healthcare, law enforcement, and social services. |
| **Evaluation of the Implementation of a Tool or Protocol** | | | | | |
| Roe-Sepowitz, D. 2024 (99)<br><br>U.S. | Evaluation of the implementation of a screening tool for HT<br><br>Provide privacy, Educate, Ask, Respect, and Respond (PEARR) tool | **Training and Support for Healthcare Workers**<br>Formal training: Structured educational modules on HT, trauma-informed care, and the PEARR Tool provide formal training for healthcare workers.<br>**Contextual Adaptation and Customization**<br>Setting-specific adaptation: Adjustments to training formats (e.g., PDFs, mini-trainings) and the availability of informational posters in staff areas demonstrate adaptation to pandemic-related constraints and hospital-specific needs.<br>**Supervision and Performance Monitoring**<br>Managerial oversight: Monitoring the implementation process and providing guidance through multidisciplinary task force teams ensures the integration of the PEARR Tool into workflows. | The study evaluated the implementation and impact of the PEARR Tool—a structured guide for healthcare professionals to assist patients experiencing violence, including HT—in three Dignity Health hospitals in central California.<br>The implementation of the PEARR Tool in three Dignity Health hospitals in central California included its integration into policies addressing abuse and violence, supported by educational modules on HT and trauma-informed care. Due to the COVID-19 pandemic, in-person training was adapted to PDF formats and mini-trainings. It was complemented by informational posters, surveys to assess the impact, reference cards for staff, and guidelines integrated into the electronic health system. Multidisciplinary teams promoted its adoption and access to supporting materials. | ED staff / EDs of three hospitals within the U.S. | **Increased awareness**<br>Greater perception of HT at the local, national, and patient levels.<br><br>**Limited use**<br>Most did not use the PEARR Tool due to not identifying victims, although those who used it found it valuable.<br><br>**Available resources**<br>Improved perception of workplace resources and support availability.<br><br>**Educational impact**<br>Training enhanced staff understanding and readiness despite pandemic challenges.<br><br>**Step adherence**<br>More staff followed PEARR Tool steps, including privacy, education, and referral to resources. |
| Duke, D. 2023 (100)<br><br>U.S. | Implementation of software for the identification of HT victims | **Information and Communication Technology (ICT)** | The primary aim of this study was to explore a technological solution using automated informatics to identify HT victims in real time | Emergency room settings | During the observation period (2019-2021), the Octavia software generated alerts for 1 to 8 potential cases per day (out of an average of 440 daily encounters). |

| | | | | | |
|---|---|---|---|---|---|
| | | Health Information Systems: The human trafficking detection software, such as Octavia, is a system based on information technology that manages and stores clinical and social data of patients, helping to identify potential trafficking victims through the automated analysis of electronic health records (EHR).<br>Use of Information and Communication Technology: The software allows for transferring information and supporting healthcare delivery, utilizing technology-based tools to identify risk patterns associated with HT in real-time.<br><br>**Organizational Strategies:**<br>Implementation Strategies - Continuous Quality: The use of the software contributes to continuous improvement processes, as it allows for evaluation cycles to adjust and enhance the detection tools based on data gathered. | within hospital and emergency room settings.<br><br>Strategy used: A software application called Octavia was implemented in three hospitals in California. This application scanned all patient encounters for social and clinical determinants that matched predictive patterns of HT. Encounters that matched these patterns were reviewed by a High-Risk Patient Navigator (HRPN), who was specially trained to identify potential victims and, when possible, made contact with them to offer assistance. | | Of the alerts generated by the Octavia software, 43.17% were reviewed by a High-Risk Patient Navigator (HRPN).<br><br>Of the reviewed cases, 24% were classified as "highly suspicious" or confirmed as victims of HT. In total, 184 high-suspicion cases were identified during the 23-month observation period.<br><br>Comparison with Pre-Implementation Baseline: Before Octavia's implementation, during 2017 and 2018, the hospitals involved only identified an average of 10 cases per year of potential HT victims.<br><br>The response from HRPNs was affected by the COVID-19 pandemic, limiting their capacity to review all alerts generated. For example, in September 2020, no alerts were reviewed due to staffing shortages caused by the pandemic response. This is reflected in the 47.3% of alerts being reviewed during that period. |

| | | | | | |
|---|---|---|---|---|---|
| Smirnoff, M. 2022 (101)<br><br>U.S. | Feasibility assessment of implementing a screening tool for HT victims.<br><br>**RAFT (Rapid Appraisal for Trafficking)** | **Contextual adaptation and customization:**<br>Setting-specific adaptation:<br>The study evaluated the implementation of the RAFT tool specifically in the ED, focusing on its integration into the unique workflow and operational constraints of this setting. Participants identified the ED as a critical point for addressing trafficking due to its role as a "front door" for healthcare access.<br>Barrier and facilitator assessment: The study explicitly identified barriers such as time limitations, lack of privacy, staff workload, and insufficient awareness of trafficking protocols. Facilitators included staff willingness to screen, the feasibility of a short tool, and the importance of education and training. | The study evaluated the feasibility of implementing RAFT (Rapid Appraisal for Trafficking), a validated four-question screening tool for labor and sex trafficking, in an ED, while identifying barriers and facilitators to its integration into routine care. Using a qualitative design and a phenomenological analysis, eight ED staff and leaders (including service directors, nurses, physicians, and a social worker) were recruited through purposive and snowball sampling. Semi-structured interviews, conducted between July and September 2019, lasting 30–45 minutes each, were transcribed, manually coded, and analyzed using thematic analysis with Dedoose software. | ED leadership and clinical staff / A tertiary, academic ED in an urban location | Three key themes emerged:<br>**Appropriateness:**<br>All participants agreed that trafficking screening aligns with the ED's mission, as the ED is often the first point of contact for addressing social issues like intimate partner violence.<br>Staff viewed a brief screening tool with fewer than five questions as feasible, with primary responsibility falling on nurses during triage.<br><br>**Dissonance:**<br>Participants identified practical challenges, including:<br>Limited staff time and privacy for sensitive conversations.<br>Reliance on recognizing "red flags" due to operational constraints.<br>Concerns about causing offense or discomfort to patients.<br>Some participants highlighted the lack of confidence in follow-up actions after a positive screen.<br><br>**Education:**<br>A strong need for staff education on trafficking indicators, existing protocols, and available resources was emphasized.<br>Participants suggested that education should also target patients through posters and signs to facilitate self-identification and access to resources. |
| Chen, E. Y.2023 (102)<br><br>U.S. | A comprehensive care model for serving survivors of HT.<br><br>**BCM Anti-Human Trafficking Program (BCM A-HTP)** | **Organizational Strategies**<br>Organizational Culture: Promotes a supportive culture for trafficking identification through training and a favorable environment for survivor care.<br>Quality Improvement: Implements continuous improvement processes to assess and adjust its detection and care practices for victims.<br>Resource Allocation: Allocates specialized personnel, time, and | A pilot program in Houston, Texas, led by the Baylor College of Medicine in collaboration with the local government, hospitals, social service agencies, and support groups, aimed at addressing HT from a public health perspective. The program, called the BCM Anti-Human Trafficking Program (BCM A-HTP), is based on four main pillars: | The program targets healthcare professionals in clinical settings, along with support organizations, government agencies, and public health researchers involved in identifying and | **Increased Detection:** Training healthcare staff resulted in a significant increase in the detection of HT cases in the associated clinical settings.<br><br>**Improved Quality of Care:** The implementation of trauma-centered care improved the quality of services offered to survivors, providing them with a safe and supportive environment during clinical visits.<br><br>**Effective Collaboration:** The program established effective collaborations with support organizations, facilitating access to essential services for survivors, such as legal assistance and shelters. |

| | | | | | |
|---|---|---|---|---|---|
| | | specific tools for training and trauma-informed care for survivors.<br><br>**Training and Support for Health Care Workers**<br>Formal Training: Provides structured training to health professionals to identify and care for trafficking cases, addressing trauma and cultural sensitivity.<br>Communities of Practice: Facilitates the exchange of experiences and best practices among healthcare professionals through a collaborative network.<br><br>**Community and Inter-Agency Collaboration**<br>Referral Systems: Collaborates with support organizations and social services to facilitate the referral of survivors to necessary services.<br>Inter-Agency Information Exchange: Establishes joint practices and trains healthcare staff in collaboration with local organizations for the identification and referral of victims.<br><br>**Outcome Evaluation and Continuous Improvement**<br>Usage Data Monitoring: Collects and analyzes data on the usage and effectiveness of its detection tools.<br>Patient Impact Evaluation: Measures the success of referrals and the impact on survivors' recovery. | Education: Training healthcare professionals to identify and respond appropriately to cases of HT through workshops and training modules covering clinical indicators, trauma-informed interviewing, and treatment plans.<br><br>Clinical Care: Providing patient-centered, trauma-focused care with mental health services and access to follow-up care. The program includes a postdoctoral fellowship in psychology specializing in HT and social work staff.<br><br>Advocacy: Collaborating with local and national organizations to coordinate support services and meet basic needs of survivors, such as shelter, medical, and legal services.<br><br>Research: Conducting studies to improve understanding of trafficking as a public health issue and to develop evidence-based treatment guidelines.<br><br>This integrated model seeks not only to identify and treat survivors of HT but also to serve as an example for other cities to replicate and enhance this public health approach in the fight against trafficking. | supporting trafficking victims. | **Ongoing Research:** Valuable data was collected for future research to refine screening and treatment protocols and to improve public health practices related to trafficking.<br><br>**Model Replicability:** Preliminary results suggest that the model could be replicated in other cities to strengthen the public health response to HT. |
| McDow and Dols, 2021 (103) | Development and | **Organizational Strategies** | A quality improvement project at a nonprofit crisis pregnancy center in | The program targets healthcare | **Detection Outcomes:**<br>Out of 309 women screened over 10 weeks, 14 (4.6%) had |

| | | | | | |
|---|---|---|---|---|---|
| U.S. | implementation of a standardized screening protocol for the identification of HT victims among women seeking prenatal care.<br><br>**Polaris Medical Assessment Tool (2010)**<br><br>**U.S. Department of Health and Human Services (USDHHS) Adult Human Trafficking Screening Toolkit (2018)** | Quality Improvement: Implementation of a standardized screening protocol with flowchart, confidential questionnaire, and EHR integration to enhance HT detection through ongoing evaluation and staff feedback.<br><br>**Training and Support for Healthcare Workers**<br>Formal training: Structured sessions for healthcare staff on HT indicators, risk factors, and protocol use, including trauma-informed approaches and case-based discussions.<br><br>**Information and Communication Technology (ICT)**<br>Health information systems: Updates to the EHR system incorporated HT screening prompts and documentation fields to support standardized data collection and facilitate follow-up care. | the United States aimed to enhance the identification of human trafficking (HT) victims among women seeking prenatal care. The strategy was structured around four core components:<br>Education: Staff and volunteers received formal training on HT indicators, trauma-informed care, and the use of a standardized screening protocol.<br>Screening: A confidential five-question tool, guided by a flowchart and integrated into clinical workflows, enabled consistent identification of potential HT victims.<br>Resource Referral: Women who screened positive were discreetly connected to legal, medical, and social support services, with hotline information made available in private spaces.<br>Sustainability: The protocol was embedded in the electronic health record system, and ongoing training procedures were established to ensure long-term implementation. | providers, ultrasound technicians, nursing assistants, and volunteers working in a nonprofit crisis pregnancy center that offers free services to women experiencing unplanned pregnancies. | positive responses. Among them, 5 (35.7%) were confirmed as HT victims, 3 (21.4%) reported abusive relationships, and 6 (42.9%) were classified as at risk.<br><br>**Patient Demographics:**<br>Most of the 14 were aged 20–29 (57.1%), Hispanic (42.9%), had annual incomes under $15,000 (78.6%), and were pregnant (85.7%). Common indicators included being threatened (reported by 7 women) and restricted freedom (6 women).<br><br>**Tool Completion and Feasibility:**<br>Of the 309 clients, 98.4% completed all screening questions. The protocol proved feasible and well integrated into clinical workflow.<br><br>**Training Outcomes**:<br>All clinical staff (100%) and most volunteers (62.5%) completed training. Post-training, 93.3% felt confident using the protocol.<br><br>**System Integration and Support:**<br>The screening tool was embedded in the EHR. All 14 identified patients received the national HT hotline number, and 2 received additional tailored resources. |
| Tiller, J. 2020 (104)<br><br>U.S. | Development and implementation of a protocol to recognize and assist HT victims in an ED setting. | **Organizational Strategies**<br>Organizational Culture: Promotes a supportive culture for trafficking identification through training and a favorable environment for survivor care.<br>Quality Improvement: Implements continuous improvement processes to assess and adjust its detection and care practices for victims.<br>Resource Allocation: Allocates specialized personnel, time, and | The study employed a practice-based approach by utilizing a published toolkit informed by existing guidelines, expert recommendations, and models from other centers to implement an ED response protocol for HT. The methodology involved understanding the local trafficking problem, networking with anti-trafficking organizations, collaborating across specialties, and developing a concise protocol | Physicians, nurses, and ancillary staff from EDs / EDs | **Protocol steps:**<br>Step One: Understand HT and Health Generally and Locally.<br>Step Two: Understand How Survivors Gain Assistance from Non-Medical Stakeholders in the Community.<br>Step Three: Organize the Medical Community to Provide a Safety Net for Survivors.<br>Step Four: Create and Convene an Interdisciplinary Protocol Committee.<br><br>**Protocol Components:**<br>Identifying Patients at Risk for Trafficking<br>Interviewing High-Risk Patients. |

| | | | | | |
|---|---|---|---|---|---|
| | | specific tools for training and trauma-informed care for survivors. | focused on identifying at-risk patients, applying trauma-informed care, documenting appropriately, and providing resources for patients beyond medical care. | | Safety Considerations.<br>Procedures for External Reporting.<br>Strategies for Responding to Patients Who Decline Assistance.<br>Procedures Regarding Documentation.<br>Guidelines for Forensic Examination. |
| Chang, K. S. G. 2015 (105)<br><br>U.S. | Evaluation of the implementation and effectiveness of the commercial sexual exploitation of children screening protocol in a clinical setting | **Outcome evaluation and continuous improvement**<br><u>Usage data monitoring</u>: The strategy included monitoring data on the protocol’s usage frequency and effectiveness in detecting CSEC, providing qualitative and quantitative analyses to assess impact.<br><u>Patient impact assessment</u>: The strategy evaluated the tool’s impact in terms of patient outcomes, including the prevalence of CSEC among those screened and the protocol's effectiveness in identifying cases. | The evaluation strategy used a retrospective cohort design, reviewing 621 medical records of female patients aged 13 to 23 in a clinic, collecting demographic information, sexual health data, and risk factors such as sexual abuse and school absenteeism. Descriptive statistics were applied to determine the prevalence of CSEC and associated risk factors. Additionally, univariate and multivariate logistic regression models were used to identify predictors of commercial sexual exploitation, considering variables such as history of sexually transmitted infections, number of sexual partners, and other concurrent risk factors. | Primary care providers at the Asian Health Services (AHS) Teen Clinic in Oakland, California / A community clinical environment focused on serving at-risk adolescents, particularly those of Asian descent and other vulnerable groups | Of the 621 female patients whose medical records were reviewed, 177 patients (28.5%) were specifically screened for commercial sexual exploitation of children (CSEC). Among the screened patients, 13 (7.3%) reported having experienced commercial sexual exploitation.<br><br>**Risk factors**<br>Statistical analyses revealed significant associations between certain risk factors and the likelihood of CSEC:<br>Patients with a history of sexually transmitted infections (STIs) were nearly 7 times more likely to have been sexually exploited compared to those without an STI history.<br>Patients with more than 2 current sexual partners were 15 times more likely to have experienced sexual exploitation.<br>Those with more than 10 lifetime sexual partners had a 19 times higher likelihood of having been sexually exploited.<br>Patients with 2 or more concurrent risk factors had a 6 times higher probability of exploitation compared to those with fewer than 2 risk factors. |
| **Use of ICD-10 Codes** | | | | | |
| Dell, N. 2023 (106)<br><br>U.S | Measuring the use of ICD-10 codes for identifying HT victims in hospitals | **Outcome Evaluation and Continuous Improvement**<br><u>Usage Data Monitoring:</u><br>The study evaluates the frequency and effectiveness of ICD-10-CM codes used to document HT in EDs, highlighting gaps in their application. This aligns with monitoring how detection tools are utilized and identifying areas for improvement.<br>**Policy and Regulatory Compliance**<br><u>Regulatory Compliance:</u> | The study aimed to identify the characteristics of U.S. ED patients documented as experiencing HT (forced labor or sexual exploitation) using ICD-10-CM codes. It analyzed data from the 2019 Nationwide Emergency Department Sample (NEDS), a 20% stratified sample of hospital-owned EDs, focusing on the ICD-10-CM codes introduced in 2018 for classifying HT. Descriptive statistical methods and logistic regression models were applied to examine sociodemographic | Healthcare professionals, public health researchers, and policymakers / EDs<br><br>U.S | **Low Prevalence:** Only 0.0016% of ED visits documented HT as an external cause of morbidity (517 cases out of 33.1 million visits).<br><br>**Types of Exploitation:**<br>Sexual exploitation (71.6%) was more frequent than labor exploitation (28.4%).<br><br>**Demographic Characteristics:**<br>The majority were female (87.3%) and minors (30.8%). Predominantly from large metropolitan areas.<br><br>**Economic Factors:**<br>Approximately 40% lived in ZIP codes with a median annual household income below $48,000. |

| | | | | | |
|---|---|---|---|---|---|
| | | The study emphasizes ethical concerns, such as stigma, confidentiality, and patient consent, which are critical for aligning with regulatory requirements on patient rights and mandatory reporting. This ensures compliance with standards governing the documentation and handling of sensitive cases like HT. | characteristics and identify correlations. | | 41% of trafficking cases involved patients covered by Medicaid.<br><br>**Limitations:**<br>ICD-10-CM codes are not being applied consistently, underestimating the actual prevalence of HT. |
| Gutfraind, A. 2023 (107)<br><br>U.S | Measuring the use of ICD-10 codes for identifying HT victims in hospitals | **Outcome Evaluation and Continuous Improvement**<br>Usage Data Monitoring: The study assesses the use of ICD-10 codes across various clinical settings, gathering data on their frequency of use and the types of exploitation reported.<br>Patient Impact Evaluation: Analysis of patient data, such as the prevalence of mental health issues, to assess the impact of identification and documentation on addressing victims' needs.<br><br>**Policy and Regulatory Compliance**<br>Regulatory Compliance: Alignment with confidentiality regulations and victims' rights, ensuring the safe and ethical use of ICD-10 codes in the context of HT. | The study analyzed the use of specific ICD-10 codes to identify cases of HT in healthcare settings in the U.S. By examining medical records that included these codes, the study aimed to assess the effectiveness of healthcare systems in detecting and documenting cases of human exploitation. | Healthcare professionals and hospital administrators / Hospitals and clinical environments in the U.S | Cases of labor and sexual exploitation were identified through the ICD-10 codes, demonstrating that the coding system can be useful for identifying victims in healthcare. The study revealed that many victims exhibited mental health issues, such as depression and anxiety, as well as physical conditions associated with exploitation.<br>There was a low frequency of code usage, indicating a need for more training in the healthcare sector to optimize identification and documentation of these cases.<br><br>**Adoption Rate:** 5.8% annual increase in code adoption, slower than the dataset's overall growth (6.7% per year).<br>Medical Providers: 1,810 providers used the codes (0.19% of total), with 77% reporting only one patient.<br>Principal Diagnosis: Codes used as the principal or admitting diagnosis in 28% of cases.<br>Patient Data: 2,793 patients, with 1,248 recently trafficked; 86% experienced sexual exploitation, 14% labor exploitation, and 0.8% both.<br>Demographics: Predominantly female (83%), insured by Medicaid (63%), median age 20 (IQR: 15–35), 21% under 15, 52% under 25.<br>Race/Ethnicity: 49% White, 35% Black, 11% Latin-American, 3% Asian-American.<br>Medical Needs: High prevalence of sexually-transmitted infections, mental health conditions (anxiety: 21%, PTSD: 20%, major depression: 18%), high ED utilization.<br>Annual Medical Costs: Mean cost $31,055 in the 12 months after diagnosis, median cost $5,254, compared to Medicaid enrollees' mean cost of $6,556 per year. |

| | | | | | |
|---|---|---|---|---|---|
| | | | | | First Report of Trafficking: 55% outside hospital/ED settings, 25% during office visits, 8% as new patients, 10% in psychiatric encounters, 4% in behavioral therapy, 25% in E.Ds. |
| Garg, A. 2022 (108)<br><br>U.S. | Measuring the use of ICD-10 codes for identifying HT victims in hospitals | **Outcome Evaluation and Continuous Improvement**<br><u>Usage Data Monitoring</u>: The study assesses the use of ICD-10 codes, gathering data on their frequency of use and the types of exploitation reported.<br><u>Patient Impact Evaluation</u>: Analysis of patient data, such as the prevalence of mental health issues, to assess the impact of identification.<br><u>Continuous Improvement:</u> Continuous monitoring and refinement of the process by which healthcare systems utilize these codes. Data analysis is used to assess the effectiveness of the implementation, identify gaps, and drive improvements in how health systems and providers recognize and report HT cases.<br><br>**Policy and Regulatory Compliance**<br><u>Regulatory Compliance</u>: Alignment with confidentiality regulations and victims' rights, ensuring the safe and ethical use of ICD-10 codes in the context of HT. | The objective of this study was to use a large, multicenter database of pediatric hospitalizations in the U.S. to describe the utilization of ICD-10-CM codes related to child trafficking, as well as the demographic and clinical characteristics of these children. | Healthcare professionals / Pediatric healthcare centers | These results indicate low utilization of ICD-10-CM codes to identify children who are victims of trafficking, especially in academic pediatric healthcare centers, suggesting a lack of awareness among healthcare providers about the scale of the issue.<br><br>**Utilization of ICD-10-CM codes:** Only 0.005% of patient encounters (293 cases) included ICD-10-CM codes related to HT.<br><br>**Patient demographics:** 90% of the patients were female. 38% were Non-Hispanic Black, and 28.3% were Non-Hispanic White. 59% of the patients had public insurance.<br><br>**Mental health disorders:** 64.8% of the patients had a mental health disorder diagnosis at the initial visit. 32.1% of the principal diagnoses were related to mental health disorders.<br><br>**Hospital readmissions:** 16% of patients (48 cases) were readmitted to the hospital within 30 days of their initial hospitalization.<br><br>**Most commonly used ICD-10 codes**: The most common code was Y07.6 (33.8%), related to "multiple perpetrators of maltreatment and neglect."<br>The second most common code was Z62.813 (21.2%), related to "personal history of forced labor or sexual exploitation in childhood." |
| Kerr, P.L. 2022 (109)<br><br>U.S | Measuring the use of ICD-10 codes for identifying HT victims in hospitals | **Outcome Evaluation and Continuous Improvement**<br><u>Usage Data Monitoring</u>: The study assesses the use of ICD-10 codes across various clinical settings, gathering data on their frequency | A retrospective data analysis using the TriNetX database, focusing on the use of ICD-10-CM codes for HT in the U.S. Clinical encounters with these codes, patient demographic data, and comorbid diagnoses were | Healthcare professionals / 48 healthcare organizations | The study by Kerr (2022) found that out of 69,740,144 patients, only 298 had HT-related ICD-10-CM codes, mostly concentrated in the Southern U.S. (40.9%). The majority of patients were young women (average age 26), primarily White (53%) or African American (28.2%). |

| | | | | | |
|---|---|---|---|---|---|
| | | of use and the types of exploitation reported.<br>Patient Impact Evaluation: Analysis of patient data, such as the prevalence of mental health issues, to assess the impact of identification.<br>Continuous Improvement: continuous monitoring and refinement of the process by which healthcare systems utilize these codes. Data analysis is used to assess the effectiveness of the implementation, identify gaps, and drive improvements in how health systems and providers recognize and report HT cases.<br><br>**Policy and Regulatory Compliance**<br>Regulatory Compliance: Alignment with confidentiality regulations and victims' rights, ensuring the safe and ethical use of ICD-10 codes in the context of HT. | analyzed through descriptive statistics. | | The most used codes were for suspected forced sexual exploitation (32.2%) and personal history of exploitation (27.1%), with labor exploitation codes applied in less than 4% of cases. Comorbid diagnoses included psychiatric disorders (69.8%), particularly depression (51.7%) and anxiety (43%), as well as PTSD (33.2%) and substance use disorders (46.3%).<br><br>These findings suggest low use of trafficking codes and underscore the need for improved training in the healthcare system for better identification of trafficking cases. |

ASH: Asian Health Services, BCM-A-HTP: Baylor College of Medicine Anti-Human Trafficking Program, BSN: Bachelor of Science in Nursing, CEST: Children exposed to sex trafficking, CMDA: Christian Medical & Dental Associations, CME: Continuing Medical Education, CSE: Child Sexual Exploitation/ Commercially Sexually Exploited, CSEC: Commercial Sexual Exploitation of Children/ Commercially Sexually Exploited Children, ED: Emergency Department, EHR: EM: Emergency Medicine, EMR: Electronic Medical Record, EMS: Emergency Medical Services, FM: Family Medicine, FNP: HCP: Health Care Providers, HOPE: Healthcare Observations for the Prevention and Eradication, HRPN: High-Risk Patient Navigator, HT: Human Trafficking, HTMSH: Human Trafficking Training, ICD-10: International Classification of Diseases, ICT: Information and Communication Technology, IOM: International Organization for Migration, IPV: Intimate Partner Violence, LIFT: Learn to Identify and Fight Trafficking, LMS: Learning Management System, MSH: Medical Safe Haven, NEDS: National Emergency Department Sample, OBGYN: Obstetrics/Gynecology, PEARR: Provide privacy, Educate, Ask, Respect, and Respond (Tool); PED: Pediatric Emergency Department, PPE: Medical-Patient Interaction Training, PTSD: Post-Traumatic Stress Disorder, RAFT: Rapid Appraisal for Trafficking, RNs: Registered nurses, SD: Standard Deviation, SOAR: Significant increases in Stop, Observe, Ask, Refer; ST: Sex trafficking, STD: Sexually Transmitted Disease, STI: Sexually Transmitted Infection, TIC: Trauma-Informed Care, U.S.: United States.

### Framework on Human Trafficking

Based on the Palermo Protocol and UNODC Resources

**Definition:** Palermo Protocol's definition of human trafficking

According to Article 3 of the Protocol to Prevent, Suppress and Punish Trafficking in Persons, Especially Women and Children, supplementing the United Nations Convention against Transnational Organized Crime:

> "Trafficking in persons shall mean the recruitment, transportation, transfer, harbouring or receipt of persons, by means of the threat or use of force or other forms of coercion, of abduction, of fraud, of deception, of the abuse of power or of a position of vulnerability or of the giving or receiving of payments or benefits to achieve the consent of a person having control over another person, for the purpose of exploitation."

**Note:** In the case of children, proof of "means" (e.g., coercion, deception) is not required to establish a trafficking offense.

***Typologies of Human Trafficking***:

- Sexual Exploitation
- Forced Labour
- Forced Begging
- Exploitation in Criminal Activities
- Organ Removal
- Forced Marriage
- Child Soldier Recruitment
- Commercial Surrogacy under Coercion

***United Nations Response Framework***

- **Prevention:** Public awareness, reducing vulnerability, regulating migration and work
- **Protection of Victims:** Identification, provision of health/legal/social services, safe reintegration
- **Prosecution:** Criminalization, investigation, prosecution of traffickers
- **International Cooperation:** Cross-border collaboration, data sharing, joint investigations
- **Monitoring & Data Collection:** National trafficking observatories, global reporting mechanisms

22. Academy of Medical Royal Colleges. Child sexual exploitation: improving recognition and response in health settings [Internet]. United Kingdom; 2014. Available from: aomrc.org.uk

23. CSEC Collaborative Response Team. Commercial Sexual Exploitation of Children (CSEC) Protocol for San Luis Obispo County. 2017; Available from: https://www.slocounty.ca.gov/departments/social-services/forms-documents/child-welfare-services/informational-documents/csec-protocol

24. CSEC Collaborative Response Team. Commercially sexually exploited children (CSEC): Screening tool user guide. 2015; Available from: https://www.cwda.org/sites/main/files/file-attachments/slo-csec-screening-tool-user-guide-3-5-15.pdf?1455665818

25. American College of Obstetricians and Gynecologists' Committee on Health Care for Underserved Women. Human Trafficking [Internet]. United States; 2011 [cited 2025 Feb 10]. Available from: https://www.acog.org/clinical/clinical-guidance/committee-opinion/articles/2019/09/human-trafficking

26. Shared Hope International. Domestic Minor Sex Trafficking Intervene Identifying and Responding to America's Prostituted Youth. Two-Part Resource Package Practioner Guide and Intake Tool & Training Video [Internet]. United States; 2010. Available from: https://www.thresholdglobalworks.com/pdfs/sex-trafficking-guide.pdf

27. National Human Trafficking Resource Center (NHTRC). Framework for a Human Trafficking Protocol in Healthcare Settings [Internet]. United States; Available from: https://humantraffickinghotline.org/en/resources/framework-human-trafficking-protocol-healthcare-settings#:~:text=This%20resource%20provides%20a%20framework%20for%20a%20human%20trafficking%20protocol#:~:text=This%20resource%20provides%20a%20framework%20for%20a%20human%20trafficking%20protocol

28. HEAL Traffi cking. healtrafficking.org. 2023 [cited 2024 Oct 14]. Guia para desenvolver uma resposta às víti mas de tráfi co de pessoas em ambientes de assistência à saúde. Available from: https://drive.google.com/file/d/1ZorgkJJwrW2R8iF3p74txOJg42HZDY-w/view

29. Hughes DM. Hiding in Plain Sight: A Practical Guide to Identifying Victims of Trafficking in the U.S. 2003 Jan 1 [cited 2025 Feb 8]; Available from: https://www.academia.edu/35381082/Hiding_in_Plain_Sight_A_Practical_Guide_to_Identifying_Victims_of_Trafficking_in_the_U_S

30. Virginia Hospital & Healthcare Association. Human Trafficking: Care and Response. A Guide for Health Care Professionals. 2021;

31. Alpert E, Ahn R, Albright E, Purcell G, Burke T, Macias-Konstantopoulos W. Human Trafficking: Guidebook on Identification, Assessment, and Response in the Health Care Setting. MGH Human Trafficking Initiative, Division of Global Health and Human Rights, Department of Emergency Medicine, Massachusetts General Hospital, Boston, MA and Committee on Violence Intervention and Prevention, Massachusetts Medical Society, Waltham, MA; 2014.

39. Connecticut Department of Children and Families. Practice Guide for Intake and Investigative Response To Human Trafficking of Children. United States; 2015.

40. Peck JL, Meadows-Oliver M, Hays SM, Maaks DG. White Paper: Recognizing Child Trafficking as a Critical Emerging Health Threat. J Pediatr Health Care. 2021 May 1;35(3):260–9.

41. HHS Office of Population Affairs (OPA) Title X Family Planning Program. Resource Guide on Human Trafficking for Title X Family Planning Clinics.

52. London Safeguarding Children Board. The London Child Sexual Exploitation Operating Protocol. 2015;

53. CommonSpirit Health PSC HEAL Trafficking. The PEARR Tool: Trauma-informed approach to victim assistance in health care settings. 2020; Available from: https://www.dignityhealth.org/content/dam/dignity-health/pdfs/pearrtoolm15nofield2019.pdf

54. Sabella D. The Role of the Nurse in Combating Human Trafficking. AJN Am J Nurs. 2011 Feb;111(2):28.

55. American Hospital Association. 10 Red Flags that Your Patient Could be a Victim. United States; 2021.

56. Polaris Project. Identifying Victims of Human Trafficking Potential Indicators & Red Flags. United States; 2010.

62. Polaris Project. On-Ramps, Intersections, and Exit Routes: A Roadmap for Systems and Industries to Prevent and Disrupt Human Trafficking. 2018;

63. Macy RJ, Graham LM. Identifying domestic and international sex-trafficking victims during human service provision. Trauma Violence Abuse. 2012 Apr;13(2):59–76.

64. Hainaut M, Thompson KJ, Ha CJ, Herzog HL, Roberts T, Ades V. Are Screening Tools for Identifying Human Trafficking Victims in Health Care Settings Validated? A Scoping Review. Public Health Rep Wash DC 1974. 2022;137(1_suppl):63S-72S.

65. Polaris Project. Medical Assessment Tool | Polaris Project [Internet]. United States: National Human Trafficking Resource Center; 2010. Available from: https://humantraffickinghotline.org/sites/default/files/Assessment%20Tool%20-%20Medical%20Professionals.pdf

66. Benavente B, Ballester Brage L, Pich Solé J, Pereda Beltrán N. Risk Factors for Commercial Sexual Exploitation of Children and Adolescents: Results of an International Delphi Panel. Psicothema. 2021 Aug;33(3):449–55.

67. Farrell SE, Bosson R, Stoklosa HM. Using Theory and Technology to Build an Interprofessional Community of Human Trafficking Educators. J Contin Educ Health Prof. 2022 May 17;10.1097/CEH.0000000000000585.

97. Lumpkin CL, Taboada A. Identification and Referral for Human Trafficking Survivors in Health Care Settings: Survey Report.

109. Kerr PL, Bryant G. Use of ICD-10 Codes for Human Trafficking: Analysis of Data From a Large, Multisite Clinical Database in the United States. Public Health Rep Wash DC 1974. 2022;137(1_suppl):83S-90S.
